# Health-economic burden of chikungunya infection and potential cost-effectiveness of preventive vaccination in 31 countries

**DOI:** 10.1101/2025.09.26.25336724

**Authors:** Junwen Zhou, Natasha Salant, Hale-Seda Radoykova, Janey Messina, William Wint, Joshua Longbottom, Katherine M Holohan, Katelyn A Dinkel, Mira L T Sytsma, Andrew A Torkelson, Luciano Pamplona de Góes Cavalcanti, T Déirdre Hollingsworth, Jennifer Lord, David R M Smith, Koen B. Pouwels

**Author notes:** Corresponding author: Junwen Zhou. These authors contributed equally (Junwen Zhou, Natasha Salant). These authors contributed equally (T Déirdre Hollingsworth, Jennifer Lord, David R M Smith, Koen B. Pouwels).

## Abstract

As the first chikungunya vaccines become available, estimates of their cost-effectiveness are needed. We developed a modelling framework to project the health-economic burden of chikungunya across 31 countries. We simulated several preventive vaccination campaigns and estimated threshold vaccination costs as the cumulative societal costs averted per vaccine dose. From 2025 to 2050, we projected a cumulative 5.96 million (5.10 million-6.93 million) DALYs and $76.5 billion ($57.4 billion-$95.7 billion) societal costs. A five-year population-wide campaign targeting 50% of individuals aged ≥12 years, followed by a ten-year annual routine adolescent vaccination programme, averted 681,000 (569,000-802,000) DALYs and $11.8 billion ($8.73 billion-$14.9 billion) societal costs. Standalone population-wide campaigns were the most cost-effective, with threshold vaccination costs ranging from $1.11 ($0.77-$1.44) in the Democratic Republic of Congo to $48.1 ($36.0-$60.3) in Panama. Preventive deployment of a chikungunya vaccine with a favourable safety profile could substantially reduce the future burden of chikungunya across endemic settings.

## INTRODUCTION

Chikungunya is a mosquito-borne disease caused by *Alphavirus chikungunya* (CHIKV), an arbovirus transmitted primarily through infected *Aedes* species. First isolated during an outbreak in Tanzania in 1952/1953^1^, CHIKV has since spread globally through international travel and trade, with outbreaks reported across Africa, Asia, the Americas, Europe and various islands in the Caribbean Sea and Indian and Pacific Oceans^2^.

CHIKV infection poses a considerable health and economic burden. Acute infection is characterised by fever and joint pain and has a similar clinical presentation to several other acute arboviral diseases, such as Zika and dengue. Approximately half of symptomatic cases are estimated to lead to chronic disease characterised by painful and persistent post-acute arthralgia^3^. The impact of chronic chikungunya on quality of life is substantial, with 94% of participants with post-acute arthralgia in a longitudinal study in Brazil reporting difficulty with ordinary daily activities, 88% reporting difficulty walking and 62% reporting mental distress^3^. The consequences for health systems and economic productivity are therefore substantial^4^.

In February 2023, the Pan American Health Organization reported an increase in reported chikungunya cases and deaths and recommended that affected states intensify preparation and response to outbreaks^5^. Recent outbreaks, including in La Réunion and China, underscore the continued epidemic potential of CHIKV^6^. In November 2023, the world’s first CHIKV vaccine, VLA1553 (Ixchiq®), was approved by the United States Food and Drug Administration, and in July 2024 received market authorisation in Europe. In early 2025, a second, recombinant CHIKV vaccine, PXVX0317 (Vimkunya®), received market authorisation in Europe and the United States. Both vaccines were approved based on immunogenicity from phase 3 clinical trials without effectiveness evidence^7,8^.

Two studies have explored the global burden of chikungunya without considering vaccination: a systematic review summarising the health burden reported from 2010 to 2019^9^, and a model-based analysis estimating the health-economic burden from 2011 to 2020^10^. Another study has modelled the transmission dynamics of Paraguay’s 2023 outbreak and assessed a counterfactual vaccination campaign^11^. Building on estimates from this study, two recent modelling analyses estimated the annual health burden of CHIKV infection and the potential health benefits of hypothetical vaccination. One analysis included global campaigns^12^ while the other focused specifically on Brazil^13^. However, no study has projected the long-term future burden of CHIKV, linked CHIKV incidence to country-level environmental suitability, or assessed the health-economic benefits of alternative vaccine strategies. Such evidence is urgently needed to guide target prioritisation and to inform vaccine investment. To support these efforts, we developed a modelling framework to project the health-economic burden of chikungunya from 2025 to 2050 and to evaluate the potential cost-effectiveness of preventive vaccination campaigns across 31 countries in 5 continents.

## METHODS

### Model overview

We developed a modelling framework to project the health-economic burden of CHIKV infection from 2025 to 2050 and to estimate the potential cost-effectiveness of a series of preventive CHIKV vaccination campaigns. The analytical time horizon was chosen to fully capture the impacts of the included vaccination campaigns (see below). This analysis focuses on 31 countries across 5 continents: Belize, Bolivia, Brazil, Democratic Republic of the Congo, Republic of the Congo, Colombia, Costa Rica, Djibouti, Dominican Republic, Ecuador, Ethiopia, Grenada, Guatemala, Guyana, Honduras,

Haiti, Indonesia, India, Cambodia, St. Lucia, Mexico, Malaysia, Nicaragua, Panama, Peru, Philippines, Paraguay, Sudan, El Salvador, Chad, Thailand. Additionally, we estimated the health burden and corresponding efficiency of vaccination programmes in Venezuela. We did not, however, estimate economic consequences in Venezuela because reliable estimates of economic parameters are unavailable due to economic collapse (i.e., hyperinflation) in recent years. These countries were selected due to reporting either substantial and recent outbreaks (>1,000 cases from 2019 to 2022, with >2,000 cases overall) or less recent but substantial outbreaks (<1,000 cases from 2019 to 2022 but >12,000 cases overall) according to reported case counts from 1952 to 2022^14^. Countries reporting only sporadic cases and overseas territories were excluded. Maldives and Tonga were excluded from the list due to unreliable estimation of CHIKV suitability across archipelagos consisting of many small islands.

The model consists of six components. First, a global suitability surface for CHIKV vectors, including both *Ae. aegypti* and *Ae. Albopictus*, was estimated using both random forest (RF) and boosted regression trees (BRTs) with vector occurrence data and environmental covariates. Second, global CHIKV suitability was estimated using an ensemble of BRTs, geolocated CHIKV infection occurrences and environmental covariates, masking by vector suitability. Third, historical force of infection (FoI) dynamics estimated in different areas of the world using age-seroprevalence data were taken from Kang *et al*^15^. Fourth, the relationship between CHIKV suitability and FoI was estimated using a logistic growth model. Fifth, time-varying infection incidence was simulated by applying annualised FoI estimates in each country from the first year of CHIKV exposure in that country to 2050, simulating accumulation and loss of age-specific seroprevalence over time and adjusting the population at risk accordingly. Finally, a decision-analytic model was used to estimate country-specific health-economic outcomes of CHIKV infection with and without CHIKV vaccination. Effectiveness estimates for vaccines such as VLA1553 and PXVX0317 are not yet available, so this study considered a hypothetical vaccine with characteristics varied in sensitivity analyses. We conservatively assumed the vaccine to have a direct impact on disease prevention but no impact on CHIKV transmission. An illustrated model schematic is provided in **Figure S1** and additional methodological details for each model component are provided in **Supplementary methods**.

### Vector suitability

The distribution of the two principal CHIKV vectors, *Ae. aegypti* and *Ae. albopictus*, was estimated using an ensemble of BRT and RF models, following the vector modelling framework by the European Centre for Disease Prevention and Control^16^. Models were trained on global vector occurrence data from VectorNet (https://www.ecdc.europa.eu/en/about-us/partnerships-and-networks/disease-and-laboratory-networks/vector-net) and the Global Biodiversity Information Facility (www.gbif.org), and included covariates selected from the H2020 MOOD environmental data suite (https://mood-h2020.eu/) (**Table SS1**): indices of vegetation greenness, day and night land surface temperature and middle infrared derived from a 2012-2021 time series, together with land use, elevation and human population density. Ensemble predictions from ten replicate BRT and RF models were combined to generate vector suitability surfaces at 5 km spatial resolution for use in subsequent analyses.

### CHIKV suitability

CHIKV suitability was estimated using BRT species distribution modelling as the probability of CHIKV occurrence in humans, also at a 5-kilometre spatial resolution. The model was trained on a global occurrence database containing 1,801 geolocated points and polygons, representing locations where at least one human CHIKV infection was reported. Occurrence data were compiled from peer-reviewed literature and ProMED Mail reports from 1952 until July 2023. Polygons larger than 2,500 square kilometres were excluded to reduce spatial uncertainty. We additionally removed four occurrence points located in high-altitude areas with an estimated temperature suitability of 0, which were likely to represent imported cases rather than local transmission. Pseudo-absences were generated and weighted based on the estimated temperature niche of *Ae. aegypti*. Model covariates included precipitation, land surface temperature and vegetation greenness, population density, and suitability surfaces for *Ae. aegypti* and *Ae. albopictus*, which were selected for their potential influence on CHIKV transmission suitability. The CHIKV suitability surface was then masked using the temperature suitability for *Ae. aegypti* and *Ae. albopictus*, ensuring that CHIKV suitability was set to zero when temperature suitability of both vectors was zero. (**Fig SS1-3**) In a sensitivity analysis, we used an alternative CHIKV suitability surface estimated by Lim *et al.*^17^ which incorporated a broader set of covariates, including socioeconomic variables. (**Table SS2**)

### Force of infection (FoI)

FoI was defined as the annual probability of a susceptible individual becoming infected using estimates from Kang *et al*.^15^ They estimated long-term average annual FoI based on cross-sectional age-stratified seroprevalence datasets covering 76 locations in 28 countries and territories. They tested both time-constant FoI and time-varying FoI models to reflect endemic transmission and epidemic outbreaks, respectively, and selected the model with the better fit to produce long-term average FoI estimates in each location. In epidemic settings, they simulated outbreak frequencies over a 100-year period to derive comparable long-term average annual FoI across locations, allowing global comparison of transmission intensity. The locations for which FoI was estimated were geolocated using Global Administrative Areas (GADM) data version 4.1 (https://gadm.org/), by matching each site to the smallest available administrative unit.

### Mapping CHIKV suitability to FoI

The relationship between CHIKV suitability and FoI was estimated to extrapolate infection incidence to areas without seroprevalence data. A logistic growth curve model was fitted using estimated suitability as the only covariate and estimated FoI as the outcome. This model was used to calculate average annual FoI from CHIKV suitability at the grid cell level. Country-level FoI was calculated as the average FoI across all cells in a country, weighted by population size. To propagate uncertainty in both suitability and FoI, the logistic growth curve model was fitted on a combination of 100 bootstrapped suitability estimates from the BRTs and 100 draws from the FoI estimates derived from Kang *et al*^15^. (**Table SS3-4, Fig SS4**)

### Projecting infections through time

As CHIKV antibodies provide long-lasting immunity^18^, seropositivity was assumed to be lifelong and only seronegative individuals were considered at risk for CHIKV infection. Annual infection incidence in each country was estimated based on the country’s reported first year of CHIKV exposure^19,20^, average annual FoI and population size. Annual country-level population estimates stratified by age from 2025-2050 were taken from the United Nations World Population Prospects^21^. Age-specific CHIKV seropositivity in 2025 was simulated by simulating annual FoI among seronegative individuals (the population at risk) in each age group from the first year of CHIKV exposure reported in each country to 2025 (**Table SS5, Fig SS5**). Annual infection incidence was then projected annually from 2025 to 2050 using this algorithm to account for time-varying change in age-specific seroprevalence. (**Table SS6**, **Fig SS6-8**)

### Age- and sex-specific disease risk

Evidence suggests CHIKV infection risk is similar regardless of age and sex^11,22^, but illness severity varies^11,23^. Age- and sex-specific risks of symptomatic infection were estimated based on data from Nicaragua (2015)^24^ and scaled to match the age and sex distribution of 635,195 cases reported in Brazil (2015-2021)^25^. In sensitivity analyses, alternative data from the Philippines (2012-2013)^26^ and the Maldives (2019)^27^ were considered. (**Table SS7-8, Fig SS9**) Symptomatic cases (all acute) were divided into detected (medically attended) and undetected groups, and detected cases were further divided into mild/moderate cases (outpatient care) and severe cases (inpatient care). In the primary analysis, undetected symptomatic cases were assumed to have milder disease severity (represented in the model by a lower disability weight) than detected mild/moderate cases, while equal severity was assumed in a sensitivity analysis. The probability of infection detection was estimated using data from Paraguay (2023), where 6.3% of all infections were detected^11,12^; and the probability of hospitalisation among detected cases was estimated using data from Brazil (2017-2025), where 3.2% of detected infections were hospitalised^28^. In a sensitivity analysis, an infection detection rate of 3.6% was assumed, matching estimates from Brazil suggesting an increase in the infection detection rate from 0.024% in 2014 to 3.6% in 2024^13^. Age- and sex-specific risks of severe illness and hospitalisation were scaled to hospitalisation patterns from Paraguay (2022-2023)^29^, while risks of death were scaled to age- and sex-specific fatality rates from Brazil (2016-2022)^23^. (**Table SS9-10**) For chronic chikungunya, a meta-analysis found 51.0% of detected symptomatic cases develop ≥3 months of symptoms^15^, applied only to detected symptomatic cases in the base-case analysis, with sensitivity analysis extending this to undetected symptomatic cases.

### Vaccine characteristics

There are no data on the effectiveness and duration of protection of currently approved vaccines, though 94.5% of adults receiving a single dose of VLA1553 maintained antibody titres above the sero-response threshold 4 years after the vaccination^30^. Therefore, we considered a hypothetical single-dose CHIKV vaccine licensed for prophylactic administration to adolescents and adults. The vaccine was assumed to provide 10 years of partial protection against disease and was conservatively assumed to have no impact on infection or transmission. In the base-case scenario, the vaccine was 70% effective against all symptomatic disease, while efficacies of 50% and 90% were considered in sensitivity analyses.

Although we modelled a hypothetical vaccine, we included vaccine-associated adverse events. No serious safety concerns have been reported for PXVX0317. However, post-marketing surveillance of the live-attenuated vaccine VLA1533 has identified rare but serious adverse events, prompting recommendations by some regulatory authorities to pause vaccination in older adults^31^. Although two severe neurological adverse events, including one fatal case^32^, have been attributed to VLA1553, we assumed that large-scale preventive vaccination programmes would only be implemented if the overall risk of severe or fatal neurological outcomes due to vaccination is negligibly small.

For the base-case analysis, we adopted a similar approach to Daniel *et al*^31^, and included age-specific probabilities of serious adverse event (SAE) using rates reported by Valneva for IXHCIQ (7/32,949 vaccinees aged 18-64 and 22/18,445 vaccinees aged 65+)^33^, assuming adolescents aged 12-17 have the same probability of event as those aged 18-64 (**Table SS23**). Consistent with Daniel *et al*^31^, we assumed that these SAEs had the same duration, disability weight and healthcare costs as medically attended acute mild/moderate chikungunya^31^. We did not assign additional vaccine-associated mortality or severe disability beyond that captured by these SAEs. In addition, we assumed no association between vaccine uptake and disease risk or serostatus.

In a scenario analysis, we modelled the broader safety profile reported for VLA1553 in randomised controlled trials^34–36^ and post-marketing surveillance^37^. Specifically, we included age-specific probabilities of mild, moderate and severe myalgia/arthralgia (**Table SS24**). These events were substantially more frequent than the SAEs included in the base-case analysis, with the highest probability observed for mild myalgia/arthralgia in 12–17 years old adolescents (0.12, 95% CI 0.07-0.18). Disability weights were assumed to match those of CHIKV symptoms of the corresponding severity, with a duration of four days based on the VLA1553’s package leaflet^38^. We additionally included chronic or prolonged vaccine-associated adverse events, stratified by severity based on post-marketing surveillance data^37^ (**Table SS24**). Finally, we performed a further scenario analysis that excluded all vaccine-associated adverse events.

### Vaccination campaigns

Three main vaccination strategies were simulated in which doses of a hypothetical CHIKV vaccine were allocated preventively to target groups living in all 31 included countries and Venezuela. The vaccination strategies include: (1) population-wide campaigns, defined as one-off vaccine administration aiming to reach a high level of vaccine coverage across a wide age range over a fixed timeline; (2) routine campaigns, defined as sustained vaccine administration aiming to reach a high level of vaccine coverage among individuals of a specific age each year; and (3) the combination of both campaign types. (**Figure S2-S3**) For population-wide campaigns, we simulated three scenarios targeting individuals aged 12-99 over five years (2025-2029), with annual target coverage of 2%, 4% and 10%, reaching, respectively, 10%, 20% and 50% target coverage after five years. For routine campaigns, we simulated one scenario targeting individuals aged 12 with an annual target coverage of 65% over 2025-2040. For combined campaigns, we combined this routine campaign with each of the three preventive campaign coverage scenarios, assuming an initial phase of general population vaccination over five years followed by routine vaccination for the remaining 11 years. In all scenarios, the number of vaccine doses delivered was reduced by 5% relative to vaccine target coverage to account for vaccine wastage (i.e. actual achieved coverage = target coverage × 95%). **Table S1** shows the total vaccine demand forecast for each scenario.

### Health and economic outcomes

The following health outcomes were included: (i) CHIKV infections, (ii) symptomatic chikungunya, defined as symptomatic CHIKV infections and stratified into acute mild/moderate symptoms or severe symptoms, (iii) chikungunya hospitalisations (due to acute severe symptoms), (iv) chikungunya deaths, (v) chronic chikungunya, and (vi) disability-adjusted life-years (DALYs) (**Table SS11-12**).

Economic outcomes were estimated in all 31 countries. The following economic outcomes were included: (i) healthcare costs, stratified by setting (outpatient/inpatient) and payer (government-reimbursed/out-of-pocket (OOP)) (**Table SS13-14**); (ii) instances of catastrophic healthcare expenditure or impoverishing healthcare expenditure resulting from OOP healthcare costs (**Table SS15-17, Fig SS10**); (iii) productivity losses due to reduced labour force participation because of acute symptomatic chikungunya, chronic chikungunya or death (**Table SS18-20**); (iv) monetised DALYs, quantified using country-specific health opportunity costs^39^ (**Table SS21**); and (v) the value of statistical life (VSL) and value of statistical life-years (VSLY) lost due to chikungunya mortality (**Table SS22**); (vi) societal costs, defined as the sum of all healthcare costs, productivity losses and monetised DALYs; and (vii) dose-weighted mean threshold vaccination costs, calculated for each vaccination scenario by dividing societal costs averted by the number of doses administered. The threshold vaccination cost represents the maximum price of the vaccine, including administration costs, at which vaccination remains cost-effective from a societal perspective. For two-dose vaccines (such as PXVX0317), the corresponding threshold vaccination costs per dose would be half the costs estimated in the current analysis for one-dose vaccines. Costs beyond the first year of the study horizon (2025) are discounted at 3.5% annually, or 0% in sensitivity analysis. All economic outcomes are reported in International dollars ($) 2023.

### Simulation and statistical reporting

Parameter uncertainty was incorporated by randomly drawing parameter values from estimated distributions for each of 1000 Monte Carlo simulations. Random parameters varied in each run include: CHIKV suitability; annual FoI estimates; and health outcome probabilities, their distributions by age and sex and their durations and associated disability weights. Final health and economic outcomes, as well as outcomes averted by vaccination, are reported as means and 95% uncertainty intervals (UIs) of outcome distributions across all simulations. Disease burden estimates are reported in accordance with the GATHER statement (**Table S2**).

All analyses were run using R version 4.3.1.

## RESULTS

### Chikungunya burden without vaccination

Population-weighted CHIKV suitability varied substantially between countries, ranging from 0.327 (95%UI 0.292-0.361) in Ethiopia to 0.974 (0.971-0.978) in El Salvador. The variability of all 5-kilometre × 5-kilometre polygon suitability within a country was smallest in El Salvador [median (Q1-Q3) 0.975 (0.968-0.980)] and largest in Malaysia [0.811 (0.129-0.961)]. (**Figure 1**) Country-level FoI was lowest in Bolivia [95%UI 0.006 (0.005-0.007)] and highest in Cambodia [0.015 (0.013-0.017)]. The predicted number of CHIKV infections in 2025 was 27.2 million (26.0 million-28.5 million) across the 31 countries and Venezuela, with the lowest in Grenada [906 (793-1,020)] and the highest in India [13.7 million (12.6 million-14.9 million)]. In the sensitivity analysis using the suitability surface from Lim *et al*^17^, the predicted FoI and infection numbers were similar in most countries, resulting in a slightly higher estimate of total CHIKV infections in 2025 across the 31countries and Venezuela [28.0 million (25.4 million-30.6 million)].

**Figure 1.**
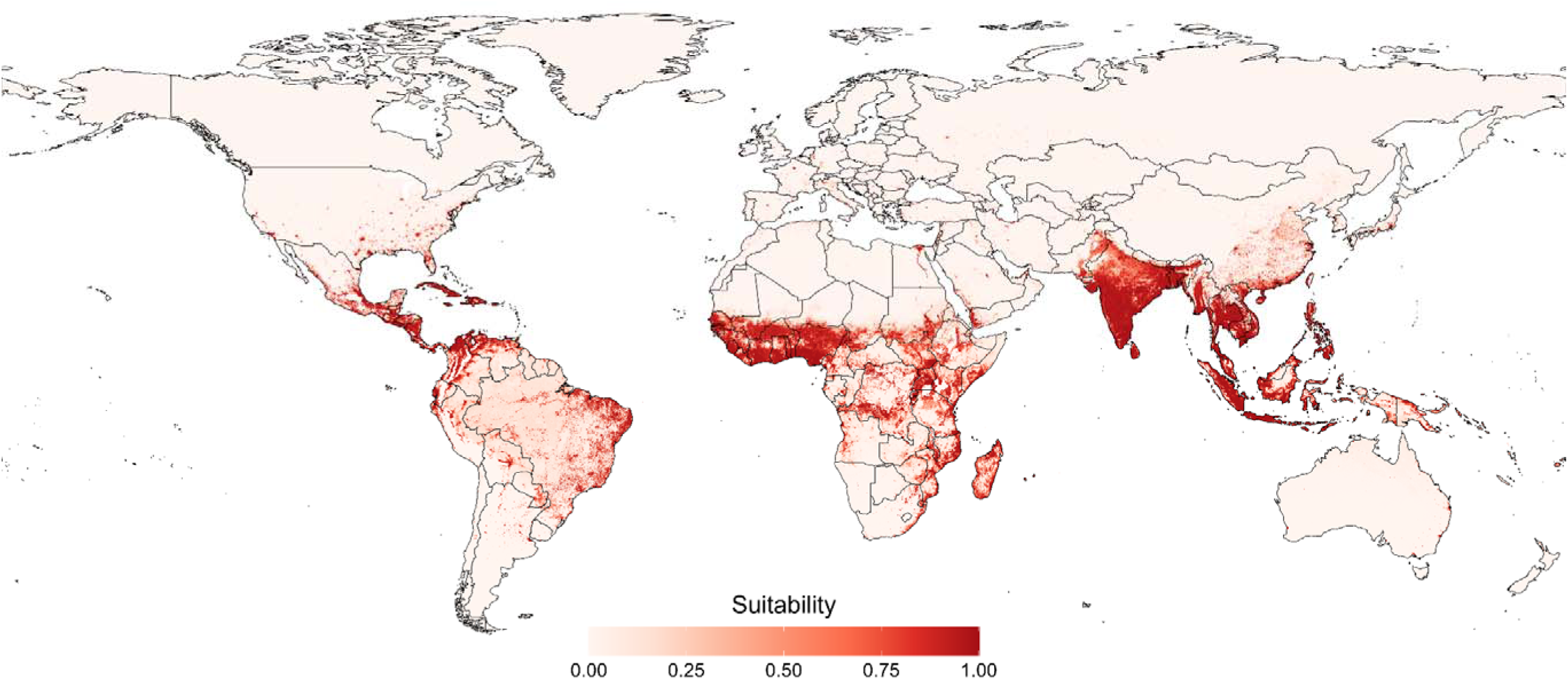
Global mean predicted suitability for chikungunya masked by temperature suitability of vector.

Over the full model horizon, from 2025 to 2050, there were a cumulative 737 million (708 million–770 million) CHIKV infections in the 31 countries and Venezuela. In the absence of vaccination, these infections led to substantial cases of symptomatic chikungunya, hospitalisation, death and chronic chikungunya, resulting in a cumulative 5.96 million (5.10 million–6.93 million) DALYs. (**Table 1**) Chronic chikungunya caused the greatest share of chikungunya DALYs, followed by death. (**Figure 2A-2B, Table S3**)

**Figure 2.**
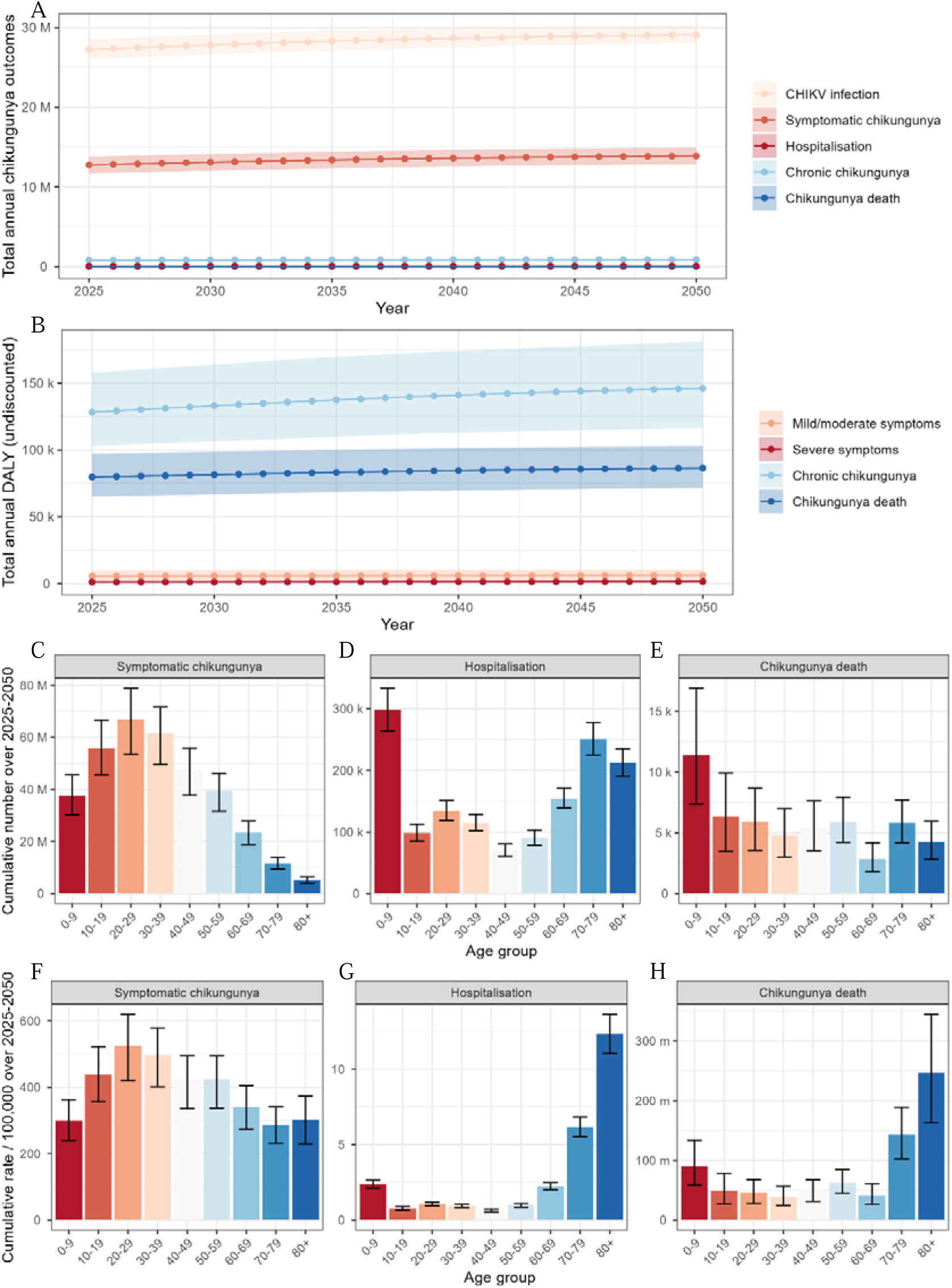
Projected burden of CHIKV infection from 2025 to 2050 across 31 countries and Venezuela in the absence of vaccination and associations with age. (**A**) Change over time in the annual number of CHIKV infection outcomes. (**B**) Change over time in annual DALYs associated with chikungunya. (**C**) The cumulative total number of symptomatic chikungunya cases, stratified by age group. (**D**) The cumulative total number of chikungunya hospitalisations (severe symptoms), stratified by age group. (**E**) The cumulative total number of chikungunya deaths, stratified by age group. (**F**) The cumulative incidence of symptomatic chikungunya per 100,000 person-years, stratified by age group. (**G**) The cumulative incidence of chikungunya hospitalisation (severe symptoms) per 100,000 person-years, stratified by age group. (**H**) The cumulative incidence of chikungunya deaths per 100,000 person-years, stratified by age group. In panels A and B, lines represent means and shading represents 95% uncertainty intervals. In panels C through G, bar heights represent means and error bars represent 95% uncertainty intervals.

**Table 1.** Projected cumulative total health and economic burden of CHIKV infection from 2025 to 2050 across 31 countries and Venezuela in the absence of vaccination. Future costs are discounted annually at 3.5%/year. CHIKV = Alphavirus chikungunya, DALY = disability-adjusted life-year, VSL = value of statistical life, VSLY = value of statistical life-years, I$ = International dollar, K = thousand, M = million, B = billion, T = trillion.

| **Outcome** | **Mean (95% uncertainty interval) cumulative totals** | **Mean (95% uncertainty interval) cumulative totals per 100,000 person-years** |
| --- | --- | --- |
| *Health burden* |  |  |
| CHIKV infection (n) | 737M (708M, 770M) | 881 (845, 920) |
| Symptomatic chikungunya (n) | 349M (322M, 376M) | 417 (384, 450) |
| Hospitalisation (n) | 1.42M (1.31M, 1.53M) | 1.70 (1.57, 1.83) |
| Chronic chikungunya (n) | 21.4M (18.3M, 24.7M) | 25.5 (21.8, 29.4) |
| Death (n) | 52.7K (45.3K, 60.1K) | 0.063 (0.054, 0.072) |
| DALYs (n) | 5.96M (5.10M, 6.93M) | 7.12 (6.10, 8.28) |
| *Economic burden\** |  |  |
| Healthcare costs (I\$ 2023) | 1.84B (1.69B, 1.99B) | 2.22K (2.03K, 2.40K) |
| Catastrophic healthcare expenditure (n) | 391K (356K, 423K) | 0.471 (0.429, 0.510) |
| Impoverishing healthcare expenditure (n) | 347K (308K, 388K) | 0.419 (0.371, 0.468) |
| Productivity losses (I\$ 2023) | 56.9B (38.6B, 74.8B) | 68.6K (46.6K, 90.2K) |
| Monetised DALYs (I\$ 2023) | 17.8B (14.9B, 21.2B) | 21.4K (17.9K, 25.6K) |
| VSL (I\$ 2023) | 149B (129B, 169B) | 180K (155K, 204K) |
| VSLY (I\$ 2023) | 159B (132B, 191B) | 192K (159K, 230K) |
| Societal costs (I\$ 2023) | 76.5B (57.4B, 95.7B) | 92.2K (69.2K, 115K) |

\* Venezuela was excluded from the analyses for the economic burden because there are no reliable estimates of economic parameters available in the country due to its recent economic collapse. Societal costs are the sum of healthcare costs, productivity losses and monetised DALYs.

The projected burden of chikungunya varied greatly across age groups. (**Figure 2C-2H**) Annual symptomatic chikungunya per 100,000 population was lowest in children aged 0-9 [299 (240-363)] and those older than 70 [286 (231-341) to 302 (230-373)], and highest in people aged 20-29 [525 (420-620)]. In terms of hospitalisation and death, adults aged 70+ had by far the greatest incidence. (**Table S4**)

The cumulative societal costs of chikungunya were estimated at $76.5 billion (57.4 billion-95.7 billion). Productivity loss was the main driver of societal costs, followed by monetised DALYs, and, lastly, healthcare expenditure. (**Table 1**) Acute mild/moderate chikungunya was the main driver of healthcare expenditure and productivity losses, whereas chronic chikungunya made the largest contributions to monetised DALYs. (**Table S5**) Estimated DALYs per 100,000 person-years ranged from 4.22 (3.50-5.05) in Bolivia to 9.49 (7.90-11.3) in Costa Rica, whereas estimated societal costs per 100,000 person-years ranged from $9,870 ($7,210-$12,500) in Chad to $454,000 ($347,000-$571,000) in Panama. (**Table S6**)

### Vaccine impact

The benefits of a population-wide vaccination campaign targeting individuals aged 12+ over 5 years depended on the share of the population vaccinated annually. Under base-case assumptions of 70% vaccine efficacy and a vaccine-induced immune response lasting 10 years, campaigns targeting 2% and 10% annual coverage, respectively, required 236 million and 1.18 billion doses (Venezuela accounting for about 1%), and averted 105,000 (87,700-124,000) and 523,000 (439,000-619,000) DALYs, as well as $2.16 billion ($1.58 billion-$2.74 billion) and $10.8 billion ($7.90 billion-$13.7 billion) in societal costs across the 31 countries. A routine campaign targeting 65% of children on their 12^th^ birthday over a period of 16 years required a similar number of vaccine doses (511 million) as the population-wide campaign targeting 4% annual coverage (473 million doses), but averted less burden, including 232,000 (174,000-296,000) DALYs and $1.65 billion ($1.24 billion-$2.10 billion) in societal costs. Combined campaigns required the most doses and averted the greatest share of health and economic outcomes. The most ambitious combined campaign targeting the greatest annual coverage during the initial population-wide phase (10%) led to the greatest health and economic benefits, averting 681,000 (569,000-802,000) DALYs and $11.8 billion ($8.73 billion-$14.9 billion) in societal costs. (**Table 2**, **Table S1**)

**Table 2.** Projected cumulative total health outcomes and economic costs averted due to CHIKV vaccination from 2025 to 2050 across 31 countries and Venezuela, depending on the vaccination campaign. Vaccine was base-case assumed 70% efficacy and future costs are discounted annually at 3.5%/year. DALY = disability-adjusted life-year, VSL = value of statistical life, VSLY = value of statistical life-years, I$ = International dollar, K = thousand, M = million, B = billion.

|  | Cumulative totals (95%UI) by different vaccination campaign strategies |  |  |  |  |  |  |
| --- | --- | --- | --- | --- | --- | --- | --- |
| Type | Population wide |  |  | Routine | Population-wide + Routine (P+R) |  |  |
| Age | 12-99 |  |  | 12 | P: 12-99; R: 12 |  |  |
| Vaccination period | 2025-2029 |  |  | 2025-2040 | P: 2025-2029; R: 2030-2040 |  |  |
| Annual target coverage^ | 2% | 4% | 10% | 65% | P: 2%; R: 65% | P: 4%; R: 65% | P: 10%; R: 65% |
| **Health outcomes averted** |  |  |  |  |  |  |  |
| Symptomatic chikungunya (n) | 6.85M (6.26M, 7.38M) | 13.7M (12.5M, 14.8M) | 34.3M (31.3M, 36.9M) | 15.4M (13.0M, 17.8M) | 17.3M (15.3M, 19.3M) | 24.2M (21.8M, 26.5M) | 44.7M (40.8M, 48.6M) |
| Hospitalisation (n) | 24.7K (22.8K, 26.5K) | 49.4K (45.6K, 53.0K) | 124K (114K, 132K) | 28.0K (24.6K, 31.6K) | 43.7K (39.9K, 47.4K) | 68.5K (62.9K, 73.8K) | 143K (131K, 153K) |
| Chronic chikungunya (n) | 419K (356K, 482K) | 839K (712K, 965K) | 2.10M (1.78M, 2.41M) | 941K (760K, 1.15M) | 1.06M (884K, 1.26M) | 1.48M (1.25M, 1.73M) | 2.74M (2.32M, 3.18M) |
| Chikungunya death (n) | 890 (761, 1.02K) | 1.78K (1.52K, 2.03K) | 4.45K (3.80K, 5.08K) | 1.66K (1.05K, 2.41K) | 2.02K (1.56K, 2.61K) | 2.91K (2.33K, 3.56K) | 5.58K (4.67K, 6.50K) |
| DALYs (n) | 105K (87.7K, 124K) | 209K (175K, 247K) | 523K (439K, 619K) | 232K (174K, 296K) | 262K (214K, 317K) | 367K (304K, 435K) | 681K (569K, 802K) |
| **Economic costs averted\*** |  |  |  |  |  |  |  |
| Healthcare costs (I\$ 2023) | 45.5M (41.2M, 49.7M) | 91.0M (82.5M, 99.3M) | 228M (206M, 248M) | 64.7M (54.9M, 74.5M) | 84.8M (76.7M, 94.2M) | 130M (118M, 143M) | 267M (242M, 292M) |
| Catastrophic healthcare expenditure (n) | 8.92K (8.08K, 9.72K) | 17.8K (16.2K, 19.4K) | 44.6K (40.4K, 48.6K) | 10.4K (9.03K, 11.8K) | 16.3K (14.7K, 17.8K) | 25.2K (22.9K, 27.4K) | 52.0K (47.3K, 56.5K) |
| Impoverishing healthcare expenditure (n) | 8.43K (7.45K, 9.44K) | 16.9K (14.9K, 18.9K) | 42.1K (37.2K, 47.2K) | 7.12K (6.00K, 8.34K) | 13.3K (11.7K, 14.9K) | 21.7K (19.1K, 24.2K) | 47.0K (41.5K, 52.4K) |
| Productivity losses (I\$ 2023) | 1.66B (1.11B, 2.20B) | 3.32B (2.22B, 4.41B) | 8.31B (5.56B, 11.0B) | 1.08B (731M, 1.47B) | 2.32B (1.58B, 3.07B) | 3.98B (2.70B, 5.26B) | 8.97B (6.02B, 11.9B) |
| Monetised DALYs (I\$ 2023) | 456M (374M, 549M) | 911M (749M, 1.10B) | 2.28B (1.87B, 2.75B) | 510M (387M, 644M) | 765M (632M, 914M) | 1.22B (1.01B, 1.45B) | 2.59B (2.15B, 3.10B) |
| VSL losses (I\$ 2023) | 3.24B (2.77B, 3.68B) | 6.47B (5.55B, 7.36B) | 16.2B (13.9B, 18.4B) | 4.39B (2.76B, 6.42B) | 5.90B (4.69B, 7.41B) | 9.14B (7.54B, 11.0B) | 18.8B (16.0B, 21.7B) |
| Type | Population wide |  |  | Routine | Population-wide + Routine (P+R) |  |  |
| Age | 12-99 |  |  | 12 | P: 12-99; R: 12 |  |  |
| Vaccination period | 2025-2029 |  |  | 2025-2040 | P: 2025-2029; R: 2030-2040 |  |  |
| Annual target coverage^ | 2% | 4% | 10% | 65% | P: 2%; R: 65% | P: 4%; R: 65% | P: 10%; R: 65% |
| VSLY losses (I\$ 2023) | 2.67B (2.21B, 3.14B) | 5.34B (4.42B, 6.28B) | 13.3B (11.1B, 15.7B) | 7.08B (4.39B, 10.4B) | 6.96B (5.11B, 9.35B) | 9.63B (7.38B, 12.4B) | 17.6B (14.2B, 21.6B) |
| Societal costs (I\$ 2023) | 2.16B (1.58B, 2.74B) | 4.33B (3.16B, 5.48B) | 10.8B (7.90B, 13.7B) | 1.65B (1.24B, 2.10B) | 3.17B (2.35B, 3.98B) | 5.33B (3.96B, 6.71B) | 11.8B (8.73B, 14.9B) |

^ Actual achieved coverage = 95% of target coverage, accounting for 5% wastage. \*Venezuela was excluded from the analyses because there is no reliable estimate of economic parameter available in the country due to its economic collapse in the recent years. Societal costs are the sum of healthcare costs, productivity losses and monetised DALYs.

The cost-effectiveness of vaccination campaigns was measured using the threshold vaccination cost, i.e. the cumulative health-economic benefits accrued per dose of vaccine (for a two-dose vaccine, the estimate should be halved) calculated across all 31 countries. Population-wide campaigns were the most cost-effective, with a dose-weighted mean threshold vaccination cost of $9.90 ($7.23-$12.5), while routine campaigns were least cost-effective, with a threshold vaccination cost of $4.16 ($3.12-$5.30). Population-wide campaigns targeting different annual coverages had the same threshold vaccination costs, as per-dose vaccine impacts scaled linearly with vaccine coverage in the absence of an impact on transmission. Combined campaigns had intermediate cost-effectiveness, with threshold vaccination costs ranging from $6.81 ($5.05-$8.55) to $8.83 ($6.52-$11.1), respectively, under assumptions of 2% and 10% annual target coverage during the population-wide phase of the campaign. (**Table 3**)

**Table 3.**
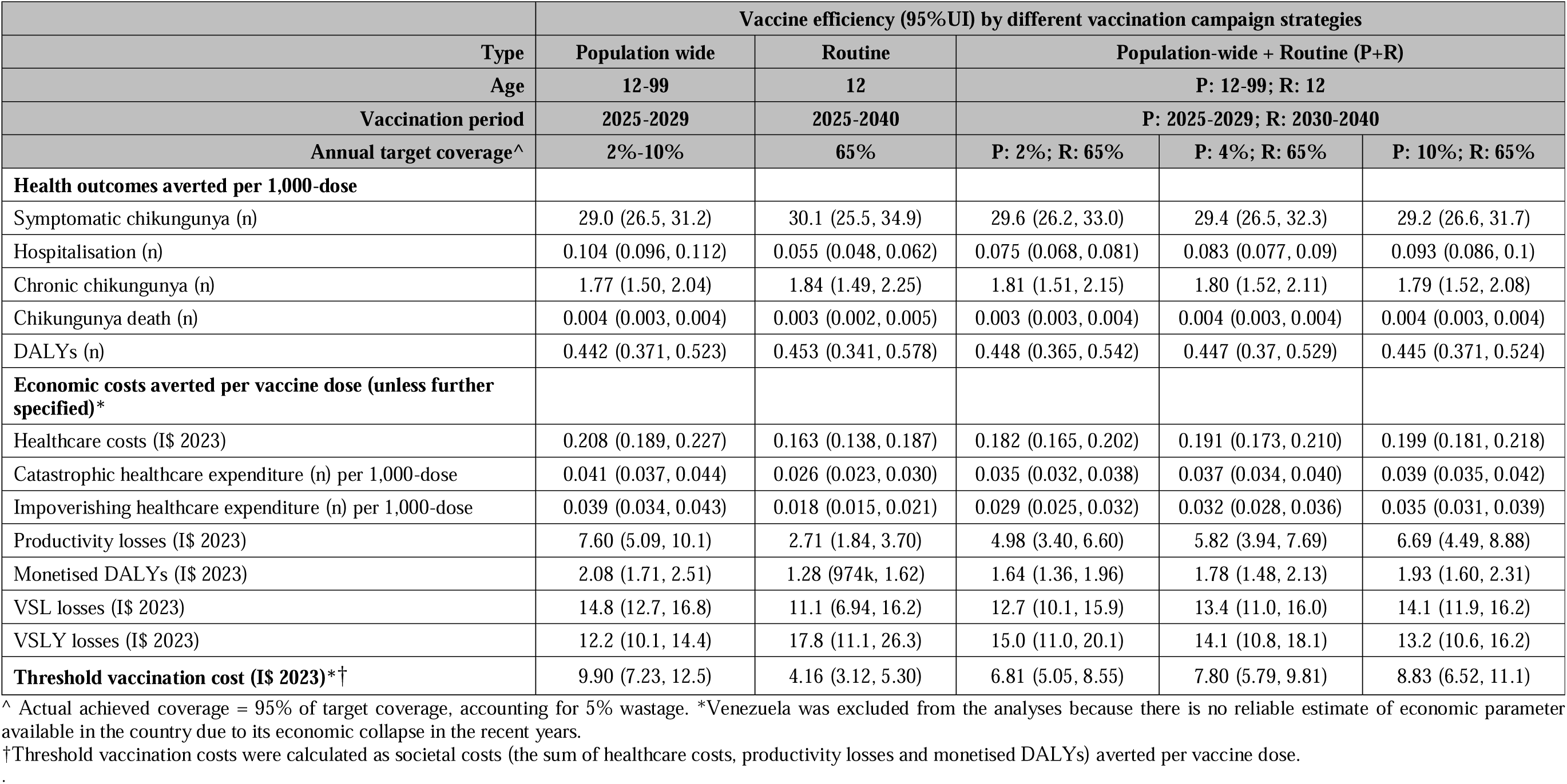
Efficiency in averting health outcomes, economic costs and threshold vaccination costs under different vaccination campaign scenarios. Base-case assumptions include 70% vaccine efficacy against disease and an annual 3.5% discount rate for future costs. DALY = disability-adjusted life-year, VSL = value of statistical life, VSLY = value of statistical life-years, I$ = International dollar.

Vaccine impacts and cost-effectiveness were also measured at the individual country level. (**Figure 3, Figure S4**) Threshold vaccination costs for population-wide campaigns ranged from $1.11 ($0.77- $1.44) in Democratic Republic of Congo to $48.1 ($36.0-$60.3) in Panama, and for routine campaigns from $0.57 ($0.41-$0.74) to $21.6 ($16.2-$27.6) in these same countries. (**Figure 3**) Threshold vaccination costs for population wide-campaigns were above $20 and $40, respectively, in 11 countries (35.5%) and 1 country (3.2%), whereas those for routine campaigns were above $20 only in Panama. (**Table S7**)

**Figure 3.**
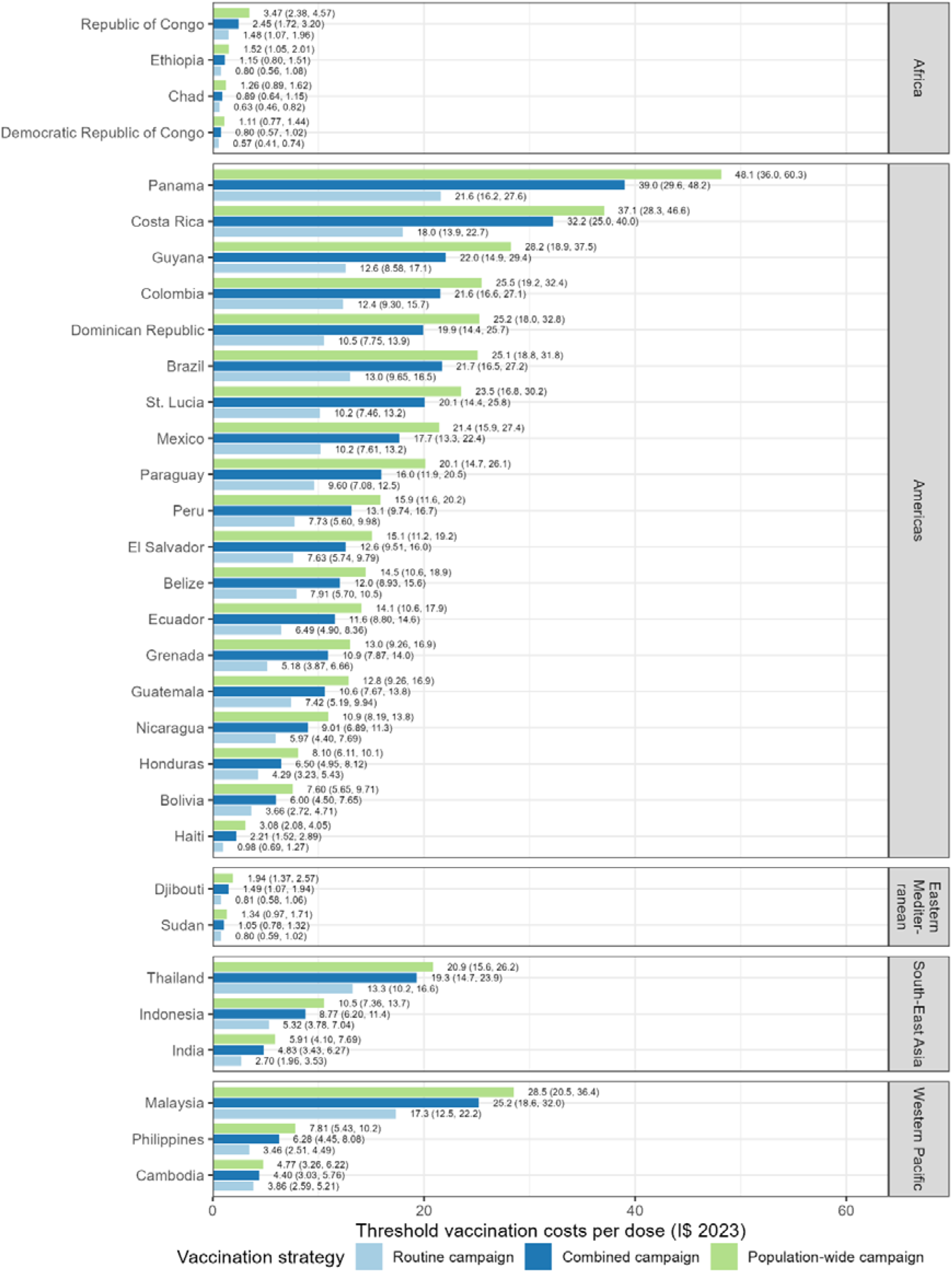
Threshold vaccination costs by different vaccination strategies in each of the 31 countries analysed. Combined campaign analysed starts with the population-wide campaign over 5 years targeting 20% coverage in total (actual achieved coverage = 95% of target coverage accounting for 5% wastage). Venezuela was excluded from health-economic analyses due to a lack of reliable estimates of economic parameters due to its economic collapse in the recent years. Future costs and life-years are discounted annually at 3.5%/year. I$ = International dollar.

In the base-case analysis, vaccine-associated adverse events resulted in DALYs and societal costs that were both equal to less than 1% of the DALYs and societal costs averted through vaccination. However, in the scenario analysis considering a broader vaccine safety profile as observed in randomised trials and post-marketing surveillance for VLA1553, including mild, moderate, and severe myalgia/arthralgia and chronic symptoms but still excluding vaccine-induced mortality, vaccine-associated adverse events led to DALY equivalent to approximately 70% of the DALY averted through chikungunya prevention across all vaccination strategies. In addition, the societal costs attributable to vaccine-associated adverse events slightly exceeded the costs averted from disease prevention under the population-wide vaccination strategy, while remaining slightly lower than the costs averted under strategies that included routine vaccination. (**Table 4**)

**Table 4.**
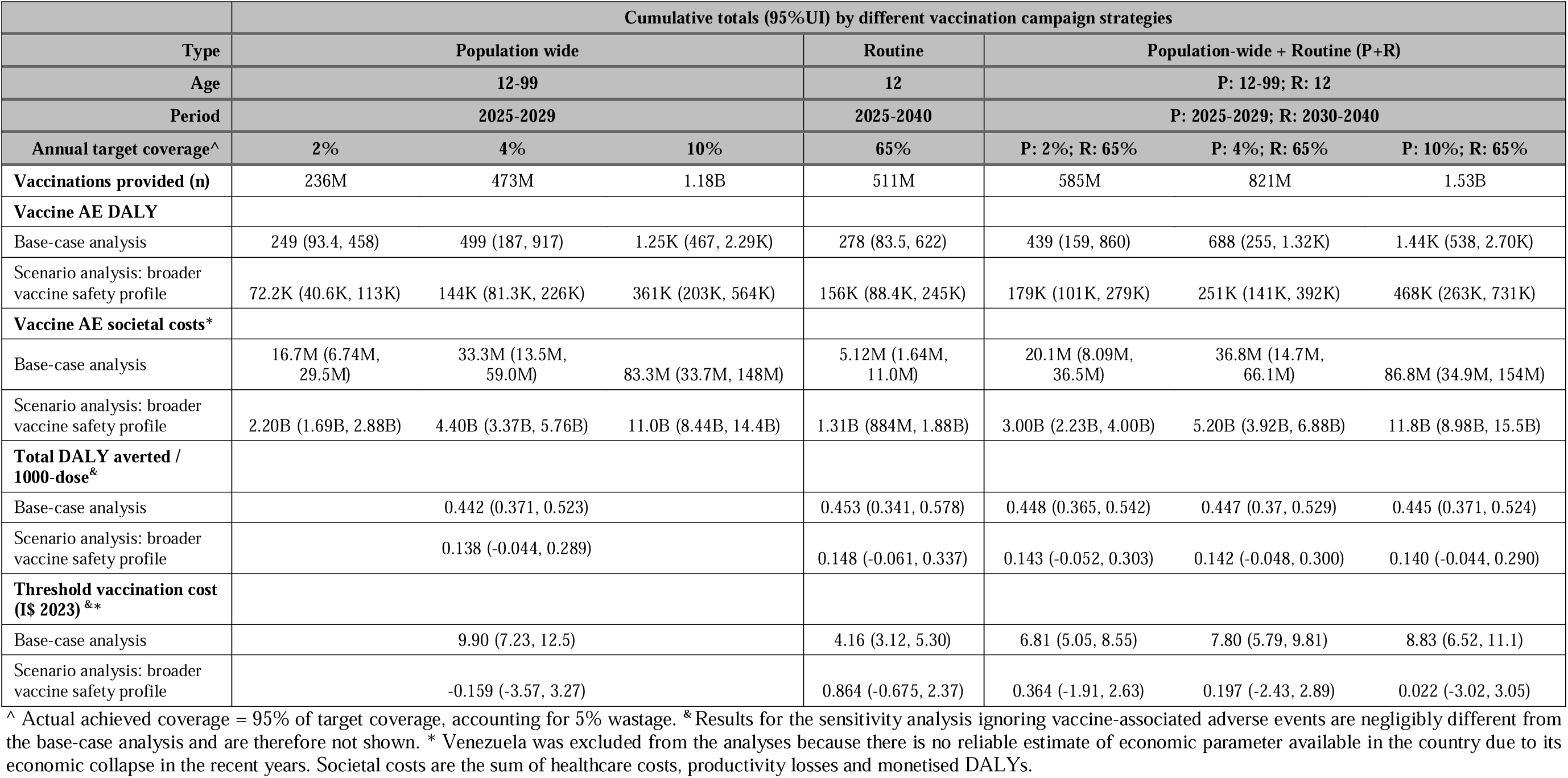
Projected cumulative DALY and societal costs associated with vaccine AE and resulting overall efficiency in total DALY averted and threshold vaccination costs from 2025 to 2050 across 31 countries and Venezuela, depending on the vaccination campaign. Vaccine was base-case assumed 70% efficacy and future costs are discounted annually at 3.5%/year. DALY = disability-adjusted life-year, AE = adverse event, I$ = International dollar, K = thousand, M = million, B = billion.

Sensitivity analyses varying vaccine efficacy, parameter inputs and model assumptions had important impacts on projected vaccine benefits and thus threshold vaccination cost. (**Figure S5, Table S8-S13**) The sensitivity analysis using an estimate of symptom probability from the Philippines (0.180) instead of Nicaragua (0.505) led to a roughly 3-fold reduction in estimated burden, vaccine impact and threshold vaccination costs. The sensitivity analysis assuming undetected acute mild/moderate symptomatic infections share the same disability weight and probability of developing chronic symptoms as detected acute mild/moderate symptomatic infections led to an approximately 6-fold increase in DALYs and a 4-fold increase in societal costs. However, the order of vaccination campaigns in terms of threshold vaccination cost was robust to sensitivity analyses, with population-wide campaigns always leading to greatest per-dose benefits. (**Table S12-S13**)

## DISCUSSION

This study projected the health-economic burden of chikungunya across 31 countries, mostly LMICs, from 2025-2050, and estimated the health-economic benefits of different preventive vaccine rollout strategies. We estimated that CHIKV could cause 737 million infections, 52,700 deaths and 5.96 million DALYs over this period, generating $76.5 billion in societal costs. A combined vaccination strategy of an initial population-wide campaign followed by annual routine adolescent vaccination had the greatest impact, averting a cumulative estimated 5,580 deaths, 681,000 DALYs and $11.8 billion in societal costs. However, the population-wide campaign alone was the most cost-effective strategy, with threshold vaccination costs ranging from $1.11 ($0.77-$1.44) per dose in Democratic Republic of Congo to $48.1 ($36.0-$60.3) per dose in Panama, assuming a single-dose vaccine with a relatively favourable safety profile and a ten-year immunological response. Threshold vaccination costs exceeded $20 in 11 of the 31 countries, a price comparable to the per-dose costs of human papillomavirus vaccination in Latin American LMICs through the Revolving Fund, including an estimated $5 for vaccine delivery^40,41^. These findings suggest that, in a substantial proportion of endemic settings, preventive CHIKV vaccination could remain cost-effective at prices comparable to existing routinely delivered vaccines.

The health burden of chikungunya was predicted to vary substantially across the 31 included countries (range of means per 100,000 person-years: 527 to 1,080 CHIKV infections; 4.22 to 9.49 DALYs). This heterogeneity is explained by the inclusion of country-specific estimates of population demographics, vector and CHIKV suitability, seroprevalence and FoI. Further accounting for economic heterogeneity, including country-specific treatment costs, labour force participation and wages, as well as cost-effectiveness thresholds derived from country-specific healthcare spending opportunity costs, led to greater heterogeneity in the economic burden associated with CHIKV, ranging across included countries from $9,870 to $454,000 in cumulative societal costs per 100,000 person-years. This heterogeneity underlies variability in country-level cost-effectiveness predictions. The economic burden of CHIKV infection was driven primarily by productivity losses, and adult-targeted population-wide campaigns being more cost-effective than adolescent-targeted routine campaigns reflects both greater average risks of disease and greater labour force participation in adults relative to adolescents.

Our vaccine impact projections rely on vaccine efficacy assumptions that reflect a hypothetical vaccine due to limited real-world data. While recent phase 3 trials for VLA1553 and PXVX0317^7,8^ report high immunogenicity, including up to 4 years in the case of VLA1553^30^, the durability of protection, effect on clinical disease, and impact on transmission remain uncertain. Our results may therefore underestimate the benefits of vaccination, particularly if vaccines reduce transmission or provide longer-lasting immunity. In addition, for simplicity we characterised CHIKV transmission using long-term average FoI predictions that assume the same annual rate of infection over time. However, CHIKV transmission is characterised in many regions by sporadic epidemics with high attack rates. Accounting for time-varying FoI would likely have resulted in similar mean incidence estimates given our model’s long time horizon, but would further capture the temporal clustering of infections, which could influence vaccine impact given the assumed duration of protection of 10 years and discounting of costs. Future cost-effectiveness analyses should incorporate real-world data on the duration of vaccine effectiveness against disease and transmission, once such data become available, and use these to account for outbreak dynamics in epidemic settings. Future analyses should also consider alternative vaccination strategies, including reactive vaccination during outbreaks and targeted vaccination of subgroups in whom the balance of benefits, risks and costs is expected to be the most favourable.

These modelling results are sensitive to certain assumptions and uncertainties. There is recognised concern that chikungunya severity and mortality risk may be underestimated^42,43^, and the evolution of new lineages or emergence of existing lineages in areas with low pre-existing immunity could result in outbreaks that are more severe than those characterised by our model^44^. Assumptions regarding symptom probability and disease risk in detected (medically attended) versus undetected symptomatic infections had the largest impact on our results. However, despite variation in total cumulative chikungunya burden and vaccine impact across population groups in these sensitivity analyses, the relative efficiency of included vaccine strategies was consistent, with population-wide campaigns always leading to the greatest cost-effectiveness. Further, in a sensitivity analysis using an alternative CHIKV suitability surface from the literature, estimates of annual FoI at the country level were consistent, resulting in only a 3% increase in total infections over the study period.

Our health burden projections are comparable to a recent model-based analysis of chikungunya’s global burden by dos Santos *et al*^12^, which estimated fewer average annual infections (20.5 million) than our study (28.4 million) across the 31 countries included in both studies (theirs excluded Grenada), although their estimates did not take future projected population growth into account and treated outbreak probability and size as stochastic events, independent of underlying country characteristics such as CHIKV suitability. In turn, they estimated fewer annual symptomatic infections (10.2 million vs. 13.4 million), and infections resulting chronic symptoms (491,000 vs. 822,000), but a similar number of deaths (2,460 vs. 2,030), leading to overall lower annual DALY burden (162,000 vs. 229,000).

Importantly, our study provides the first estimates of chikungunya’s economic burden and the cost-effectiveness of vaccination across multiple settings. Our estimates of the health-economic burden of chikungunya and benefits of vaccination may inform decision-making regarding the rollout of both currently licensed and forthcoming CHIKV vaccines. Estimates of vaccine impact in our analysis varied greatly across included countries, suggesting that the returns-on-investment and therefore health-economic prioritisation of CHIKV vaccination is likely to be highly context specific. These findings should be interpreted in light of ongoing vaccine policy and regulatory discussions. Recent regulatory decisions, including the temporary suspension of VLA1553 use in older age groups due to safety signals, underscore the importance of carefully balancing benefits and risks across target populations^32^. While our base-case analysis incorporates vaccine-related adverse events, these accounted for less than 1% of the overall health-economic benefits of vaccination, supporting a favourable benefit–risk profile in most settings. However, in a scenario analysis incorporating the broader safety profile reported for VLA1553 in randomised trials and post-marketing surveillance vaccine associated adverse events substantially reduced net health and economic benefits of preventive vaccination. This highlights that the cost-effectiveness of preventive CHIKV vaccination is highly sensitive to assumptions regarding vaccine safety.

It is also important to note that these scenarios were based on the safety profile of the live-attenuated vaccine VLA1553 and should not be interpreted as reflecting the safety profile of other CHIKV vaccines, such as the non-replicating virus-like particle vaccine PXVX0317. Our analysis did not explicitly incorporate vaccine-associated mortality or assign disability beyond that included in the reported adverse-event profile. Nevertheless, reports of rare but severe neurological adverse events following VLA1553, including a fatal case in which the vaccine-strain virus was detected in the central nervous system, underscore the importance of continued pharmacovigilance, investigation of host and viral factors predisposing to these events, and optimisation of vaccine safety profiles. Addressing these issues will be critical to maintaining public confidence, informing age-specific recommendations, and maximising the health and economic benefits of future chikungunya vaccination programmes.

The greatest challenge to long-term prospective arbovirus burden modelling is fundamental uncertainty regarding epidemic risk. We made conservative assumptions throughout our modelling pipeline to avoid overestimating future burden and have not taken into account future climate and environmental change affecting CHIKV suitability, leading to potential underestimation of future burden in several, but not all, considered countries^45^. We also excluded countries reporting no evidence of substantial outbreaks up to 2022. We assumed that natural CHIKV infection confers lifelong immune protection against reinfection, while vaccination only protects against disease for 10 years, therefore representing a cautious scenario for both infection burden estimation and vaccine impact. Finally, we did not consider the emergence of novel variants with vaccine-escape properties.

In conclusion, this study has provided detailed projections of the health-economic burden of chikungunya across 31 countries and has estimated the potential returns-on-investment of several proposed strategies for preventive CHIKV vaccination. These results may inform investment decisions for current and forthcoming CHIKV vaccines.

## Data Availability

All data used in this study are publicly available. The code and minimum dataset required to reproduce results are available at https://github.com/zhoujunwen/HE_MODELS_CHIK_VacDisease.

## Acknowledgements

This work was conducted by the OxLiv Consortium and funded by the Coalition for Epidemic Preparedness Innovations (CEPI) through Vaccine Impact Assessment project funding. We acknowledge the CEPI project team (Project Lead: Christinah Mukandavire) for continuous support and the project’s external advisory team for their invaluable feedback. KBP and DRMS are supported by the Medical Research Foundation (MRF-160-0017-ELP-POUW-C0909). TDH thanks the Li Ka Shing Foundation for institutional funding. The views expressed are those of the authors and not necessarily those of the institutions with which they are affiliated.

## Contributions

KBP, JeL and TDH acquired funding and supervised the work. NS, DRMS, HSR, JZ, KMH and LPGC reviewed the literature and synthesised data. DRMS, NS, KBP, AAT, MLTS and KAD developed the vaccine administration strategies. JM, WW, JL, JeL, JZ, and KBP conducted CHIKV suitability modelling. JZ, HSR and KBP conducted chikungunya and force of infection mapping. JZ, NS and DRMS developed all other model components, conducted the health economic analyses and produced results. JZ, DRMS, KBP and TDH interpreted results. The underlying data were verified by JZ, DRMS, KBP and TDH, and all authors had full access to the study data and accept responsibility to submit for publication. JZ wrote the first draft with supervision from DRMS and KBP. The final version of this manuscript was reviewed and approved by all authors.

