## Supplementary methods for "Health-economic burden of chikungunya infection and potential cost-effectiveness of preventive vaccination in 31 countries"

### SUPPLEMENTARY MATERIALS

### 1. SUPPLEMENTARY TABLES AND FIGURES

***Figure S1. Model structure showing the clinical progression of individuals with chikungunya virus (Alphavirus chikungunya, CHIKV) infection through different health states.***

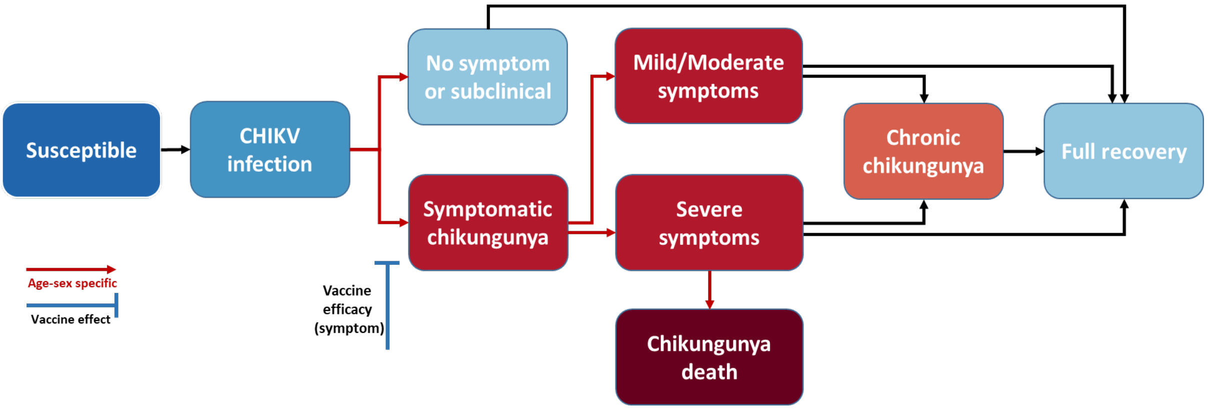

Healthcare use and economic outcomes associated with each health state are not shown. Red arrows show model transition probabilities that vary depending on individuals' age and/or sex. See methods and supplementary appendix for technical details regarding model structure, parameterisation, simulation and outcome calculation.

***Figure S2. Example of three key types of vaccination campaign over 2025-2040***

**A**. Population-wide vaccination (age 12-99, annual coverage 10%, period 2025-2029)

| Age, years \ Year | 2025 | 2026 | 2027 | 2028 | 2029 | 2030 | 2031 | … | 2040 |
|---|---|---|---|---|---|---|---|---|---|
| 99 | 10% | 20% | 30% | 40% | 50% | 50% | 50% | 50% | 50% |
| … | 10% | 20% | 30% | 40% | 50% | … | … | … | … |
| 16 | 10% | 20% | 30% | 40% | 50% | 40% | 30% | … | 0 |
| 15 | 10% | 20% | 30% | 40% | 40% | 30% | 20% | … | 0 |
| 14 | 10% | 20% | 30% | 30% | 30% | 20% | 10% | 0 | 0 |
| 13 | 10% | 20% | 20% | 20% | 20% | 10% | 0 | 0 | 0 |
| 12 | 10% | 10% | 10% | 10% | 10% | 0 | 0 | 0 | 0 |
| … | 0 | 0 | 0 | 0 | 0 | 0 | 0 | 0 | 0 |
| 0 | 0 | 0 | 0 | 0 | 0 | 0 | 0 | 0 | 0 |

**B**. Routine vaccination (age 12, annual coverage 65%, period 2025-2040)

| Age, years \ Year | 2025 | 2026 | 2027 | 2028 | 2029 | 2030 | 2031 | … | 2040 |
|---|---|---|---|---|---|---|---|---|---|
| 99 | 0 | 0 | 0 | 0 | 0 | 0 | 0 | 0 | 0 |
| … | 0 | 0 | 0 | 0 | 0 | … | … | … | … |
| 16 | 0 | 0 | 0 | 0 | 65% | 65% | 65% | 65% | 65% |
| 15 | 0 | 0 | 0 | 65% | 65% | 65% | 65% | 65% | 65% |
| 14 | 0 | 0 | 65% | 65% | 65% | 65% | 65% | 65% | 65% |
| 13 | 0 | 65% | 65% | 65% | 65% | 65% | 65% | 65% | 65% |
| 12 | 65% | 65% | 65% | 65% | 65% | 65% | 65% | 65% | 65% |
| … | 0 | 0 | 0 | 0 | 0 | 0 | 0 | 0 | 0 |
| 0 | 0 | 0 | 0 | 0 | 0 | 0 | 0 | 0 | 0 |

**C**. Population-wide vaccination (A) over 2025-2029; routine vaccination (B) over 2030-2040

| Age, years \ Year | 2025 | 2026 | 2027 | 2028 | 2029 | 2030 | 2031 | … | 2040 |
|---|---|---|---|---|---|---|---|---|---|
| 99 | 10% | 20% | 30% | 40% | 50% | 50% | 50% | 50% | 50% |
| … | 10% | 20% | 30% | 40% | 50% | … | … | … | … |
| 16 | 10% | 20% | 30% | 40% | 50% | 40% | 30% | … | 65% |
| 15 | 10% | 20% | 30% | 40% | 40% | 30% | 20% | … | 65% |
| 14 | 10% | 20% | 30% | 30% | 30% | 20% | 10% | 65% | 65% |
| 13 | 10% | 20% | 20% | 20% | 20% | 10% | 65% | 65% | 65% |
| 12 | 10% | 10% | 10% | 10% | 10% | 65% | 65% | 65% | 65% |
| … | 0 | 0 | 0 | 0 | 0 | 0 | 0 | 0 | 0 |
| 0 | 0 | 0 | 0 | 0 | 0 | 0 | 0 | 0 | 0 |

The Numbers in each cell refer to the percentage of all individuals of the corresponding age (y-axis) that have previously been vaccinated (not including wastage) by year's end.

***Figure S3. Example of proportion of population protected by the vaccine with 10 years protection over 2025-2050 in three types of vaccination campaign.***

**A**. Population-wide vaccination (age 12-99, annual coverage 10%, period 2025-2029)

| Age, years | 2025 | 2026 | 2027 | 2028 | 2029 | 2030 | 2031 | 2032 | 2033 | 2034 | 2035 | 2036 | 2037 | 2038 | 2039 | 2040 | 2041 | 2042 | … | 2048 | 2049 | 2050 |
|---|---|---|---|---|---|---|---|---|---|---|---|---|---|---|---|---|---|---|---|---|---|---|
| 23-99 | 10% | 20% | 30% | 40% | 50% | 50% | 50% | 50% | 50% | 50% | 40% | 30% | 20% | 10% | 0 | 0 | 0 | 0 | 0 | 0 | 0 | 0 |
| 22 | 10% | 20% | 30% | 40% | 50% | 50% | 50% | 50% | 50% | 50% | 40% | 30% | 20% | 10% | 0 | 0 | 0 | 0 | 0 | 0 | 0 | 0 |
| 21 | 10% | 20% | 30% | 40% | 50% | 50% | 50% | 50% | 50% | 50% | 40% | 30% | 20% | 10% | 0 | 0 | 0 | 0 | 0 | 0 | 0 | 0 |
| 20 | 10% | 20% | 30% | 40% | 50% | 50% | 50% | 50% | 50% | 40% | 30% | 20% | 10% | 0 | 0 | 0 | 0 | 0 | 0 | 0 | 0 | 0 |
| 19 | 10% | 20% | 30% | 40% | 50% | 50% | 50% | 50% | 40% | 30% | 20% | 10% | 0 | 0 | 0 | 0 | 0 | 0 | 0 | 0 | 0 | 0 |
| 18 | 10% | 20% | 30% | 40% | 50% | 50% | 50% | 40% | 30% | 20% | 10% | 0 | 0 | 0 | 0 | 0 | 0 | 0 | 0 | 0 | 0 | 0 |
| 17 | 10% | 20% | 30% | 40% | 50% | 50% | 40% | 30% | 20% | 10% | 0 | 0 | 0 | 0 | 0 | 0 | 0 | 0 | 0 | 0 | 0 | 0 |
| 16 | 10% | 20% | 30% | 40% | 50% | 40% | 30% | 20% | 10% | 0 | 0 | 0 | 0 | 0 | 0 | 0 | 0 | 0 | 0 | 0 | 0 | 0 |
| 15 | 10% | 20% | 30% | 40% | 40% | 30% | 20% | 10% | 0 | 0 | 0 | 0 | 0 | 0 | 0 | 0 | 0 | 0 | 0 | 0 | 0 | 0 |
| 14 | 10% | 20% | 30% | 30% | 30% | 20% | 10% | 0 | 0 | 0 | 0 | 0 | 0 | 0 | 0 | 0 | 0 | 0 | 0 | 0 | 0 | 0 |
| 13 | 10% | 20% | 20% | 20% | 20% | 10% | 0 | 0 | 0 | 0 | 0 | 0 | 0 | 0 | 0 | 0 | 0 | 0 | 0 | 0 | 0 | 0 |
| 12 | 10% | 10% | 10% | 10% | 10% | 0 | 0 | 0 | 0 | 0 | 0 | 0 | 0 | 0 | 0 | 0 | 0 | 0 | 0 | 0 | 0 | 0 |
| … | 0 | 0 | 0 | 0 | 0 | 0 | 0 | 0 | 0 | 0 | 0 | 0 | 0 | 0 | 0 | 0 | 0 | 0 | 0 | 0 | 0 | 0 |
| 0 | 0 | 0 | 0 | 0 | 0 | 0 | 0 | 0 | 0 | 0 | 0 | 0 | 0 | 0 | 0 | 0 | 0 | 0 | 0 | 0 | 0 | 0 |

Year

**B**. Routine vaccination (age 12, annual coverage 65%, period 2025-2040)

| Age, years | 2025 | 2026 | 2027 | 2028 | 2029 | 2030 | 2031 | 2032 | 2033 | 2034 | 2035 | 2036 | 2037 | 2038 | 2039 | 2040 | 2041 | 2042 | … | 2048 | 2049 | 2050 |
|---|---|---|---|---|---|---|---|---|---|---|---|---|---|---|---|---|---|---|---|---|---|---|
| 23-99 | 0 | 0 | 0 | 0 | 0 | 0 | 0 | 0 | 0 | 0 | 0 | 0 | 0 | 0 | 0 | 0 | 0 | 0 | 0 | 0 | 0 | 0 |
| 22 | 0 | 0 | 0 | 0 | 0 | 0 | 0 | 0 | 0 | 0 | 0 | 0 | 0 | 0 | 0 | 0 | 0 | 0 | 0 | 0 | 0 | 0 |
| 21 | 0 | 0 | 0 | 0 | 0 | 0 | 0 | 0 | 0 | 65% | 65% | 65% | 65% | 65% | 65% | 65% | 65% | 65% | 65% | 65% | 65% | 0 |
| 20 | 0 | 0 | 0 | 0 | 0 | 0 | 0 | 0 | 65% | 65% | 65% | 65% | 65% | 65% | 65% | 65% | 65% | 65% | 65% | 65% | 0 | 0 |
| 19 | 0 | 0 | 0 | 0 | 0 | 0 | 0 | 65% | 65% | 65% | 65% | 65% | 65% | 65% | 65% | 65% | 65% | 65% | 65% | 0 | 0 | 0 |
| 18 | 0 | 0 | 0 | 0 | 0 | 0 | 65% | 65% | 65% | 65% | 65% | 65% | 65% | 65% | 65% | 65% | 65% | 65% | … | 0 | 0 | 0 |
| 17 | 0 | 0 | 0 | 0 | 0 | 65% | 65% | 65% | 65% | 65% | 65% | 65% | 65% | 65% | 65% | 65% | 65% | 65% | … | 0 | 0 | 0 |
| 16 | 0 | 0 | 0 | 0 | 65% | 65% | 65% | 65% | 65% | 65% | 65% | 65% | 65% | 65% | 65% | 65% | 65% | 65% | … | 0 | 0 | 0 |
| 15 | 0 | 0 | 0 | 65% | 65% | 65% | 65% | 65% | 65% | 65% | 65% | 65% | 65% | 65% | 65% | 65% | 65% | 65% | … | 0 | 0 | 0 |
| 14 | 0 | 0 | 65% | 65% | 65% | 65% | 65% | 65% | 65% | 65% | 65% | 65% | 65% | 65% | 65% | 65% | 65% | 65% | 0 | 0 | 0 | 0 |
| 13 | 0 | 65% | 65% | 65% | 65% | 65% | 65% | 65% | 65% | 65% | 65% | 65% | 65% | 65% | 65% | 65% | 65% | 0 | 0 | 0 | 0 | 0 |
| 12 | 65% | 65% | 65% | 65% | 65% | 65% | 65% | 65% | 65% | 65% | 65% | 65% | 65% | 65% | 65% | 65% | 0 | 0 | 0 | 0 | 0 | 0 |
| … | 0 | 0 | 0 | 0 | 0 | 0 | 0 | 0 | 0 | 0 | 0 | 0 | 0 | 0 | 0 | 0 | 0 | 0 | 0 | 0 | 0 | 0 |
| 0 | 0 | 0 | 0 | 0 | 0 | 0 | 0 | 0 | 0 | 0 | 0 | 0 | 0 | 0 | 0 | 0 | 0 | 0 | 0 | 0 | 0 | 0 |

Year

**C**. Population-wide vaccination (A) over 2025-2029; routine vaccination (B) over 2030-2040

| Age, years | 2025 | 2026 | 2027 | 2028 | 2029 | 2030 | 2031 | 2032 | 2033 | 2034 | 2035 | 2036 | 2037 | 2038 | 2039 | 2040 | 2041 | 2042 | … | 2048 | 2049 | 2050 |
|---|---|---|---|---|---|---|---|---|---|---|---|---|---|---|---|---|---|---|---|---|---|---|
| 23-99 | 10% | 20% | 30% | 40% | 50% | 50% | 50% | 50% | 50% | 50% | 40% | 30% | 20% | 10% | 0 | 0 | 0 | 0 | 0 | 0 | 0 | 0 |
| 22 | 10% | 20% | 30% | 40% | 50% | 50% | 50% | 50% | 50% | 50% | 40% | 30% | 20% | 10% | 0 | 0 | 0 | 0 | 0 | 0 | 0 | 0 |
| 21 | 10% | 20% | 30% | 40% | 50% | 50% | 50% | 50% | 50% | 50% | 40% | 30% | 20% | 10% | 65% | 65% | 65% | 65% | 65% | 65% | 65% | 0 |
| 20 | 10% | 20% | 30% | 40% | 50% | 50% | 50% | 50% | 50% | 40% | 30% | 20% | 10% | 65% | 65% | 65% | 65% | 65% | 65% | 65% | 0 | 0 |
| 19 | 10% | 20% | 30% | 40% | 50% | 50% | 50% | 50% | 40% | 30% | 20% | 10% | 65% | 65% | 65% | 65% | 65% | 65% | 65% | 0 | 0 | 0 |
| 18 | 10% | 20% | 30% | 40% | 50% | 50% | 50% | 40% | 30% | 20% | 10% | 65% | 65% | 65% | 65% | 65% | 65% | 65% | … | 0 | 0 | 0 |
| 17 | 10% | 20% | 30% | 40% | 50% | 50% | 40% | 30% | 20% | 10% | 65% | 65% | 65% | 65% | 65% | 65% | 65% | 65% | … | 0 | 0 | 0 |
| 16 | 10% | 20% | 30% | 40% | 50% | 40% | 30% | 20% | 10% | 65% | 65% | 65% | 65% | 65% | 65% | 65% | 65% | 65% | … | 0 | 0 | 0 |
| 15 | 10% | 20% | 30% | 40% | 40% | 30% | 20% | 10% | 65% | 65% | 65% | 65% | 65% | 65% | 65% | 65% | 65% | 65% | … | 0 | 0 | 0 |
| 14 | 10% | 20% | 30% | 30% | 30% | 20% | 10% | 65% | 65% | 65% | 65% | 65% | 65% | 65% | 65% | 65% | 65% | 65% | 0 | 0 | 0 | 0 |
| 13 | 10% | 20% | 20% | 20% | 20% | 10% | 65% | 65% | 65% | 65% | 65% | 65% | 65% | 65% | 65% | 65% | 65% | 0 | 0 | 0 | 0 | 0 |
| 12 | 10% | 10% | 10% | 10% | 10% | 65% | 65% | 65% | 65% | 65% | 65% | 65% | 65% | 65% | 65% | 65% | 0 | 0 | 0 | 0 | 0 | 0 |
| … | 0 | 0 | 0 | 0 | 0 | 0 | 0 | 0 | 0 | 0 | 0 | 0 | 0 | 0 | 0 | 0 | 0 | 0 | 0 | 0 | 0 | 0 |
| 0 | 0 | 0 | 0 | 0 | 0 | 0 | 0 | 0 | 0 | 0 | 0 | 0 | 0 | 0 | 0 | 0 | 0 | 0 | 0 | 0 | 0 | 0 |

Year

The numbers in each cell refer to the percentage of all individuals of the corresponding age (y-axis) remaining protected by the vaccine by year's end. Wastage was not included in the example.

*Table S1. Vaccine demand by year and by age of vaccination across 31 countries and Venezuela.*

| | Total vaccine demand by different vaccination campaign scenarios | | | | | | |
|---|---|---|---|---|---|---|---|
| **Type** | **Population-wide** | | | **Routine** | **Population-wide + Routine (P+R)** | | |
| **Target age** | **12-99** | | | **12** | **P: 12-99; R: 12** | | |
| **Vaccination period** | **2025-2029** | | | **2025-2040** | **P: 2025-2029; R: 2030-2040** | | |
| **Annual target coverage** | **2%** | **4%** | **10%** | **65%** | **P: 2%<br>R: 65%** | **P: 4%<br>R: 65%** | **P: 10%<br>R: 65%** |
| **Annual by year** | | | | | | | |
| 2025 | 46.1M | 92.1M | 230M | 33.0M | 46.1M | 92.1M | 230M |
| 2026 | 46.7M | 93.4M | 233M | 32.6M | 46.7M | 93.4M | 233M |
| 2027 | 47.3M | 94.6M | 237M | 32.4M | 47.3M | 94.6M | 237M |
| 2028 | 47.9M | 95.8M | 240M | 32.5M | 47.9M | 95.8M | 240M |
| 2029 | 48.5M | 97.0M | 243M | 32.4M | 48.5M | 97.0M | 243M |
| 2030 | 0 | 0 | 0 | 32.3M | 32.3M | 32.3M | 32.3M |
| 2031 | 0 | 0 | 0 | 32.2M | 32.2M | 32.2M | 32.2M |
| 2032 | 0 | 0 | 0 | 31.8M | 31.8M | 31.8M | 31.8M |
| 2033 | 0 | 0 | 0 | 31.5M | 31.5M | 31.5M | 31.5M |
| 2034 | 0 | 0 | 0 | 31.4M | 31.4M | 31.4M | 31.4M |
| 2035 | 0 | 0 | 0 | 31.5M | 31.5M | 31.5M | 31.5M |
| 2036 | 0 | 0 | 0 | 31.6M | 31.6M | 31.6M | 31.6M |
| 2037 | 0 | 0 | 0 | 31.6M | 31.6M | 31.6M | 31.6M |
| 2038 | 0 | 0 | 0 | 31.6M | 31.6M | 31.6M | 31.6M |
| 2039 | 0 | 0 | 0 | 31.6M | 31.6M | 31.6M | 31.6M |
| 2040 | 0 | 0 | 0 | 31.6M | 31.6M | 31.6M | 31.6M |
| *Total* | *236M* | *473M* | *1.18B* | *511M* | *585M* | *821M* | *1.53B* |
| **Cumulative by age of vaccination** | | | | | | | |
| 0-9 | 0 | 0 | 0 | 0 | 0 | 0 | 0 |
| 10-19 | 40.3M | 80.6M | 202M | 511M | 389M | 429M | 550M |
| 20-29 | 49.2M | 98.4M | 246M | 0 | 49.2M | 98.4M | 246M |
| 30-39 | 44.9M | 89.8M | 225M | 0 | 44.9M | 89.8M | 225M |
| 40-49 | 37.9M | 75.9M | 190M | 0 | 37.9M | 75.9M | 190M |
| 50-59 | 29.1M | 58.3M | 146M | 0 | 29.1M | 58.3M | 146M |
| 60-69 | 20.5M | 40.9M | 102M | 0 | 20.5M | 40.9M | 102M |
| 70-79 | 10.8M | 21.5M | 53.8M | 0 | 10.8M | 21.5M | 53.8M |
| 80+ | 3.73M | 7.45M | 18.6M | 0 | 3.73M | 7.45M | 18.6M |
| *Total* | *236M* | *473M* | *1.18B* | *511M* | *585M* | *821M* | *1.53B* |

The demand figures account for projected age-specific population changes over time, and are the same as the target coverage (actual achieved coverage = 95% of target coverage accounting for 5% wastage). P = population-wide, R = Routine, M = million, B = Billion.

***Table S2. Guidelines for Accurate and Transparent Health Estimates Reporting (GATHER) checklist[1]***

| Item | Checklist item | Reporting |
|---|---|---|
| **Objectives and funding** | | |
| **1** | Define the indicator(s), populations (including age, sex, and geographic entities), and time period(s) for which estimates were made. | Methods [*Model overview, Health and economic outcomes*] |
| **2** | List the funding sources for the work. | Methods [*Role of the funder*] |
| **Data inputs** | | |
| *For all data inputs from multiple sources that are synthesized as part of the study:* | | |
| **3** | Describe how the data were identified and how the data were accessed. | Methods; Supplementary methods |
| **4** | Specify the inclusion and exclusion criteria. Identify all ad-hoc exclusions. | Methods; Supplementary methods |
| **5** | Provide information on all included data sources and their main characteristics. For each data source used, report reference information or contact name/institution, population represented, data collection method, year(s) of data collection, sex and age range, diagnostic criteria or measurement method, and sample size, as relevant. | Supplementary methods |
| **6** | Identify and describe any categories of input data that have potentially important biases (e.g., based on characteristics listed in item 5). | Supplementary methods |
| *For data inputs that contribute to the analysis but were not synthesized as part of the study:* | | |
| **7** | Describe and give sources for any other data inputs. | Supplementary methods |
| *For all data inputs:* | | |
| **8** | Provide all data inputs in a file format from which data can be efficiently extracted (e.g., a spreadsheet rather than a PDF), including all relevant meta-data listed in item 5. For any data inputs that cannot be shared because of ethical or legal reasons, such as third-party ownership, provide a contact name or the name of the institution that retains the right to the data. | Data sharing statement |
| **Data analysis** | | |
| **9** | Provide a conceptual overview of the data analysis method. A diagram may be helpful. | Methods [*Model overview*]; Figure S1 |
| **10** | Provide a detailed description of all steps of the analysis, including mathematical formulae. This description should cover, as relevant, data cleaning, data pre-processing, data adjustments and weighting of data sources, and mathematical or statistical model(s). | Supplementary methods |
| **11** | Describe how candidate models were evaluated and how the final model(s) were selected. | Methods [*Simulation and statistical reporting*]; Supplementary methods |
| **12** | Provide the results of an evaluation of model performance, if done, as well as the results of any relevant sensitivity analysis. | Methods [*Simulation and statistical reporting*]; Table S8-13, Figure S5 |
| **13** | Describe methods for calculating uncertainty of the estimates. State which sources of uncertainty were, and were not, accounted for in the uncertainty analysis. | Methods [*Simulation and statistical reporting*] |
| **14** | State how analytic or statistical source code used to generate estimates can be accessed. | Data sharing statement |
| **Results and Discussion** | | |
| **15** | Provide published estimates in a file format from which data can be efficiently extracted. | Table 1-4, Figure 3, Table S1, S3-S13, Figure S4 |
| **16** | Report a quantitative measure of the uncertainty of the estimates (e.g. uncertainty intervals). | Reported throughout |
| **17** | Interpret results in light of existing evidence. If updating a previous set of estimates, describe the reasons for changes in estimates. | Discussion |
| **18** | Discuss limitations of the estimates. Include a discussion of any modelling assumptions or data limitations that affect interpretation of the estimates. | Discussion |

***Table S3. Projected annual health burden of CHIKV infection over 2025 to 2050 across 31 countries and Venezuela in the absence of vaccination.***

| | **Mean (95% UI) totals in the year** | | | | | |
|---|---|---|---|---|---|---|
| | **2025** | **2030** | **2035** | **2040** | **2045** | **2050** |
| **Health outcome** | | | | | | |
| CHIKV infection (n) | 27.2M (26.0M, 28.6M) | 27.8M (26.6M, 29.1M) | 28.3M (27.1M, 29.6M) | 28.7M (27.5M, 29.9M) | 28.9M (27.8M, 30.2M) | 29.1M (28.0M, 30.3M) |
| Symptomatic chikungunya (n) | 12.7M (11.7M, 13.8M) | 13.1M (12.0M, 14.1M) | 13.4M (12.3M, 14.4M) | 13.6M (12.5M, 14.7M) | 13.8M (12.7M, 14.8M) | 13.9M (12.8M, 14.9M) |
| Hospitalisation (n) | 47.6K (43.7K, 51.3K) | 50.3K (46.3K, 54.2K) | 53.4K (49.2K, 57.3K) | 56.3K (52.0K, 60.4K) | 59.2K (54.7K, 63.4K) | 62.1K (57.5K, 66.5K) |
| Chronic chikungunya (n) | 780K (664K, 902K) | 802K (682K, 925K) | 819K (699K, 945K) | 833K (710K, 960K) | 843K (720K, 970K) | 850K (727K, 977K) |
| Chikungunya death (n) | 1.84K (1.58K, 2.13K) | 1.92K (1.65K, 2.20K) | 2.00K (1.72K, 2.28K) | 2.07K (1.78K, 2.36K) | 2.13K (1.84K, 2.42K) | 2.19K (1.89K, 2.48K) |
| **DALY due to different reasons** | | | | | | |
| Acute mild/Moderate symptoms | 5.54K (2.45K, 9.70K) | 5.69K (2.51K, 9.94K) | 5.81K (2.57K, 10.1K) | 5.90K (2.61K, 10.3K) | 5.96K (2.64K, 10.4K) | 6.01K (2.66K, 10.4K) |
| Acute severe symptoms | 1.19K (532, 1.90K) | 1.26K (563, 2.02K) | 1.34K (598, 2.14K) | 1.41K (632, 2.26K) | 1.48K (665, 2.39K) | 1.55K (698, 2.50K) |
| Chronic chikungunya | 128K (103K, 158K) | 133K (106K, 164K) | 137K (110K, 170K) | 141K (113K, 174K) | 144K (115K, 178K) | 146K (117K, 181K) |
| Chikungunya death | 79.8K (65.0K, 97.0K) | 81.5K (66.6K, 98.6K) | 83.2K (68.4K, 100K) | 84.6K (69.7K, 101K) | 85.7K (70.8K, 102K) | 86.5K (71.7K, 103K) |

CHIKV = Chikungunya virus, DALY = disability-adjusted life-year, K = thousand, M = million

***Table S4. Projected cumulative health burden of CHIKV infection by age groups from 2025 to 2050 across 31 countries and Venezuela in the absence of vaccination.***

| | **Mean results (95% UI) in the age group** | | | | | | | | |
|---|---|---|---|---|---|---|---|---|---|
| | **0-9** | **10-19** | **20-29** | **30-39** | **40-49** | **50-59** | **60-69** | **70-79** | **80+** |
| **Cumulative totals** | | | | | | | | | |
| Acute symptomatic chikungunya | 37.6M (30.2M, 45.7M) | 55.7M (45.6M, 66.6M) | 66.8M (53.5M, 78.9M) | 61.4M (49.7M, 71.6M) | 47.7M (37.9M, 55.8M) | 39.6M (31.6M, 46.3M) | 23.4M (18.8M, 27.9M) | 11.7M (9.40M, 13.9M) | 5.21M (3.96M, 6.44M) |
| Hospitalisation | 298K (264K, 333K) | 98.5K (84.9K, 113K) | 135K (119K, 151K) | 115K (102K, 129K) | 70.2K (60.4K, 80.6K) | 90.1K (78.7K, 103K) | 154K (139K, 171K) | 251K (225K, 277K) | 212K (190K, 235K) |
| Chikungunya death | 11.4K (7.40K, 16.9K) | 6.33K (3.46K, 9.93K) | 5.88K (3.55K, 8.66K) | 4.79K (3.01K, 6.97K) | 5.46K (3.53K, 7.63K) | 5.90K (4.21K, 7.92K) | 2.81K (1.79K, 4.18K) | 5.83K (4.18K, 7.71K) | 4.26K (2.81K, 5.95K) |
| Chronic chikungunya | 2.30M (1.78M, 2.85M) | 3.41M (2.68M, 4.30M) | 4.09M (3.16M, 5.08M) | 3.76M (2.91M, 4.57M) | 2.92M (2.27M, 3.56M) | 2.42M (1.89M, 2.93M) | 1.43M (1.11M, 1.78M) | 711K (553K, 894K) | 317K (235K, 402K) |
| DALYs | 1.13M (810K, 1.54M) | 870K (629K, 1.15M) | 894K (664K, 1.15M) | 742K (554K, 935K) | 870K (620K, 1.14M) | 720K (527K, 928K) | 389K (278K, 509K) | 239K (177K, 309K) | 107K (75.6K, 141K) |

| | Mean results (95% UI) in the age group | | | | | | | | |
|---|---|---|---|---|---|---|---|---|---|
| | 0-9 | 10-19 | 20-29 | 30-39 | 40-49 | 50-59 | 60-69 | 70-79 | 80+ |
| **Cumulative totals per 100,000 person-years** | | | | | | | | | |
| Acute symptomatic chikungunya | 299 (240, 363) | 438 (358, 523) | 525 (420, 620) | 497 (402, 579) | 424 (336, 495) | 424 (338, 495) | 341 (274, 405) | 286 (231, 341) | 302 (230, 373) |
| Hospitalisation | 2.37 (2.10, 2.65) | 0.773 (0.666, 0.886) | 1.06 (0.932, 1.19) | 0.93 (0.825, 1.04) | 0.623 (0.536, 0.716) | 0.963 (0.841, 1.10) | 2.24 (2.02, 2.48) | 6.16 (5.53, 6.81) | 12.3 (11.0, 13.6) |
| Chikungunya death | 0.09 (0.059, 0.134) | 0.05 (0.027, 0.078) | 0.046 (0.028, 0.068) | 0.039 (0.024, 0.056) | 0.048 (0.031, 0.068) | 0.063 (0.045, 0.085) | 0.041 (0.026, 0.061) | 0.143 (0.103, 0.189) | 0.247 (0.163, 0.345) |
| Chronic chikungunya | 18.3 (14.1, 22.6) | 26.8 (21.1, 33.8) | 32.2 (24.8, 40.0) | 30.4 (23.5, 37.0) | 25.9 (20.1, 31.6) | 25.9 (20.2, 31.3) | 20.9 (16.2, 25.8) | 17.5 (13.6, 21.9) | 18.4 (13.6, 23.3) |
| DALYs | 8.99 (6.43, 12.2) | 6.83 (4.94, 9.07) | 7.03 (5.22, 9.04) | 6.00 (4.48, 7.56) | 7.72 (5.50, 10.1) | 7.70 (5.63, 9.92) | 5.65 (4.04, 7.40) | 5.87 (4.35, 7.58) | 6.18 (4.38, 8.16) |

CHIKV = Chikungunya virus, DALY = disability-adjusted life-year, K = thousand, M = million

#### *Table S5. Projected cumulative economic burden of CHIKV infection over 2025 to 2050 across 31 countries in the absence of vaccination.*

| | Cumulative totals (95%UI) | | | Cumulative totals per 100,000 person-year (95% UI) | | |
|---|---|---|---|---|---|---|
| | Healthcare costs | Productivity loss | Monetized DALYs | Healthcare costs | Productivity loss | Monetized DALYs |
| **Acute mild/moderate symptoms** | 1.30B (1.20B, 1.40B) | 32.7B (15.8B, 50.0B) | 540M (237M, 943M) | 1.57K (1.45K, 1.69K) | 39.4K (19.0K, 60.2K) | 651 (286, 1.14K) |
| **Acute severe symptoms** | 250M (231M, 268M) | 132M (61.8M, 200M) | 149M (66.3M, 239M) | 301 (278, 324) | 159 (74.5, 241) | 179 (79.9, 288) |
| **Chronic chikungunya** | 285M (222M, 368M) | 19.9B (15.8B, 24.5B) | 13.5B (10.8B, 16.6B) | 344 (267, 444) | 24.0K (19.1K, 29.6K) | 16.2K (13.0K, 20.0K) |
| **Chikungunya death** | - | 4.13B (3.36B, 4.92B) | 3.62B (3.10B, 4.17B) | - | 4.98K (4.06K, 5.93K) | 4.37K (3.74K, 5.02K) |

Venezuela was excluded from the analyses because there is no reliable estimate of economic parameter available in the country due to its economic collapse in the recent years.

CHIKV = Chikungunya virus, DALY = disability-adjusted life-year, K = thousand, M = million, B = billion

#### *Table S6. Projected cumulative CHIKV infections, DALYs and societal costs over 2025 to 2050 by 31 countries and Venezuela in the absence of vaccination*

| Country | Cumulative total infections | | Cumulative total DALYs | | Cumulative total societal costs (I$ 2023) | |
|---|---|---|---|---|---|---|
| | Mean (95%UI) | Mean per 100,000 person years (95%UI) | Mean (95%UI) | Mean per 100,000 person years (95%UI) | Mean (95%UI) | Mean per 100,000 person years (95%UI) |
| **Belize** | 109K (98.5K, 120K) | 877 (793, 969) | 903 (745, 1.09K) | 7.27 (6.00, 8.78) | 17.0M (12.6M, 21.6M) | 137K (102K, 174K) |
| **Bolivia** | 1.98M (1.77M, 2.23M) | 527 (471, 593) | 15.9K (13.2K, 19.0K) | 4.22 (3.50, 5.05) | 266M (198M, 338M) | 70.8K (52.5K, 89.8K) |
| **Brazil** | 58.4M (53.6M, 64.1M) | 1.03K (947, 1.13K) | 510K (426K, 610K) | 9.03 (7.53, 10.8) | 14.4B (11.1B, 18.0B) | 255K (196K, 319K) |
| **Cambodia** | 4.88M (4.52M, 5.27M) | 935 (867, 1.01K) | 38.3K (32.5K, 45.6K) | 7.35 (6.23, 8.75) | 246M (172M, 320M) | 47.3K (33.1K, 61.3K) |

| | | | | | | |
|---|---|---|---|---|---|---|
| **Chad** | 7.11M (6.43M, 7.90M) | 924 (835, 1.03K) | 49.3K (40.8K, 59.5K) | 6.40 (5.29, 7.72) | 76.0M (55.5M, 96.5M) | 9.87K (7.21K, 12.5K) |
| **Colombia** | 12.1M (11.0M, 13.4M) | 811 (737, 899) | 107K (88.1K, 128K) | 7.13 (5.89, 8.59) | 3.83B (2.96B, 4.76B) | 256K (198K, 319K) |
| **Costa Rica** | 1.45M (1.32M, 1.58M) | 1.05K (954, 1.14K) | 13.1K (10.9K, 15.6K) | 9.49 (7.90, 11.3) | 527M (411M, 647M) | 381K (297K, 468K) |
| **Democratic Republic of Congo** | 44.2M (40.1M, 48.8M) | 1.04K (944, 1.15K) | 323K (267K, 387K) | 7.61 (6.28, 9.12) | 419M (303M, 536M) | 9.88K (7.14K, 12.6K) |
| **Djibouti** | 248K (225K, 275K) | 699 (632, 773) | 1.93K (1.61K, 2.31K) | 5.44 (4.52, 6.49) | 6.38M (4.49M, 8.28M) | 17.9K (12.6K, 23.3K) |
| **Dominican Republic** | 3.28M (3.01M, 3.62M) | 1.02K (935, 1.13K) | 27.5K (22.8K, 33.0K) | 8.54 (7.10, 10.3) | 763M (547M, 990M) | 237K (170K, 308K) |
| **Ecuador** | 3.66M (3.30M, 4.08M) | 704 (634, 783) | 31.6K (26.2K, 38.0K) | 6.08 (5.03, 7.31) | 724M (547M, 911M) | 139K (105K, 175K) |
| **El Salvador** | 1.85M (1.68M, 2.04M) | 1.08K (984, 1.19K) | 15.5K (12.8K, 18.7K) | 9.05 (7.49, 10.9) | 259M (195M, 323M) | 151K (114K, 189K) |
| **Ethiopia** | 26.8M (24.0M, 30.1M) | 573 (515, 644) | 204K (167K, 248K) | 4.38 (3.57, 5.30) | 591M (419M, 775M) | 12.6K (8.96K, 16.6K) |
| **Grenada** | 21.5K (19.3K, 24.0K) | 712 (639, 794) | 187 (152, 228) | 6.20 (5.03, 7.55) | 4.08M (2.94M, 5.34M) | 135K (97.4K, 177K) |
| **Guatemala** | 5.22M (4.75M, 5.79M) | 917 (835, 1.02K) | 42.3K (35.4K, 50.4K) | 7.43 (6.21, 8.85) | 668M (485M, 864M) | 117K (85.2K, 152K) |
| **Guyana** | 204K (184K, 226K) | 876 (791, 974) | 1.64K (1.36K, 1.96K) | 7.06 (5.85, 8.45) | 60.1M (41.3M, 79.7M) | 258K (178K, 343K) |
| **Haiti** | 3.44M (3.10M, 3.80M) | 988 (890, 1.09K) | 26.8K (22.5K, 31.9K) | 7.69 (6.46, 9.15) | 94.0M (66.7M, 124M) | 27.0K (19.1K, 35.5K) |
| **Honduras** | 3.59M (3.27M, 3.94M) | 1.06K (963, 1.16K) | 29.0K (24.0K, 34.6K) | 8.55 (7.08, 10.2) | 252M (192M, 309M) | 74.4K (56.6K, 91.1K) |
| **India** | 369M (341M, 398M) | 891 (824, 962) | 2.97M (2.50M, 3.50M) | 7.19 (6.04, 8.47) | 24.1B (17.0B, 31.1B) | 58.2K (41.2K, 75.2K) |
| **Indonesia** | 70.3M (65.4M, 75.8M) | 882 (820, 950) | 561K (473K, 669K) | 7.03 (5.94, 8.40) | 8.40B (5.96B, 10.8B) | 105K (74.8K, 135K) |
| **Malaysia** | 9.66M (8.90M, 10.5M) | 917 (845, 999) | 81.5K (68.6K, 96.8K) | 7.74 (6.51, 9.19) | 3.27B (2.43B, 4.11B) | 310K (230K, 390K) |
| **Mexico** | 28.5M (25.8M, 31.8M) | 772 (699, 860) | 244K (203K, 294K) | 6.60 (5.49, 7.96) | 7.77B (5.83B, 9.92B) | 210K (158K, 269K) |
| **Nicaragua** | 2.24M (2.06M, 2.47M) | 1.08K (993, 1.19K) | 18.6K (15.5K, 22.3K) | 8.99 (7.50, 10.8) | 214M (166M, 267M) | 103K (80.1K, 129K) |
| **Panama** | 1.36M (1.24M, 1.49M) | 1.02K (924, 1.12K) | 11.9K (9.88K, 14.3K) | 8.89 (7.38, 10.6) | 608M (465M, 765M) | 454K (347K, 571K) |
| **Paraguay** | 2.04M (1.85M, 2.26M) | 996 (902, 1.10K) | 16.8K (14.1K, 20.1K) | 8.21 (6.88, 9.78) | 383M (284M, 486M) | 187K (139K, 237K) |
| **Peru** | 7.59M (6.87M, 8.43M) | 768 (696, 854) | 66.0K (54.1K, 79.5K) | 6.69 (5.48, 8.05) | 1.52B (1.13B, 1.95B) | 154K (114K, 197K) |
| **Philippines** | 27.7M (25.5M, 30.1M) | 839 (773, 911) | 217K (182K, 259K) | 6.58 (5.52, 7.84) | 2.55B (1.84B, 3.30B) | 77.1K (55.8K, 100.0K) |
| **Republic of Congo** | 1.90M (1.72M, 2.11M) | 844 (765, 939) | 14.3K (11.8K, 17.2K) | 6.36 (5.26, 7.62) | 71.2M (50.1M, 93.3M) | 31.6K (22.2K, 41.4K) |
| **St. Lucia** | 39.4K (35.6K, 43.7K) | 846 (765, 939) | 337 (274, 407) | 7.24 (5.89, 8.74) | 11.2M (8.05M, 14.5M) | 241K (173K, 313K) |
| **Sudan** | 16.7M (15.2M, 18.5M) | 939 (852, 1.04K) | 126K (105K, 151K) | 7.07 (5.89, 8.49) | 217M (165M, 274M) | 12.2K (9.29K, 15.4K) |
| **Thailand** | 14.4M (13.6M, 15.2M) | 793 (749, 839) | 125K (107K, 146K) | 6.93 (5.90, 8.07) | 4.23B (3.22B, 5.30B) | 234K (178K, 293K) |
| **Venezuela** | 8.05M (7.37M, 8.87M) | 1.03K (944, 1.14K) | 66.8K (55.7K, 80.8K) | 8.56 (7.13, 10.3) | -* | -* |

* Venezuela was excluded from the analyses for the economic impact because there is no reliable estimate of economic parameter available in the country due to its economic collapse in the recent years. Future costs and life-years are discounted annually at 3.5%/year. DALY = disability-adjusted life-year, I$ = International dollar, K = thousand, M = million, B = billion.

***Figure S4. Vaccine efficiency in preventing DALY over 2025-2050 by different vaccination strategies in each of the 31 countries and Venezuela.***

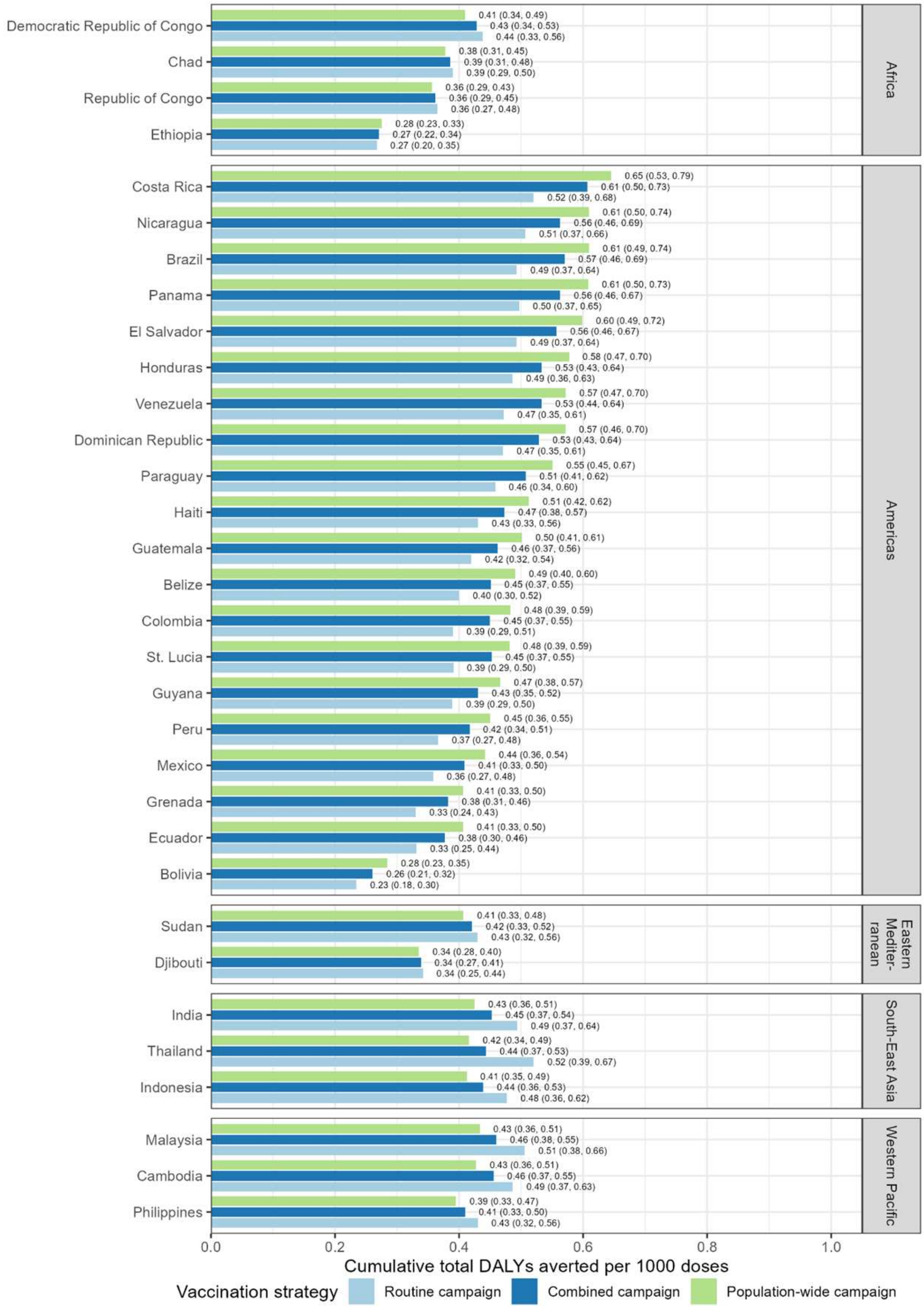

Combined campaign analysed starts with the population-wide campaign over 5 years with 20% target coverage in total (actual achieved coverage = 95% of target coverage accounting for 5% wastage). DALY = disability-adjusted life-year.

***Table S7. Countries with threshold vaccination costs above certain threshold price per dose***

| | **Based on population-wide campaign** | | **Based on routine campaign** | |
|---|---|---|---|---|
| | **Countries with threshold vaccination costs above $20*** | **Countries with threshold vaccination costs above $40&** | **Countries with threshold vaccination costs above $20*** | **Countries with threshold vaccination costs above $40&** |
| N (%^) | 11 (35.5%) | 1 (3.2%) | 1 (3.2%) | 0 |
| Name | Brazil, Colombia, Costa Rica, Dominican Republic, Guyana, Malaysia, Mexico, Panama, Paraguay, St. Lucia, Thailand | Panama | Panama | None |

^: the denominator is 31 (Venezuela was excluded from the analyses for the economic impact because there is no reliable estimate of economic parameter available in the country due to its economic collapse in the recent years)

*: Based on $15 per dose for HPV vaccine in low- and middle-income countries in Latin America through the Revolving Fund of the Pan American Health Organization (PAHO)[2] and $5 delivery costs

&: Based on double of $20 per dose used above and close to the median price per dose for HPV in high income country[3]

***Figure S5. Sensitivity analyses of projected cumulative total health burden of CHIKV infection, stratified by age, from 2025 to 2050 across 31 countries and Venezuela in the absence of vaccination.***

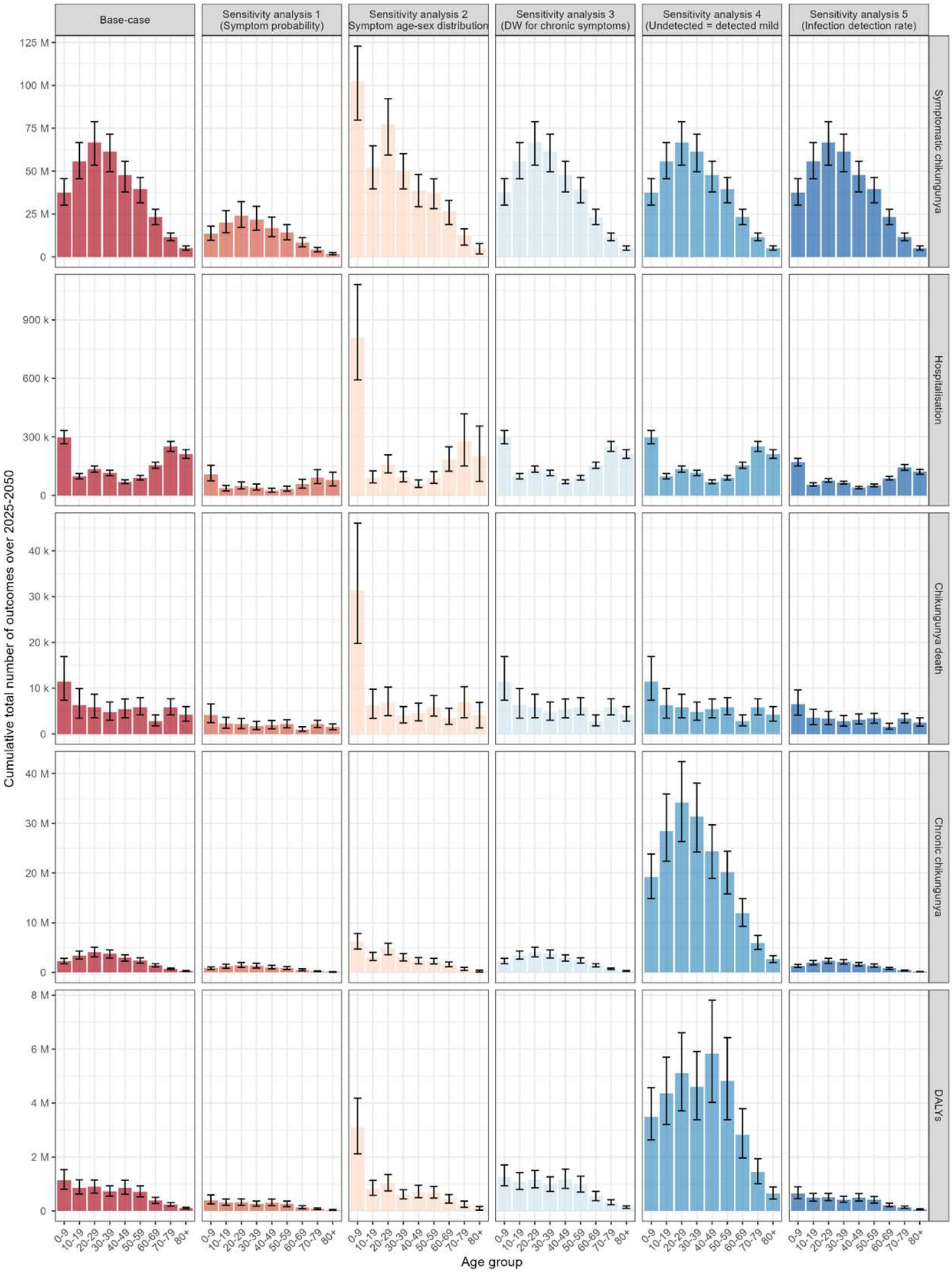

Sensitivity analysis for symptomatic infection used estimates with lower probability of symptomatic infection than base-case [Mean (95%CI): 0.180 (0.110, 0.260) vs. 0.505 (0.380, 0.627)]. Sensitivity analysis for symptom age-sex distribution used alternative distribution of symptom by age and sex [supplementary method 2d]. Sensitivity analysis for DW for chronic symptoms used estimates with higher disability weight for chronic chikungunya [Mean (95%CI): 0.331 (0.306, 0.355) vs. 0.229 (0.216, 0.242)]. Sensitivity analysis "undetected = detected mild" used estimates with higher disability weight for undetected symptomatic infections [mean (95CI): 0.110 (0.074, 0.145) vs. 0.006 (0.002, 0.012)] and the same probability of chronic symptoms in undetected symptomatic infections as detected symptomatic infections (base-case: no chronic symptoms). Sensitivity analysis “infection detection rate” used estimates with lower rate of detection of infection in healthcare system [0.36 vs. 0.63], which impacts on the subsequent parameters including probability of detected mild symptoms, hospitalization and chronic symptoms.

***Table S8. Projected cumulative health outcomes averted due to CHIKV vaccination from 2025 to 2050 across 31 countries and Venezuela, depending on the vaccination campaign and the vaccine's efficacy against disease.***

| **Health outcomes averted** | **Cumulative totals (95% UI) by different vaccination campaign strategies and levels of vaccine efficacy** | | | | | | | |
|---|---|---|---|---|---|---|---|---|
| | **Type** | **Population wide** | | | **Routine** | **Population-wide + Routine (P+R)** | | |
| | **Age** | **12-99** | | | **12** | **P: 12-99; R: 12** | | |
| | **Vaccination period** | **2025-2029** | | | **2025-2040** | **P: 2025-2029; R: 2030-2040** | | |
| | **Annual target coverage^** | **2%** | **4%** | **10%** | **65%** | **P: 2%; R: 65%** | **P: 4%; R: 65%** | **P: 10%; R: 65%** |
| Symptomatic chikungunya (n) | Vaccine efficacy: 90% | 8.81M (8.05M, 9.49M) | 17.6M (16.1M, 19.0M) | 44.1M (40.3M, 47.4M) | 19.8M (16.8M, 22.9M) | 22.2M (19.7M, 24.8M) | 31.1M (28.0M, 34.1M) | 57.5M (52.4M, 62.4M) |
| | Vaccine efficacy: 70% | 6.85M (6.26M, 7.38M) | 13.7M (12.5M, 14.8M) | 34.3M (31.3M, 36.9M) | 15.4M (13.0M, 17.8M) | 17.3M (15.3M, 19.3M) | 24.2M (21.8M, 26.5M) | 44.7M (40.8M, 48.6M) |
| | Vaccine efficacy: 50% | 4.90M (4.47M, 5.27M) | 9.79M (8.94M, 10.5M) | 24.5M (22.4M, 26.4M) | 11.0M (9.31M, 12.7M) | 12.4M (10.9M, 13.8M) | 17.3M (15.6M, 19.0M) | 31.9M (29.1M, 34.7M) |
| Hospitalisation (n) | Vaccine efficacy: 90% | 31.8K (29.3K, 34.1K) | 63.5K (58.6K, 68.1K) | 159K (147K, 170K) | 36.0K (31.6K, 40.6K) | 56.2K (51.4K, 60.9K) | 88.0K (80.9K, 94.9K) | 183K (169K, 197K) |
| | Vaccine efficacy: 70% | 24.7K (22.8K, 26.5K) | 49.4K (45.6K, 53.0K) | 124K (114K, 132K) | 28.0K (24.6K, 31.6K) | 43.7K (39.9K, 47.4K) | 68.5K (62.9K, 73.8K) | 143K (131K, 153K) |
| | Vaccine efficacy: 50% | 17.6K (16.3K, 18.9K) | 35.3K (32.6K, 37.8K) | 88.2K (81.5K, 94.6K) | 20.0K (17.5K, 22.5K) | 31.2K (28.5K, 33.8K) | 48.9K (45.0K, 52.7K) | 102K (93.7K, 109K) |
| Chronic chikungunya (n) | Vaccine efficacy: 90% | 539K (457K, 620K) | 1.08M (915K, 1.24M) | 2.70M (2.29M, 3.10M) | 1.21M (977K, 1.48M) | 1.36M (1.14M, 1.61M) | 1.90M (1.61M, 2.23M) | 3.52M (2.99M, 4.09M) |
| | Vaccine efficacy: 70% | 419K (356K, 482K) | 839K (712K, 965K) | 2.10M (1.78M, 2.41M) | 941K (760K, 1.15M) | 1.06M (884K, 1.26M) | 1.48M (1.25M, 1.73M) | 2.74M (2.32M, 3.18M) |
| | Vaccine efficacy: 50% | 300K (254K, 345K) | 599K (508K, 689K) | 1.50M (1.27M, 1.72M) | 672K (543K, 823K) | 756K (632K, 897K) | 1.06M (894K, 1.24M) | 1.96M (1.66M, 2.27M) |
| Death (n) | Vaccine efficacy: 90% | 1.14K (978, 1.31K) | 2.29K (1.96K, 2.61K) | 5.72K (4.89K, 6.53K) | 2.13K (1.36K, 3.10K) | 2.59K (2.01K, 3.35K) | 3.74K (3.00K, 4.58K) | 7.17K (6.00K, 8.36K) |
| | Vaccine efficacy: 70% | 890 (761, 1.02K) | 1.78K (1.52K, 2.03K) | 4.45K (3.80K, 5.08K) | 1.66K (1.05K, 2.41K) | 2.02K (1.56K, 2.61K) | 2.91K (2.33K, 3.56K) | 5.58K (4.67K, 6.50K) |
| | Vaccine efficacy: 50% | 636 (543, 726) | 1.27K (1.09K, 1.45K) | 3.18K (2.72K, 3.63K) | 1.18K (753, 1.72K) | 1.44K (1.11K, 1.86K) | 2.08K (1.66K, 2.55K) | 3.98K (3.33K, 4.64K) |
| DALYs (n) | Vaccine efficacy: 90% | 135K (113K, 159K) | 269K (226K, 318K) | 673K (564K, 795K) | 298K (224K, 380K) | 337K (275K, 408K) | 472K (391K, 559K) | 876K (731K, 1.03M) |

| Health outcomes averted | Cumulative totals (95% UI) by different vaccination campaign strategies and levels of vaccine efficacy | | | | | | | |
|---|---|---|---|---|---|---|---|---|
| | **Type** | **Population wide** | | | **Routine** | **Population-wide + Routine (P+R)** | | |
| | **Age** | **12-99** | | | **12** | **P: 12-99; R: 12** | | |
| | **Vaccination period** | **2025-2029** | | | **2025-2040** | **P: 2025-2029; R: 2030-2040** | | |
| | **Annual target coverage^** | **2%** | **4%** | **10%** | **65%** | **P: 2%; R: 65%** | **P: 4%; R: 65%** | **P: 10%; R: 65%** |
| | Vaccine efficacy: 70% | 105K (87.7K, 124K) | 209K (175K, 247K) | 523K (439K, 619K) | 232K (174K, 296K) | 262K (214K, 317K) | 367K (304K, 435K) | 681K (569K, 802K) |
| | Vaccine efficacy: 50% | 74.7K (62.6K, 88.3K) | 149K (125K, 177K) | 373K (313K, 441K) | 165K (124K, 211K) | 187K (153K, 227K) | 262K (217K, 310K) | 486K (406K, 572K) |

^ Actual achieved coverage = 95% of target coverage, accounting for 5% wastage. DALY = disability-adjusted life-year, K = thousand, M = million.

*Table S9. Projected cumulative economic costs averted due to CHIKV vaccination from 2025 to 2050 across 31 countries, depending on the vaccination campaign and the vaccine's efficacy against disease.*

| **Economic costs averted** | **Cumulative totals (95%UI) by different vaccination campaign strategies and levels of vaccine efficacy** | | | | | | | |
|---|---|---|---|---|---|---|---|---|
| | **Type** | **Population wide** | | | **Routine** | **Population-wide + Routine (P+R)** | | |
| | **Age** | **12-99** | | | **12** | **P: 12-99; R: 12** | | |
| | **Vaccination period** | **2025-2029** | | | **2025-2040** | **P: 2025-2029; R: 2030-2040** | | |
| | **Annual target coverage^** | **2%** | **4%** | **10%** | **65%** | **P: 2%; R: 65%** | **P: 4%; R: 65%** | **P: 10%; R: 65%** |
| Healthcare costs (I$ 2023) | Vaccine efficacy: 90% | 58.7M (53.2M, 64.0M) | 117M (106M, 128M) | 293M (266M, 320M) | 83.3M (70.7M, 95.9M) | 109M (98.9M, 121M) | 168M (153M, 185M) | 344M (313M, 376M) |
| | Vaccine efficacy: 70% | 45.5M (41.2M, 49.7M) | 91.0M (82.5M, 99.3M) | 228M (206M, 248M) | 64.7M (54.9M, 74.5M) | 84.8M (76.7M, 94.2M) | 130M (118M, 143M) | 267M (242M, 292M) |
| | Vaccine efficacy: 50% | 32.4M (29.3M, 35.3M) | 64.7M (58.6M, 70.7M) | 162M (147M, 177M) | 46.1M (39.1M, 53.1M) | 60.4M (54.6M, 67.1M) | 92.7M (84.2M, 102M) | 190M (172M, 208M) |
| Catastrophic healthcare expenditure (n) | Vaccine efficacy: 90% | 11.5K (10.4K, 12.5K) | 23.0K (20.8K, 25.0K) | 57.5K (52.1K, 62.6K) | 13.4K (11.6K, 15.2K) | 21.0K (18.9K, 22.9K) | 32.5K (29.5K, 35.2K) | 67.0K (60.9K, 72.7K) |
| | Vaccine efficacy: 70% | 8.92K (8.08K, 9.72K) | 17.8K (16.2K, 19.4K) | 44.6K (40.4K, 48.6K) | 10.4K (9.03K, 11.8K) | 16.3K (14.7K, 17.8K) | 25.2K (22.9K, 27.4K) | 52.0K (47.3K, 56.5K) |
| | Vaccine efficacy: 50% | 6.35K (5.74K, 6.92K) | 12.7K (11.5K, 13.8K) | 31.7K (28.7K, 34.6K) | 7.45K (6.44K, 8.44K) | 11.6K (10.4K, 12.7K) | 18.0K (16.3K, 19.5K) | 37.0K (33.6K, 40.2K) |
| Impoverishing healthcare expenditure (n) | Vaccine efficacy: 90% | 10.9K (9.60K, 12.2K) | 21.7K (19.2K, 24.3K) | 54.3K (48.0K, 60.8K) | 9.16K (7.72K, 10.7K) | 17.1K (15.1K, 19.2K) | 27.9K (24.7K, 31.2K) | 60.5K (53.5K, 67.4K) |
| | Vaccine efficacy: 70% | 8.43K (7.45K, 9.44K) | 16.9K (14.9K, 18.9K) | 42.1K (37.2K, 47.2K) | 7.12K (6.00K, 8.34K) | 13.3K (11.7K, 14.9K) | 21.7K (19.1K, 24.2K) | 47.0K (41.5K, 52.4K) |
| | Vaccine efficacy: 50% | 6.00K (5.30K, 6.72K) | 12.0K (10.6K, 13.4K) | 30.0K (26.5K, 33.6K) | 5.08K (4.28K, 5.95K) | 9.45K (8.34K, 10.6K) | 15.4K (13.6K, 17.3K) | 33.4K (29.6K, 37.3K) |
| Productivity losses (I$ 2023) | Vaccine efficacy: 90% | 2.14B (1.43B, 2.83B) | 4.27B (2.86B, 5.66B) | 10.7B (7.15B, 14.2B) | 1.39B (940M, 1.89B) | 2.98B (2.03B, 3.95B) | 5.12B (3.47B, 6.76B) | 11.5B (7.74B, 15.3B) |
| | Vaccine efficacy: 70% | 1.66B (1.11B, 2.20B) | 3.32B (2.22B, 4.41B) | 8.31B (5.56B, 11.0B) | 1.08B (731M, 1.47B) | 2.32B (1.58B, 3.07B) | 3.98B (2.70B, 5.26B) | 8.97B (6.02B, 11.9B) |
| | Vaccine efficacy: 50% | 1.19B (795M, 1.57B) | 2.37B (1.59B, 3.15B) | 5.94B (3.97B, 7.87B) | 770M (522M, 1.05B) | 1.66B (1.13B, 2.19B) | 2.84B (1.93B, 3.75B) | 6.41B (4.30B, 8.49B) |
| | Vaccine efficacy: 90% | 586M (482M, 707M) | 1.17B (963M, 1.41B) | 2.93B (2.41B, 3.53B) | 657M (498M, 829M) | 984M (813M, 1.18B) | 1.57B (1.30B, 1.87B) | 3.33B (2.76B, 3.99B) |

| Economic costs averted | | Cumulative totals (95%UI) by different vaccination campaign strategies and levels of vaccine efficacy | | | | | | |
|---|---|---|---|---|---|---|---|---|
| | Type | Population wide | | | Routine | Population-wide + Routine (P+R) | | |
| | Age | 12-99 | | | 12 | P: 12-99; R: 12 | | |
| | Vaccination period | 2025-2029 | | | 2025-2040 | P: 2025-2029; R: 2030-2040 | | |
| | Annual target coverage^ | 2% | 4% | 10% | 65% | P: 2%; R: 65% | P: 4%; R: 65% | P: 10%; R: 65% |
| Monetised DALYs (I$ 2023) | Vaccine efficacy: 70% | 456M (374M, 549M) | 911M (749M, 1.10B) | 2.28B (1.87B, 2.75B) | 510M (387M, 644M) | 765M (632M, 914M) | 1.22B (1.01B, 1.45B) | 2.59B (2.15B, 3.10B) |
| | Vaccine efficacy: 50% | 325M (267M, 392M) | 650M (534M, 784M) | 1.63B (1.34B, 1.96B) | 364M (276M, 460M) | 546M (451M, 652M) | 871M (722M, 1.04B) | 1.85B (1.53B, 2.21B) |
| VSL losses (I$ 2023) | Vaccine efficacy: 90% | 4.16B (3.57B, 4.73B) | 8.32B (7.13B, 9.47B) | 20.8B (17.8B, 23.7B) | 5.65B (3.54B, 8.26B) | 7.59B (6.04B, 9.52B) | 11.8B (9.69B, 14.1B) | 24.2B (20.5B, 28.0B) |
| | Vaccine efficacy: 70% | 3.24B (2.77B, 3.68B) | 6.47B (5.55B, 7.36B) | 16.2B (13.9B, 18.4B) | 4.39B (2.76B, 6.42B) | 5.90B (4.69B, 7.41B) | 9.14B (7.54B, 11.0B) | 18.8B (16.0B, 21.7B) |
| | Vaccine efficacy: 50% | 2.31B (1.98B, 2.63B) | 4.62B (3.96B, 5.26B) | 11.6B (9.91B, 13.1B) | 3.14B (1.97B, 4.59B) | 4.22B (3.35B, 5.29B) | 6.53B (5.38B, 7.82B) | 13.5B (11.4B, 15.5B) |
| VSLY losses (I$ 2023) | Vaccine efficacy: 90% | 3.43B (2.84B, 4.04B) | 6.86B (5.69B, 8.08B) | 17.2B (14.2B, 20.2B) | 9.10B (5.65B, 13.4B) | 8.95B (6.57B, 12.0B) | 12.4B (9.49B, 15.9B) | 22.7B (18.3B, 27.8B) |
| | Vaccine efficacy: 70% | 2.67B (2.21B, 3.14B) | 5.34B (4.42B, 6.28B) | 13.3B (11.1B, 15.7B) | 7.08B (4.39B, 10.4B) | 6.96B (5.11B, 9.35B) | 9.63B (7.38B, 12.4B) | 17.6B (14.2B, 21.6B) |
| | Vaccine efficacy: 50% | 1.91B (1.58B, 2.24B) | 3.81B (3.16B, 4.49B) | 9.53B (7.90B, 11.2B) | 5.05B (3.14B, 7.45B) | 4.97B (3.65B, 6.68B) | 6.88B (5.27B, 8.83B) | 12.6B (10.2B, 15.5B) |
| Societal costs (I$ 2023) | Vaccine efficacy: 90% | 2.78B (2.03B, 3.52B) | 5.56B (4.06B, 7.04B) | 13.9B (10.2B, 17.6B) | 2.13B (1.59B, 2.71B) | 4.07B (3.02B, 5.11B) | 6.86B (5.09B, 8.62B) | 15.2B (11.2B, 19.1B) |
| | Vaccine efficacy: 70% | 2.16B (1.58B, 2.74B) | 4.33B (3.16B, 5.48B) | 10.8B (7.90B, 13.7B) | 1.65B (1.24B, 2.10B) | 3.17B (2.35B, 3.98B) | 5.33B (3.96B, 6.71B) | 11.8B (8.73B, 14.9B) |
| | Vaccine efficacy: 50% | 1.54B (1.13B, 1.96B) | 3.09B (2.26B, 3.91B) | 7.72B (5.64B, 9.78B) | 1.18B (884M, 1.50B) | 2.26B (1.68B, 2.84B) | 3.81B (2.83B, 4.79B) | 8.44B (6.23B, 10.6B) |

^ Actual achieved coverage = 95% of target coverage, accounting for 5% wastage. Venezuela was excluded from the analyses for the economic impact because there is no reliable estimate of economic parameter available in the country due to its economic collapse in the recent years. Future costs are discounted annually at 3.5%/year. DALY = disability-adjusted life-year, VSL = value of statistical life, VSLY = value of statistical life-years, I$ = International dollar, K = thousand, M = million, B = billion.

***Table S10. Sensitivity analyses of projected cumulative total health outcomes averted due to CHIKV vaccination from 2025 to 2050, depending on the vaccination campaign.***

| **Outcome** | **Outcome averted (95%UI) by different vaccination campaign scenario** | | | | | | | |
|---|---|---|---|---|---|---|---|---|
| | **Type** | **Population wide** | | | **Routine** | **Population-wide + Routine (P+R)** | | |
| | **Age** | **12-99** | | | **12** | **P: 12-99; R: 12** | | |
| | **Vaccination period** | **2025-2029** | | | **2025-2040** | **P: 2025-2029; R: 2030-2040** | | |
| | **Annual target coverage^** | **2%** | **4%** | **10%** | **65%** | **P: 2%; R: 65%** | **P: 4%; R: 65%** | **P: 10%; R: 65%** |
| Symptomatic chikungunya (n) | Base-case | 6.85M (6.26M, 7.38M) | 13.7M (12.5M, 14.8M) | 34.3M (31.3M, 36.9M) | 15.4M (13.0M, 17.8M) | 17.3M (15.3M, 19.3M) | 24.2M (21.8M, 26.5M) | 44.7M (40.8M, 48.6M) |
| | SA1: Symptomatic probability | 2.46M (2.15M, 2.80M) | 4.91M (4.29M, 5.60M) | 12.3M (10.7M, 14.0M) | 5.54M (4.28M, 7.07M) | 6.22M (5.21M, 7.40M) | 8.68M (7.47M, 10.2M) | 16.1M (14.0M, 18.6M) |
| | SA2: Symptomatic distribution | 6.59M (5.88M, 7.23M) | 13.2M (11.8M, 14.5M) | 32.9M (29.4M, 36.2M) | 15.1M (12.3M, 17.9M) | 16.9M (14.6M, 19.2M) | 23.5M (20.6M, 26.2M) | 43.2M (38.7M, 47.7M) |
| | SA3: Disability weight | 6.85M (6.26M, 7.38M) | 13.7M (12.5M, 14.8M) | 34.3M (31.3M, 36.9M) | 15.4M (13.0M, 17.8M) | 17.3M (15.3M, 19.3M) | 24.2M (21.8M, 26.5M) | 44.7M (40.8M, 48.6M) |
| | SA4: Undetected = detected mild | 6.85M (6.26M, 7.38M) | 13.7M (12.5M, 14.8M) | 34.3M (31.3M, 36.9M) | 15.4M (13.0M, 17.8M) | 17.3M (15.3M, 19.3M) | 24.2M (21.8M, 26.5M) | 44.7M (40.8M, 48.6M) |
| | SA5: Infection detection rate | 6.85M (6.26M, 7.38M) | 13.7M (12.5M, 14.8M) | 34.3M (31.3M, 36.9M) | 15.4M (13.0M, 17.8M) | 17.3M (15.3M, 19.3M) | 24.2M (21.8M, 26.5M) | 44.7M (40.8M, 48.6M) |
| Hospitalisation (n) | Base-case | 24.7K (22.8K, 26.5K) | 49.4K (45.6K, 53.0K) | 124K (114K, 132K) | 28.0K (24.6K, 31.6K) | 43.7K (39.9K, 47.4K) | 68.5K (62.9K, 73.8K) | 143K (131K, 153K) |
| | SA1: Symptomatic probability | 9.07K (7.81K, 10.5K) | 18.1K (15.6K, 21.1K) | 45.4K (39.1K, 52.7K) | 10.3K (7.69K, 13.7K) | 16.1K (13.7K, 19.0K) | 25.1K (21.7K, 29.2K) | 52.3K (45.5K, 60.8K) |
| | SA2: Symptomatic distribution | 25.6K (21.0K, 30.3K) | 51.2K (41.9K, 60.7K) | 128K (105K, 152K) | 28.0K (21.9K, 35.6K) | 44.6K (38.1K, 51.7K) | 70.2K (60.3K, 81.0K) | 147K (124K, 171K) |
| | SA3: Disability weight | 24.7K (22.8K, 26.5K) | 49.4K (45.6K, 53.0K) | 124K (114K, 132K) | 28.0K (24.6K, 31.6K) | 43.7K (39.9K, 47.4K) | 68.5K (62.9K, 73.8K) | 143K (131K, 153K) |
| | SA4: Undetected = detected mild | 24.7K (22.8K, 26.5K) | 49.4K (45.6K, 53.0K) | 124K (114K, 132K) | 28.0K (24.6K, 31.6K) | 43.7K (39.9K, 47.4K) | 68.5K (62.9K, 73.8K) | 143K (131K, 153K) |
| | SA5: Infection detection rate | 14.1K (13.0K, 15.1K) | 28.2K (26.1K, 30.3K) | 70.6K (65.2K, 75.7K) | 16.0K (14.0K, 18.0K) | 25.0K (22.8K, 27.1K) | 39.1K (36.0K, 42.2K) | 81.5K (75.0K, 87.5K) |
| Chronic chikungunya (n) | Base-case | 419K (356K, 482K) | 839K (712K, 965K) | 2.10M (1.78M, 2.41M) | 941K (760K, 1.15M) | 1.06M (884K, 1.26M) | 1.48M (1.25M, 1.73M) | 2.74M (2.32M, 3.18M) |

| Outcome | Outcome averted (95%UI) by different vaccination campaign scenario | | | | | | | |
|---|---|---|---|---|---|---|---|---|
| | Type | Population wide | | | Routine | Population-wide + Routine (P+R) | | |
| | Age | 12-99 | | | 12 | P: 12-99; R: 12 | | |
| | Vaccination period | 2025-2029 | | | 2025-2040 | P: 2025-2029; R: 2030-2040 | | |
| | Annual target coverage^ | 2% | 4% | 10% | 65% | P: 2%; R: 65% | P: 4%; R: 65% | P: 10%; R: 65% |
| | SA1: Symptomatic probability | 150K (125K, 180K) | 301K (249K, 359K) | 752K (623K, 899K) | 339K (254K, 435K) | 381K (303K, 468K) | 531K (428K, 646K) | 983K (808K, 1.17M) |
| | SA2: Symptomatic distribution | 403K (342K, 471K) | 806K (685K, 942K) | 2.02M (1.71M, 2.35M) | 926K (730K, 1.15M) | 1.03M (850K, 1.23M) | 1.44M (1.20M, 1.70M) | 2.65M (2.23M, 3.10M) |
| | SA3: Disability weight | 419K (356K, 482K) | 839K (712K, 965K) | 2.10M (1.78M, 2.41M) | 941K (760K, 1.15M) | 1.06M (884K, 1.26M) | 1.48M (1.25M, 1.73M) | 2.74M (2.32M, 3.18M) |
| | SA4: Undetected = detected mild | 3.50M (2.97M, 4.02M) | 7.00M (5.94M, 8.05M) | 17.5M (14.8M, 20.1M) | 7.85M (6.34M, 9.61M) | 8.83M (7.38M, 10.5M) | 12.3M (10.4M, 14.5M) | 22.8M (19.4M, 26.5M) |
| | SA5: Infection detection rate | 240K (207K, 277K) | 479K (414K, 555K) | 1.20M (1.03M, 1.39M) | 538K (434K, 651K) | 605K (509K, 714K) | 845K (719K, 988K) | 1.56M (1.34M, 1.82M) |
| Death (n) | Base-case | 890 (761, 1.02K) | 1.78K (1.52K, 2.03K) | 4.45K (3.80K, 5.08K) | 1.66K (1.05K, 2.41K) | 2.02K (1.56K, 2.61K) | 2.91K (2.33K, 3.56K) | 5.58K (4.67K, 6.50K) |
| | SA1: Symptomatic probability | 320 (264, 384) | 639 (529, 768) | 1.60K (1.32K, 1.92K) | 594 (352, 888) | 723 (535, 950) | 1.04K (808, 1.30K) | 2.00K (1.62K, 2.41K) |
| | SA2: Symptomatic distribution | 908 (751, 1.07K) | 1.82K (1.50K, 2.14K) | 4.54K (3.76K, 5.34K) | 1.69K (1.05K, 2.47K) | 2.06K (1.55K, 2.64K) | 2.97K (2.36K, 3.65K) | 5.69K (4.67K, 6.73K) |
| | SA3: Disability weight | 890 (761, 1.02K) | 1.78K (1.52K, 2.03K) | 4.45K (3.80K, 5.08K) | 1.66K (1.05K, 2.41K) | 2.02K (1.56K, 2.61K) | 2.91K (2.33K, 3.56K) | 5.58K (4.67K, 6.50K) |
| | SA4: Undetected = detected mild | 890 (761, 1.02K) | 1.78K (1.52K, 2.03K) | 4.45K (3.80K, 5.08K) | 1.66K (1.05K, 2.41K) | 2.02K (1.56K, 2.61K) | 2.91K (2.33K, 3.56K) | 5.58K (4.67K, 6.50K) |
| | SA5: Infection detection rate | 509 (444, 586) | 1.02K (887, 1.17K) | 2.54K (2.22K, 2.93K) | 947 (585, 1.34K) | 1.15K (874, 1.46K) | 1.66K (1.33K, 2.02K) | 3.19K (2.72K, 3.74K) |
| DALYs (n) | Base-case | 105K (87.7K, 124K) | 209K (175K, 247K) | 523K (439K, 619K) | 232K (174K, 296K) | 262K (214K, 317K) | 367K (304K, 435K) | 681K (569K, 802K) |
| | SA1: Symptomatic probability | 37.4K (30.2K, 45.4K) | 74.7K (60.4K, 90.8K) | 187K (151K, 227K) | 83.2K (58.4K, 113K) | 94.0K (71.7K, 118K) | 131K (103K, 162K) | 243K (195K, 294K) |
| | SA2: Symptomatic distribution | 101K (84.1K, 121K) | 202K (168K, 241K) | 505K (421K, 603K) | 231K (172K, 302K) | 258K (208K, 321K) | 359K (296K, 440K) | 663K (553K, 800K) |

| Outcome | Outcome averted (95%UI) by different vaccination campaign scenario | | | | | | | |
|---|---|---|---|---|---|---|---|---|
| | **Type** | **Population wide** | | | **Routine** | **Population-wide + Routine (P+R)** | | |
| | **Age** | **12-99** | | | **12** | **P: 12-99; R: 12** | | |
| | **Vaccination period** | **2025-2029** | | | **2025-2040** | **P: 2025-2029; R: 2030-2040** | | |
| | **Annual target coverage^** | **2%** | **4%** | **10%** | **65%** | **P: 2%; R: 65%** | **P: 4%; R: 65%** | **P: 10%; R: 65%** |
| | SA3: Disability weight | 140K (114K, 168K) | 279K (228K, 335K) | 698K (569K, 838K) | 292K (220K, 373K) | 338K (275K, 411K) | 478K (391K, 572K) | 896K (742K, 1.08M) |
| | SA4: Undetected = detected mild | 659K (533K, 808K) | 1.32M (1.07M, 1.62M) | 3.30M (2.67M, 4.04M) | 1.20M (900K, 1.54M) | 1.47M (1.19M, 1.80M) | 2.13M (1.72M, 2.60M) | 4.11M (3.34M, 5.04M) |
| | SA5: Infection detection rate | 60.1K (50.6K, 71.4K) | 120K (101K, 143K) | 301K (253K, 357K) | 133K (102K, 168K) | 151K (123K, 183K) | 211K (176K, 250K) | 391K (333K, 460K) |

^ Actual achieved coverage = 95% of target coverage, accounting for 5% wastage. All analyses assume 70% vaccine efficacy. Sensitivity analysis for symptomatic infection used estimates with lower probability of symptomatic infection than base-case [Mean (95%CI): 0.180 (0.110, 0.260) vs. 0.505 (0.380, 0.627)]. Sensitivity analysis for symptom age-sex distribution used alternative distribution of symptom by age and sex [supplementary method 2d]. Sensitivity analysis for DW for chronic symptoms used estimates with higher disability weight for chronic chikungunya [Mean (95%CI): 0.331 (0.306, 0.355) vs. 0.229 (0.216, 0.242)]. Sensitivity analysis "undetected = detected mild" used estimates with higher disability weight for undetected symptomatic infections [mean (95CI): 0.110 (0.074, 0.145) vs. 0.006 (0.002, 0.012)] and the same probability of chronic symptoms in undetected symptomatic infections as detected symptomatic infections, with "detected" assumed equivalent to "medically attended" (outpatient care for mild/moderate and inpatient hospitalisation for severe) (base-case: no chronic symptoms). Sensitivity analysis "infection detection rate" used estimates with lower rate of detection of infection in healthcare system [0.36 vs. 0.63], which impacts on the subsequent parameters including probability of detected mild symptoms, hospitalization and chronic symptoms. DALY = disability-adjusted life-year, K = thousand, M = million.

***Table S11. Sensitivity analyses of projected cumulative total economic costs averted due to CHIKV vaccination from 2025 to 2050, depending on the vaccination campaign.***

| **Outcome** | **Outcome averted (95%UI) by different vaccination campaign scenario** | | | | | | | |
|---|---|---|---|---|---|---|---|---|
| | **Type** | **Population wide** | | | **Routine** | **Population-wide + Routine (P+R)** | | |
| | **Age** | **12-99** | | | **12** | **P: 12-99; R: 12** | | |
| | **Vaccination period** | **2025-2029** | | | **2025-2040** | **P: 2025-2029; R: 2030-2040** | | |
| | **Annual target coverage^** | **2%** | **4%** | **10%** | **65%** | **P: 2%; R: 65%** | **P: 4%; R: 65%** | **P: 10%; R: 65%** |
| Healthcare costs (I$ 2023) | Base-case | 45.5M (41.2M, 49.7M) | 91.0M (82.5M, 99.3M) | 228M (206M, 248M) | 64.7M (54.9M, 74.5M) | 84.8M (76.7M, 94.2M) | 130M (118M, 143M) | 267M (242M, 292M) |
| | SA1: Symptomatic probability | 16.0M (13.9M, 18.3M) | 32.1M (27.7M, 36.7M) | 80.1M (69.3M, 91.6M) | 23.1M (17.6M, 29.6M) | 30.1M (25.4M, 35.5M) | 46.1M (39.7M, 53.5M) | 94.2M (81.9M, 108M) |
| | SA2: Symptomatic distribution | 44.0M (39.0M, 48.7M) | 88.0M (78.1M, 97.4M) | 220M (195M, 243M) | 63.3M (51.4M, 76.3M) | 82.4M (71.9M, 93.1M) | 126M (112M, 141M) | 258M (230M, 286M) |
| | SA3: Disability weight | 45.5M (41.2M, 49.7M) | 91.0M (82.5M, 99.3M) | 228M (206M, 248M) | 64.7M (54.9M, 74.5M) | 84.8M (76.7M, 94.2M) | 130M (118M, 143M) | 267M (242M, 292M) |
| | SA4: Undetected = detected mild | 98.7M (83.9M, 117M) | 197M (168M, 235M) | 494M (419M, 587M) | 145M (117M, 180M) | 187M (157M, 225M) | 285M (241M, 343M) | 582M (494M, 695M) |
| | SA5: Infection detection rate | 26.0M (23.7M, 28.5M) | 52.0M (47.4M, 57.1M) | 130M (118M, 143M) | 37.0M (31.3M, 42.5M) | 48.5M (43.5M, 53.6M) | 74.5M (67.4M, 82.0M) | 153M (139M, 167M) |
| Catastrophic healthcare expenditure (n) | Base-case | 8.92K (8.08K, 9.72K) | 17.8K (16.2K, 19.4K) | 44.6K (40.4K, 48.6K) | 10.4K (9.03K, 11.8K) | 16.3K (14.7K, 17.8K) | 25.2K (22.9K, 27.4K) | 52.0K (47.3K, 56.5K) |
| | SA1: Symptomatic probability | 3.15K (2.70K, 3.66K) | 6.29K (5.40K, 7.32K) | 15.7K (13.5K, 18.3K) | 3.74K (2.98K, 4.66K) | 5.79K (4.94K, 6.78K) | 8.93K (7.66K, 10.4K) | 18.4K (15.7K, 21.3K) |
| | SA2: Symptomatic distribution | 8.68K (7.74K, 9.58K) | 17.4K (15.5K, 19.2K) | 43.4K (38.7K, 47.9K) | 11.2K (9.33K, 13.0K) | 16.6K (14.5K, 18.5K) | 25.3K (22.3K, 28.1K) | 51.3K (45.6K, 56.7K) |
| | SA3: Disability weight | 8.92K (8.08K, 9.72K) | 17.8K (16.2K, 19.4K) | 44.6K (40.4K, 48.6K) | 10.4K (9.03K, 11.8K) | 16.3K (14.7K, 17.8K) | 25.2K (22.9K, 27.4K) | 52.0K (47.3K, 56.5K) |
| | SA4: Undetected = detected mild | 8.92K (8.08K, 9.72K) | 17.8K (16.2K, 19.4K) | 44.6K (40.4K, 48.6K) | 10.4K (9.03K, 11.8K) | 16.3K (14.7K, 17.8K) | 25.2K (22.9K, 27.4K) | 52.0K (47.3K, 56.5K) |
| | SA5: Infection detection rate | 5.41K (4.89K, 5.90K) | 10.8K (9.79K, 11.8K) | 27.1K (24.5K, 29.5K) | 6.43K (5.52K, 7.30K) | 9.96K (8.96K, 10.9K) | 15.4K (13.9K, 16.8K) | 31.6K (28.6K, 34.5K) |
| | Base-case | 8.43K (7.45K, 9.44K) | 16.9K (14.9K, 18.9K) | 42.1K (37.2K, 47.2K) | 7.12K (6.00K, 8.34K) | 13.3K (11.7K, 14.9K) | 21.7K (19.1K, 24.2K) | 47.0K (41.5K, 52.4K) |

| Outcome | Outcome averted (95%UI) by different vaccination campaign scenario | | | | | | | |
|---|---|---|---|---|---|---|---|---|
| | Type | Population wide | | | Routine | Population-wide + Routine (P+R) | | |
| | Age | 12-99 | | | 12 | P: 12-99; R: 12 | | |
| | Vaccination period | 2025-2029 | | | 2025-2040 | P: 2025-2029; R: 2030-2040 | | |
| | Annual target coverage^ | 2% | 4% | 10% | 65% | P: 2%; R: 65% | P: 4%; R: 65% | P: 10%; R: 65% |
| Impoverishing healthcare expenditure (n) | SA1: Symptomatic probability | 2.98K (2.52K, 3.52K) | 5.96K (5.05K, 7.04K) | 14.9K (12.6K, 17.6K) | 2.56K (2.02K, 3.25K) | 4.72K (3.97K, 5.65K) | 7.69K (6.48K, 9.12K) | 16.6K (14.1K, 19.6K) |
| | SA2: Symptomatic distribution | 8.24K (7.17K, 9.28K) | 16.5K (14.3K, 18.6K) | 41.2K (35.8K, 46.4K) | 7.83K (6.35K, 9.24K) | 13.5K (11.7K, 15.4K) | 21.8K (18.9K, 24.7K) | 46.5K (40.5K, 52.5K) |
| | SA3: Disability weight | 8.43K (7.45K, 9.44K) | 16.9K (14.9K, 18.9K) | 42.1K (37.2K, 47.2K) | 7.12K (6.00K, 8.34K) | 13.3K (11.7K, 14.9K) | 21.7K (19.1K, 24.2K) | 47.0K (41.5K, 52.4K) |
| | SA4: Undetected = detected mild | 8.43K (7.45K, 9.44K) | 16.9K (14.9K, 18.9K) | 42.1K (37.2K, 47.2K) | 7.12K (6.00K, 8.34K) | 13.3K (11.7K, 14.9K) | 21.7K (19.1K, 24.2K) | 47.0K (41.5K, 52.4K) |
| | SA5: Infection detection rate | 6.21K (5.38K, 7.17K) | 12.4K (10.8K, 14.3K) | 31.0K (26.9K, 35.8K) | 5.24K (4.32K, 6.23K) | 9.76K (8.43K, 11.3K) | 16.0K (13.8K, 18.4K) | 34.6K (30.0K, 40.0K) |
| Productivity losses (I$ 2023) | Base-case | 1.66B (1.11B, 2.20B) | 3.32B (2.22B, 4.41B) | 8.31B (5.56B, 11.0B) | 1.08B (731M, 1.47B) | 2.32B (1.58B, 3.07B) | 3.98B (2.70B, 5.26B) | 8.97B (6.02B, 11.9B) |
| | SA1: Symptomatic probability | 596M (402M, 818M) | 1.19B (805M, 1.64B) | 2.98B (2.01B, 4.09B) | 389M (252M, 544M) | 832M (572M, 1.14B) | 1.43B (984M, 1.93B) | 3.21B (2.19B, 4.38B) |
| | SA2: Symptomatic distribution | 1.62B (1.10B, 2.18B) | 3.23B (2.20B, 4.37B) | 8.08B (5.49B, 10.9B) | 1.16B (775M, 1.59B) | 2.32B (1.59B, 3.14B) | 3.94B (2.71B, 5.32B) | 8.79B (6.01B, 11.9B) |
| | SA3: Disability weight | 1.66B (1.11B, 2.20B) | 3.32B (2.22B, 4.41B) | 8.31B (5.56B, 11.0B) | 1.08B (731M, 1.47B) | 2.32B (1.58B, 3.07B) | 3.98B (2.70B, 5.26B) | 8.97B (6.02B, 11.9B) |
| | SA4: Undetected = detected mild | 6.03B (4.84B, 7.39B) | 12.1B (9.68B, 14.8B) | 30.2B (24.2B, 37.0B) | 3.11B (2.38B, 3.89B) | 7.92B (6.41B, 9.59B) | 14.0B (11.3B, 17.0B) | 32.1B (25.9B, 39.2B) |
| | SA5: Infection detection rate | 1.37B (855M, 1.90B) | 2.74B (1.71B, 3.80B) | 6.86B (4.28B, 9.49B) | 872M (526M, 1.22B) | 1.90B (1.17B, 2.63B) | 3.27B (2.03B, 4.54B) | 7.39B (4.60B, 10.2B) |
| Monetised DALYs (I$ 2023) | Base-case | 456M (374M, 549M) | 911M (749M, 1.10B) | 2.28B (1.87B, 2.75B) | 510M (387M, 644M) | 765M (632M, 914M) | 1.22B (1.01B, 1.45B) | 2.59B (2.15B, 3.10B) |
| | SA1: Symptomatic probability | 163M (129M, 201M) | 325M (259M, 401M) | 813M (646M, 1.00B) | 183M (130M, 249M) | 273M (220M, 333M) | 436M (353M, 531M) | 923M (747M, 1.13B) |
| | SA2: Symptomatic distribution | 438M (357M, 532M) | 876M (715M, 1.06B) | 2.19B (1.79B, 2.66B) | 504M (382M, 661M) | 743M (606M, 913M) | 1.18B (967M, 1.43B) | 2.49B (2.04B, 3.03B) |

| Outcome | Outcome averted (95%UI) by different vaccination campaign scenario | | | | | | | |
|---|---|---|---|---|---|---|---|---|
| | **Type** | **Population wide** | | | **Routine** | **Population-wide + Routine (P+R)** | | |
| | **Age** | **12-99** | | | **12** | **P: 12-99; R: 12** | | |
| | **Vaccination period** | **2025-2029** | | | **2025-2040** | **P: 2025-2029; R: 2030-2040** | | |
| | **Annual target coverage^** | **2%** | **4%** | **10%** | **65%** | **P: 2%; R: 65%** | **P: 4%; R: 65%** | **P: 10%; R: 65%** |
| | SA3: Disability weight | 628M (503M, 769M) | 1.26B (1.01B, 1.54B) | 3.14B (2.51B, 3.85B) | 685M (517M, 882M) | 1.04B (852M, 1.26B) | 1.67B (1.36B, 2.01B) | 3.55B (2.88B, 4.32B) |
| | SA4: Undetected = detected mild | 3.19B (2.52B, 3.93B) | 6.38B (5.05B, 7.87B) | 15.9B (12.6B, 19.7B) | 3.31B (2.47B, 4.22B) | 5.19B (4.19B, 6.40B) | 8.38B (6.78B, 10.3B) | 18.0B (14.4B, 22.1B) |
| | SA5: Infection detection rate | 262M (216M, 313M) | 524M (433M, 627M) | 1.31B (1.08B, 1.57B) | 294M (228M, 369M) | 440M (368M, 521M) | 702M (588M, 830M) | 1.49B (1.23B, 1.77B) |
| VSL losses (I$ 2023) | Base-case | 3.24B (2.77B, 3.68B) | 6.47B (5.55B, 7.36B) | 16.2B (13.9B, 18.4B) | 4.39B (2.76B, 6.42B) | 5.90B (4.69B, 7.41B) | 9.14B (7.54B, 11.0B) | 18.8B (16.0B, 21.7B) |
| | SA1: Symptomatic probability | 1.16B (964M, 1.39B) | 2.32B (1.93B, 2.79B) | 5.81B (4.82B, 6.97B) | 1.58B (922M, 2.39B) | 2.12B (1.61B, 2.70B) | 3.28B (2.62B, 4.02B) | 6.77B (5.55B, 8.08B) |
| | SA2: Symptomatic distribution | 3.30B (2.74B, 3.89B) | 6.61B (5.47B, 7.78B) | 16.5B (13.7B, 19.5B) | 4.47B (2.74B, 6.57B) | 6.02B (4.69B, 7.51B) | 9.32B (7.57B, 11.2B) | 19.2B (15.9B, 22.6B) |
| | SA3: Disability weight | 3.24B (2.77B, 3.68B) | 6.47B (5.55B, 7.36B) | 16.2B (13.9B, 18.4B) | 4.39B (2.76B, 6.42B) | 5.90B (4.69B, 7.41B) | 9.14B (7.54B, 11.0B) | 18.8B (16.0B, 21.7B) |
| | SA4: Undetected = detected mild | 3.24B (2.77B, 3.68B) | 6.47B (5.55B, 7.36B) | 16.2B (13.9B, 18.4B) | 4.39B (2.76B, 6.42B) | 5.90B (4.69B, 7.41B) | 9.14B (7.54B, 11.0B) | 18.8B (16.0B, 21.7B) |
| | SA5: Infection detection rate | 1.85B (1.61B, 2.12B) | 3.70B (3.22B, 4.23B) | 9.25B (8.06B, 10.6B) | 2.51B (1.55B, 3.55B) | 3.37B (2.64B, 4.17B) | 5.22B (4.32B, 6.21B) | 10.8B (9.26B, 12.5B) |
| VSLY losses (I$ 2023) | Base-case | 2.67B (2.21B, 3.14B) | 5.34B (4.42B, 6.28B) | 13.3B (11.1B, 15.7B) | 7.08B (4.39B, 10.4B) | 6.96B (5.11B, 9.35B) | 9.63B (7.38B, 12.4B) | 17.6B (14.2B, 21.6B) |
| | SA1: Symptomatic probability | 958M (764M, 1.19B) | 1.92B (1.53B, 2.37B) | 4.79B (3.82B, 5.93B) | 2.54B (1.46B, 3.86B) | 2.50B (1.74B, 3.42B) | 3.45B (2.55B, 4.50B) | 6.33B (4.94B, 7.90B) |
| | SA2: Symptomatic distribution | 2.67B (2.17B, 3.19B) | 5.34B (4.34B, 6.38B) | 13.4B (10.9B, 16.0B) | 7.17B (4.34B, 10.7B) | 7.02B (5.01B, 9.38B) | 9.69B (7.28B, 12.4B) | 17.7B (14.0B, 22.1B) |
| | SA3: Disability weight | 2.67B (2.21B, 3.14B) | 5.34B (4.42B, 6.28B) | 13.3B (11.1B, 15.7B) | 7.08B (4.39B, 10.4B) | 6.96B (5.11B, 9.35B) | 9.63B (7.38B, 12.4B) | 17.6B (14.2B, 21.6B) |
| | SA4: Undetected = detected mild | 2.67B (2.21B, 3.14B) | 5.34B (4.42B, 6.28B) | 13.3B (11.1B, 15.7B) | 7.08B (4.39B, 10.4B) | 6.96B (5.11B, 9.35B) | 9.63B (7.38B, 12.4B) | 17.6B (14.2B, 21.6B) |

| Outcome | Outcome averted (95%UI) by different vaccination campaign scenario | | | | | | | |
|---|---|---|---|---|---|---|---|---|
| | **Type** | **Population wide** | | | **Routine** | **Population-wide + Routine (P+R)** | | |
| | **Age** | **12-99** | | | **12** | **P: 12-99; R: 12** | | |
| | **Vaccination period** | **2025-2029** | | | **2025-2040** | **P: 2025-2029; R: 2030-2040** | | |
| | **Annual target coverage^** | **2%** | **4%** | **10%** | **65%** | **P: 2%; R: 65%** | **P: 4%; R: 65%** | **P: 10%; R: 65%** |
| | SA5: Infection detection rate | 1.53B (1.28B, 1.80B) | 3.05B (2.57B, 3.60B) | 7.63B (6.41B, 8.99B) | 4.04B (2.46B, 5.78B) | 3.98B (2.88B, 5.20B) | 5.50B (4.19B, 6.93B) | 10.1B (8.19B, 12.2B) |
| Societal costs (I$ 2023) | Base-case | 2.16B (1.58B, 2.74B) | 4.33B (3.16B, 5.48B) | 10.8B (7.90B, 13.7B) | 1.65B (1.24B, 2.10B) | 3.17B (2.35B, 3.98B) | 5.33B (3.96B, 6.71B) | 11.8B (8.73B, 14.9B) |
| | SA1: Symptomatic probability | 774M (563M, 1.01B) | 1.55B (1.13B, 2.02B) | 3.87B (2.82B, 5.05B) | 595M (416M, 795M) | 1.14B (836M, 1.47B) | 1.91B (1.41B, 2.48B) | 4.23B (3.10B, 5.52B) |
| | SA2: Symptomatic distribution | 2.10B (1.55B, 2.73B) | 4.20B (3.10B, 5.45B) | 10.5B (7.76B, 13.6B) | 1.73B (1.24B, 2.26B) | 3.15B (2.36B, 4.06B) | 5.25B (3.93B, 6.79B) | 11.5B (8.59B, 15.0B) |
| | SA3: Disability weight | 2.34B (1.75B, 2.93B) | 4.67B (3.49B, 5.87B) | 11.7B (8.73B, 14.7B) | 1.83B (1.39B, 2.31B) | 3.45B (2.63B, 4.26B) | 5.78B (4.39B, 7.20B) | 12.8B (9.62B, 16.0B) |
| | SA4: Undetected = detected mild | 9.32B (7.49B, 11.4B) | 18.6B (15.0B, 22.7B) | 46.6B (37.5B, 56.8B) | 6.56B (5.06B, 8.28B) | 13.3B (11.0B, 15.9B) | 22.6B (18.5B, 27.2B) | 50.6B (41.1B, 61.4B) |
| | SA5: Infection detection rate | 1.66B (1.12B, 2.21B) | 3.32B (2.25B, 4.41B) | 8.30B (5.62B, 11.0B) | 1.20B (833M, 1.57B) | 2.39B (1.65B, 3.16B) | 4.05B (2.78B, 5.35B) | 9.03B (6.18B, 12.0B) |

^ Actual achieved coverage = 95% of target coverage, accounting for 5% wastage. All analyses assume 70% vaccine efficacy. Sensitivity analysis for symptomatic infection used estimates with lower probability of symptomatic infection than base-case [Mean (95%CI): 0.180 (0.110, 0.260) vs. 0.505 (0.380, 0.627)]. Sensitivity analysis for symptom age-sex distribution used alternative distribution of symptom by age and sex [supplementary method 2d]. Sensitivity analysis for DW for chronic symptoms used estimates with higher disability weight for chronic chikungunya [Mean (95%CI): 0.331 (0.306, 0.355) vs. 0.229 (0.216, 0.242)]. Sensitivity analysis "undetected = detected mild" used estimates with higher disability weight for undetected symptomatic infections [mean (95CI): 0.110 (0.074, 0.145) vs. 0.006 (0.002, 0.012)] and the same probability of chronic symptoms in undetected symptomatic infections as detected symptomatic infections, with "detected" assumed equivalent to "medically attended" (outpatient care for mild/moderate and inpatient hospitalisation for severe) (base-case: no chronic symptoms). Sensitivity analysis "infection detection rate" used estimates with lower rate of detection of infection in healthcare system [0.36 vs. 0.63], which impacts on the subsequent parameters including probability of detected mild symptoms, hospitalization and chronic symptoms. Future costs and life-years are discounted annually at 3.5%/year. DALY = disability-adjusted life-year, VSL = value of statistical life, VSLY = value of statistical life-years, I$ = International dollar, K = thousand, M = million, B = billion, T= trillion. Venezuela was excluded from the analyses for the economic impact because there is no reliable estimate of economic parameter available in the country due to its economic collapse in the recent years.

***Table S12. Threshold vaccination costs across all 31 countries under different vaccine efficacy assumptions and various vaccination campaign scenarios.***

| | Threshold vaccination costs (95% UI) by different vaccination campaign strategies, I$ 2023 | | | | |
|---|---|---|---|---|---|
| **Type** | **Population wide** | **Routine** | **Population-wide + Routine (P+R)** | | |
| **Age** | **12-99** | **12** | **P: 12-99; R: 12** | | |
| **Vaccination period** | **2025-2029** | **2025-2040** | **P: 2025-2029; R: 2030-2040** | | |
| **Annual target coverage^** | **2%-10%** | **65%** | **P: 2%; R: 65%** | **P: 4%; R: 65%** | **P: 10%; R: 65%** |
| Vaccine efficacy: 90% | 12.7 (9.30, 16.1) | 5.35 (4.01, 6.81) | 8.76 (6.50, 11.0) | 10.0 (7.44, 12.6) | 11.3 (8.38, 14.3) |
| Vaccine efficacy: 70% | 9.90 (7.23, 12.5) | 4.16 (3.12, 5.30) | 6.81 (5.05, 8.55) | 7.80 (5.79, 9.81) | 8.83 (6.52, 11.1) |
| Vaccine efficacy: 50% | 7.07 (5.16, 8.95) | 2.97 (2.22, 3.78) | 4.86 (3.61, 6.10) | 5.57 (4.13, 7.00) | 6.30 (4.65, 7.94) |

^ Actual achieved coverage = 95% of target coverage, accounting for 5% wastage. Threshold vaccination costs was estimated by dividing the cumulative total societal costs over averted by vaccination 2025-2050 across 31 countries by the number of doses required across 31 countries. Venezuela was excluded from the analyses for the threshold vaccination costs because there is no reliable estimate of economic parameter available in the country due to its economic collapse in the recent years. Future costs are discounted annually at 3.5%/year. I$ = International dollar,

***Table S13. Sensitivity analyses of threshold vaccination costs across all 31 countries under various vaccination campaign scenarios.***

| | Threshold vaccination costs (95% UI) by different vaccination campaign strategies, I$ 2023 | | | | |
|---|---|---|---|---|---|
| **Type** | **Population wide** | **Routine** | **Population-wide + Routine (P+R)** | | |
| **Age** | **12-99** | **12** | **P: 12-99; R: 12** | | |
| **Vaccination period** | **2025-2029** | **2025-2040** | **P: 2025-2029; R: 2030-2040** | | |
| **Annual target coverage^** | **2%-10%** | **65%** | **P: 2%; R: 65%** | **P: 4%; R: 65%** | **P: 10%; R: 65%** |
| Base-case | 9.90 (7.23, 12.5) | 4.16 (3.12, 5.30) | 6.81 (5.05, 8.55) | 7.80 (5.79, 9.81) | 8.83 (6.52, 11.1) |
| SA1: Symptomatic probability | 3.54 (2.58, 4.62) | 1.50 (1.05, 2.00) | 2.44 (1.80, 3.15) | 2.79 (2.06, 3.62) | 3.16 (2.32, 4.12) |
| SA2: Symptomatic distribution | 9.60 (7.10, 12.5) | 4.35 (3.12, 5.68) | 6.77 (5.08, 8.72) | 7.67 (5.75, 9.93) | 8.62 (6.41, 11.2) |
| SA3: Disability weight | 10.7 (7.99, 13.4) | 4.60 (3.49, 5.80) | 7.41 (5.65, 9.16) | 8.45 (6.43, 10.5) | 9.55 (7.18, 11.9) |
| SA4: Undetected = detected mild | 42.6 (34.3, 52.0) | 16.5 (12.7, 20.8) | 28.6 (23.5, 34.3) | 33.1 (27.0, 39.8) | 37.8 (30.6, 45.8) |
| SA5: Infection detection rate | 7.59 (5.14, 10.1) | 3.03 (2.10, 3.96) | 5.14 (3.55, 6.79) | 5.92 (4.07, 7.82) | 6.74 (4.61, 8.95) |
| SA6: No discounting for future costs | 13.0 (9.67, 16.3) | 8.01 (6.00, 10.2) | 10.7 (8.18, 13.2) | 11.4 (8.63, 14.1) | 12.2 (9.19, 15.2) |

^ Actual achieved coverage = 95% of target coverage, accounting for 5% wastage. Threshold vaccination costs was estimated by dividing the cumulative total societal costs over averted by vaccination 2025-2050 across 31 countries by the number of doses required across 31 countries. All analyses assume 70% vaccine efficacy. Sensitivity analysis for symptomatic infection used estimates with lower probability of symptomatic infection than base-case [Mean (95%CI): 0.180 (0.110, 0.260) vs. 0.505 (0.380, 0.627)]. Sensitivity analysis for symptom age-sex distribution used alternative distribution of symptom by age and sex [supplementary method 2d]. Sensitivity analysis for DW for chronic symptoms used estimates with higher disability weight for chronic chikungunya [Mean (95%CI): 0.331 (0.306, 0.355) vs. 0.229 (0.216, 0.242)]. Sensitivity analysis “undetected = detected mild” used estimates with higher disability weight for undetected symptomatic infections [mean (95CI): 0.110 (0.074, 0.145) vs. 0.006 (0.002, 0.012)] and the same probability of chronic symptoms in undetected symptomatic infections as detected symptomatic infections, with “detected” assumed equivalent to “medically attended” (outpatient care for mild/moderate and inpatient hospitalisation for severe) (base-case: no chronic symptoms). Sensitivity analysis “infection detection rate” used estimates with lower rate of detection of infection in healthcare system [0.36 vs. 0.63], which impacts on the subsequent parameters including probability of detected mild symptoms, hospitalization and chronic symptoms. Venezuela was excluded from the analyses for the threshold vaccination costs because there is no reliable estimate of economic parameter available in the country due to its economic collapse in the recent years. DALY = disability-adjusted life-year, I$ = International dollar, VSLY = value of statistical life-years

### 2. SUPPLEMENTARY METHODS - MODEL CALIBRATION AND PARAMETERISATION

#### *2a. Suitability estimation for vectors of chikungunya virus*

The distribution surfaces for both major known mosquito vectors of CHIKV, *Aedes aegypti* and *Aedes albopictus*, were produced using both an ensemble of Boosted Regression Trees (BRTs) and random forest (RF) models.

Vector modelling followed the guidance set out in the ECDC technical report by Wint *et al*[4]. Covariates were selected from a covariate suite compiled by the H2020 MOOD project (Table SS1) (https://app.mood-h2020.eu/core). The models were run with the 10 best covariates which were identified by first running models with all covariates. Final covariates consisted of indices of vegetation greenness, day and night land surface temperature and middle infrared derived from a 2012-2021 timeseries of remotely sensed VIIRS imagery, as well as land use, elevation and human population. Vector training data were obtained from the VectorNet project (https://www.ecdc.europa.eu/en/about-us/partnerships-and-networks/disease-and-laboratory-networks/vector-net), combined with data extracted from the Global Biodiversity Information Facility (www.gbif.org). These data were aggregated to a nominal 10km resolution (0.01 degree). Absences were assigned to areas outside the temperature niche (Brady *et al.*[5] complemented by environmental limits for rainfall, temperature, elevation and land cover). The combined dataset was then balanced provide 10000 presence and absence points.

The vector modelling was then performed using both RF and BRT, implemented through the VECMAP® Software Suite (AVIA-GIS, Belgium), to model presence/absence of both species. The BRT learning rate parameter was adjusted to produce 1000 trees, with the Bernoulli variant that is appropriate for presence absence modelling. Ten models were run for each method, each with a 25% holdback. The results from each method were combined to produce ensemble predictions of each metric. The combination of methods tends to compensate for a tendency of BRT to overfit and RF to smooth. Both vector models were produced at a 5-kilometre spatial resolution to allow incorporation of these surfaces as covariates.

Table SS1. Covariate suite available to suitability vector & incidence modelling procedures

| | |
|---|---|
| 1 VCC1103A05: Middle infra-red mean | 43 WGC1115A25: EVI amplitude 2 |
| 2 WGC1103A15: Middle infra-red amplitude 1 | 44 WGC1115A35: EVI amplitude 3 |
| 3 WGC1103A25: Middle infra-red amplitude 2 | 45 WGC1115MN5: EVI minimum |
| 4 WGC1103A35: Middle infra-red amplitude 3 | 46 WGC1115MX5: EVI maximum |
| 5 WGC1103MN5: Middle infra-red minimum | 47 WGC1115P15: EVI phase 1 |
| 6 WGC1103MX5: Middle infra-red maximum | 48 WGC1115P25: EVI phase 2 |
| 7 WGC1103P15: Middle infra-red phase 1 | 49 WGC1115P35: EVI phase 3 |
| 8 WGC1103P25: Middle infra-red phase 2 | 50 WGC1115VR5: EVI variance |
| 9 WGC1103P35: Middle infra-red phase 3 | 51 MI30GRDP1K DEM (Elevation) +1000 |
| 10 WGC1103VR5: Middle infra-red variance | 52 WD1920A05: ERA5 Precipitation mean |
| 11 WGC1107A05: Daytime LST mean | 53 WD1920A15: ERA5 Precipitation amplitude 1 |
| 12 WGC1107A15: Daytime LST amplitude 1 | 54 WD1920A25: ERA5 Precipitation amplitude 2 |
| 13 WGC1107A25: Daytime LST amplitude 2 | 55 WD1920A35: ERA5 Precipitation amplitude 3 |
| 14 WGC1107A35: Daytime LST amplitude 3 | 56 WD1920MN5: ERA5 Precipitation minimum |
| 15 WGC1107MN5: Daytime LST minimum | 57 WD1920MX5: ERA5 Precipitation maximum |
| 16 WGC1107MX5: Daytime LST maximum | 58 WD1920P15: ERA5 Precipitation phase 1 |
| 17 WGC1107P15: Daytime LST phase 1 | 59 WD1920P25: ERA5 Precipitation phase 2 |
| 18 WGC1107P25: Daytime LST phase 2 | 60 WD1920P35: ERA5 Precipitation phase 3 |
| 19 WGC1107P35: Daytime LST phase 3 | 61 WD1920VR5: ERA5 Precipitation variance |
| 20 WGC1107VR5: Daytime LST variance | 62 VCWPOPPPP5: Worldpop Human Population density 2020 |
| 21 WGC1108A05: Nighttime LST mean | 63 VCV59EL5005: GNTED Elevation + 500 |
| 22 WGC1108A15: Nighttime LST amplitude 1 | 64 EELCBARE5: consensus % bare ground |
| 23 WGC1108A25: Nighttime LST amplitude 2 | 65 EELCDCBD35: consensus % deciduous broadleaved forest |
| 24 WGC1108A35: Nighttime LST amplitude 3 | 66 EELCEVGBD25: consensus % evergreen broadleaved forest |
| 25 WGC1108MN5: Nighttime LST minimum | 67 EELCEVGND15: consensus % evergreen needleleaved forest |
| 26 WGC1108MX5: Nighttime LST maximum | 68 EELCFLOOD85: consensus % flooded |
| 27 WGC1108P15: Nighttime LST phase 1 | 69 EELCHB5: consensus % herbaceous cover |
| 28 WGC1108P25: Nighttime LST phase 2 | 70 EELCMANAG75: consensus % managed land |
| 29 WGC1108P35: Nighttime LST phase 3 | 71 EELCOTHTR45: consensus % other land cover |

| | |
|---|---|
| 30 WGC1108VR5: Nighttime LST variance<br>31 WGC1114A05: NDVI mean<br>32 WGC1114A15: NDVI amplitude 1<br>33 WGC1114A25: NDVI amplitude 2<br>34 WGC1114A35: NDVI amplitude 3<br>35 WGC1114MN5: NDVI minimum<br>36 WGC1114MX5: NDVI maximum<br>37 WGC1114P15: NDVI phase 1<br>38 WGC1114P25: NDVI phase 2<br>39 WGC1114P35: NDVI phase 3<br>40 WGC1114VR5: NDVI variance<br>41 WGC1115A05: EVI mean<br>42 WGC1115A15: EVI amplitude 1 | 72 EELCSHRUB55: consensus % shrub cover<br>73 EELCURB95: consensus % urban<br>74 EELCWATER125: consensus % water<br>75 WD82094A05: Relative Humidity mean<br>76 WD82094A15: Relative Humidity amplitude 1<br>77 WD82094A25: Relative Humidity amplitude 2<br>78 WD82094A35: Relative Humidity amplitude 3<br>79 WD82094MN5: Relative Humidity minimum<br>80 WD82094MX5: Relative Humidity maximum<br>81 WD82094P15: Relative Humidity phase 1<br>82 WD82094P25: Relative Humidity phase 2<br>83 WD82094P35: Relative Humidity phase 3<br>84WD82094VR5: Relative Humidity variance |
| LST = Land Surface Temperature. NDVI Normalised Difference vegetation Index; EVI Enhanced Vegetation Index. DEM Digital Elevation. All files starting with VCC11, VC19 and VC82 are Fourier processed MODIS Satellite Imagery produced by the Environmental Research Group Oxford according to the methods set out in Scharlemann *et. al.* (2008)[6].<br>The Relative Humidity layers were produced as described in n Kraemer *et. al.* (2019)[7] and then Fourier Processed as described above.<br>The Elevation layer was extracted from the GMTED datasets (https://topotools.cr.usgs.gov/gmted_viewer/gmted2010_global_grids.php) and the negative values removed by adding 1000.<br>Population layers derived from layers produced by worldpop (https:// www.worldpop.org/datacatalog/)<br>All Files with VCEELC in file name were derived from the Earthenv consensus land cover data product (https://www.earthenv.org/landcover)<br>All layers extracted and standardised by ERGO for MOOD Horizon 2020 project N° 874850 (https://mood-h202.eu) | |

#### *2b. Suitability estimation for chikungunya virus*

*Chikungunya virus suitability estimation methods*

CHIKV suitability represents the probability of CHIKV occurrence in humans, which necessarily requires presence of at least one of the mosquito vector species. However, vector presence alone does not necessarily indicate CHIKV suitability, as there are areas where the primary species are present but no human infections have ever been recorded. Therefore, we estimated a global CHIKV suitability surface by modelling a relationship between environmental predictor variables, including the estimated distributions of *Ae. Albopictus* and *Ae. aegypti*, and CHIKV occurrence points.

A global occurrence database comprising point (e.g. town or city) or polygon (e.g. county or province) locations of confirmed human CHIKV infection presence was compiled from peer-reviewed literature and ProMed Mail reports from 1952 until July 2023 to reduce spatial uncertainty. An occurrence was defined as one or more reports of human CHIKV infection within a unique location in one year. We excluded polygons greater than 2,500 square kilometres to reduce spatial uncertainty. In total, 466 unique point occurrence records and 1335 unique polygon occurrence records were included after removing duplicates. Subsequently we removed four occurrence points with an estimated temperature suitability of 0 for both *Ae. aegypti* and *Ae. albopictus* (based on Brady *et al*[4]), which were all located in high-altitude areas, likely representing imported cases rather than local transmission.

Pseudo-absences, areas of potentially unsuitable conditions at unsampled locations, were generated and weighted based on a surface of temperature suitability for *Ae. aegypti* (one of the mosquito vectors of CHIKV) from Brady *et al*[5]. Temperature suitability for *Aedes albopictus* was not used for the generation of the pseudo-absences as its distribution includes temperate regions where CHIKV has never been recorded. In order to generate pseudo-absence point locations, 10,000 background points were randomly generated and weighted based in relation to temperature suitability. Using these weights, more absence points were therefore assigned to areas with low temperature suitability.

Six environmental predictor variables were selected as explanatory covariates for CHIKV occurrence and were chosen due to being known or hypothesised to contribute to suitability for human CHIKV occurrence. These included: annual mean precipitation, minimum land surface temperature, maximum annual enhanced vegetation index (EVI), population density, as well as distribution surfaces for both mosquito vectors. Global gridded values for these six covariates at the level of 5km x 5km cells were taken from a covariate data suite compiled by the H2020 MOOD project, and available on the MOOD Platform (https://app.mood-h2020.eu/core).

To establish a relationship between the covariate and the CHIKV occurrence data described above, a BRT model was used. The relationship was established at each point-level occurrence or averaged across each polygon-level occurrence. To increase the robustness of the BRT model's predictions and propagate model uncertainty in further calculations, the BRT model was fitted to 100 separate bootstraps of the data, resulting in 100 distinct BRTs. One 5 km×5 km pixel was randomly selected from within each polygon for each individual BRT in order to account for the environmental uncertainty associated with imprecise geographic data. In order to improve the weighting capacity of each of the 100 BRTs, weightings were applied to the background dataset such that the sum of the weighted background data equalled the weighted sum of the occurrence records. Each of the 100 BRTs predicts environmental suitability on a continuous scale from 0 to 1, with a prediction map then being generated by calculating the mean prediction across all BRTs for each 5 km×5 km pixel. This prediction map was subsequently masked using the temperature suitability for both *Ae. aegypti* and *Ae. albopictus* (based on Brady *et al*[4]), ensuring that CHIKV suitability was set to zero when temperature suitability of both vectors was zero.

Cross-validation was applied to each BRT, whereby 10 subsets of the data comprising 10% of the presence and background observations were assessed based on their ability to predict the distribution of the other 90% of records using the mean area under the curve (AUC) statistic. This AUC value was then averaged across the 10 sub-models and finally across all 100 BRTs in the ensemble in order to derive an overall estimate of goodness-of-fit. Additionally, to avoid AUC inflation due to spatial sorting bias, a pairwise distance sampling procedure was used, resulting in a final AUC which is lower than would be returned by standard procedures but which gives a more realistic quantification of the model's ability to extrapolate predictions to new regions.

*Suitability estimation results*

The mean predicted environmental suitability for CHIKV occurrence based on the ensemble of 100 BRTs is shown in Fig SS1. This ensemble model had an AUC of 0.87 (95% CI 0.83-0.90). The variables with the highest relative influence were environmental suitability for *Ae. aegypti* (84%, 95% CI 80-87) and population density (13%, 95% CI 10-17). The relative influence of the other environmental covariates was relatively small (upper limit of the 95% CI <4 for all). Masking using temperature suitability of both vectors had negligible influence on the CHIKV suitability (Fig SS1 vs Fig SS2).

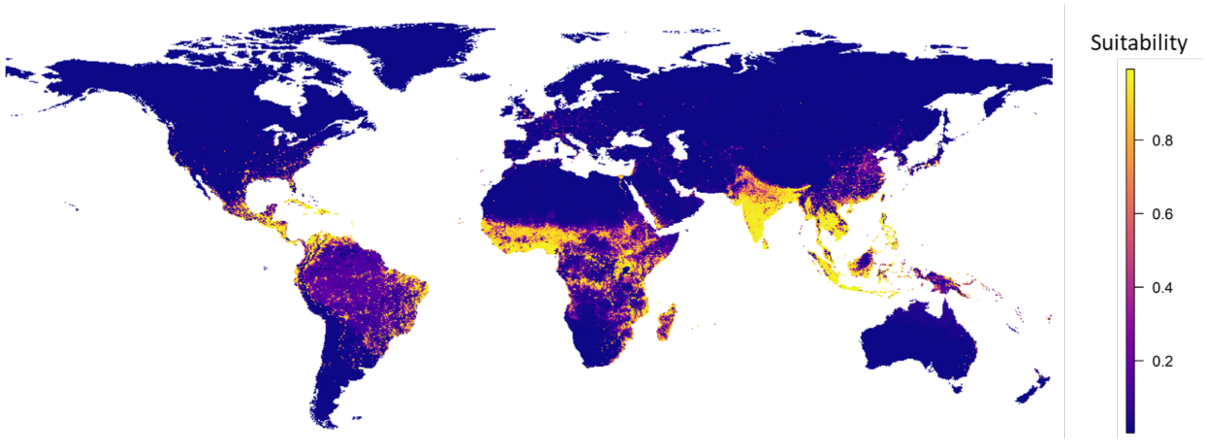

Fig SS1. Mean predicted global chikungunya suitability at the level of 5km-by-5km grid cells across 100 bootstrapped boosted regression trees. Values mapped are predicted probability of CHIKV occurrence.

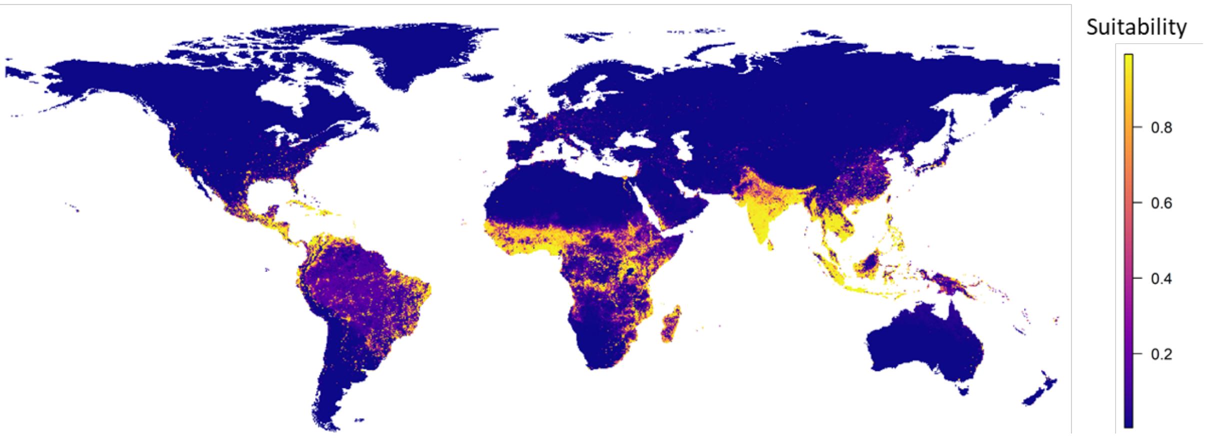

Fig SS2. Predicted suitability for Chikungunya masked by temperature suitability of vectors

*Estimated suitability in the countries of interest*

Suitability was similar across the country in most of the 31 countries and Venezuela in this study (Fig SS3). The variability of the mean estimated suitability across all 5 km x 5 km polygons in the country (excluding those with no population) was smallest in El Salvador, with a median (Q1-Q3) of 0.975 (0.969-0.980), followed by Haiti [0.980 (0.970-0.984)] and Grenada [0.973 (0.949-0.978)]; and largest in Malaysia [0.811 (0.129-0.961)], Indonesia [0.856 (0.251-0.968)] and Colombia [0.177 (0.078-0.789)]. Country-level (weighted by population size) suitability was highest in El Salvador [mean (95%UI): 0.974 (0.971-0.978)], Haiti [0.966 (0.964-0.969)], and Cambodia [0.965 (0.957-0.970)], and lowest in Ethiopia [0.327 (0.292-0.361)], Bolivia [0.335 (0.325-0.346)] and Ecuador [0.539 (0.522-0.550)] (Table SS2). Lim *et al.*[8] also estimated global CHIKV suitability. They used a joint ecological niche modelling framework for multiple *Aedes*-borne viruses (dengue, chikungunya, Zika, and yellow fever) combined with a separate surveillance model to account for spatial reporting bias, which requires additional assumptions and dependencies between diseases than our disease specific approach. While they estimated generally lower country-specific suitability, the ranking of country-specific suitability across countries was similar (Table SS2).

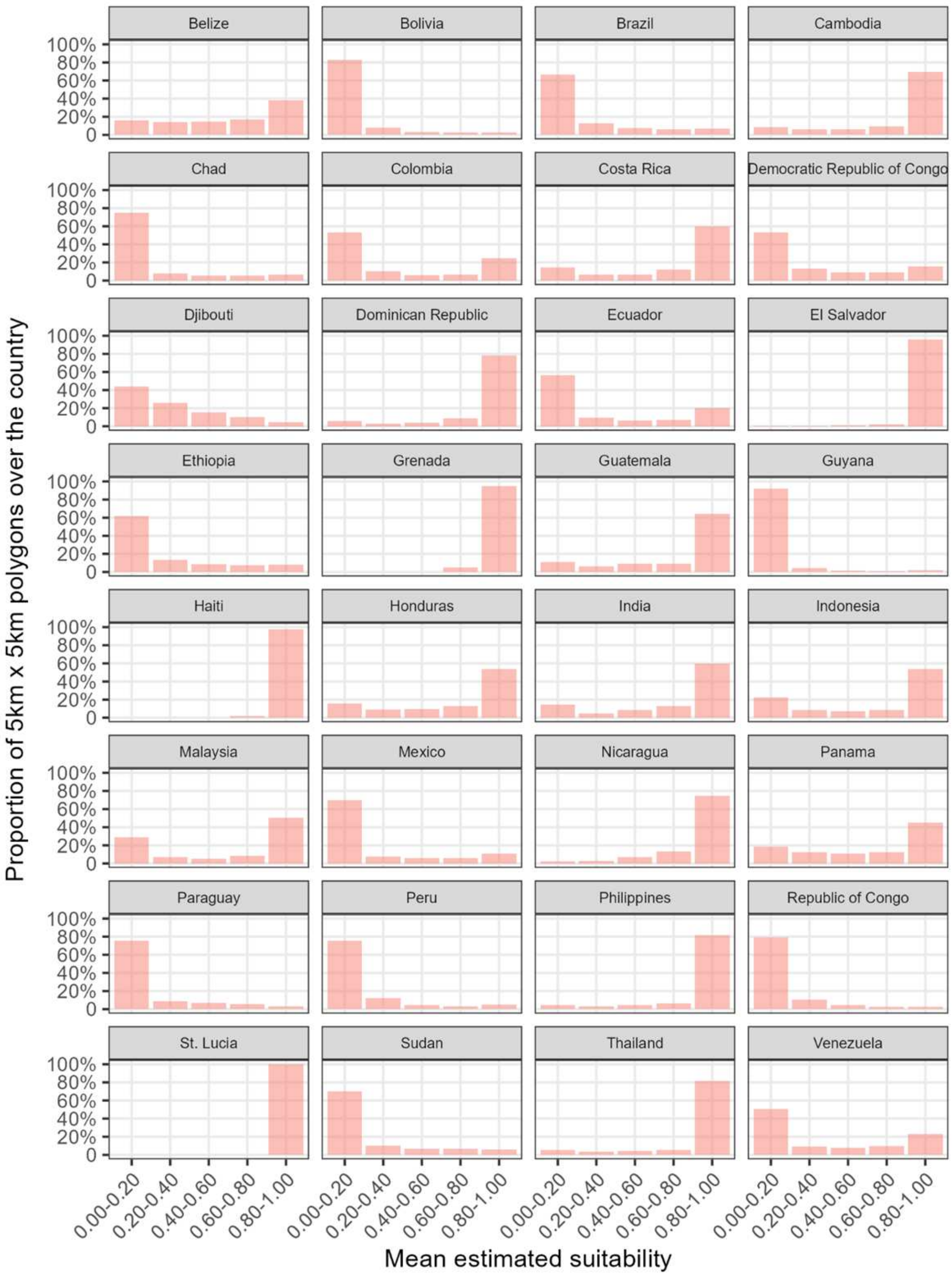

Fig SS3. Distribution of mean suitability in all 5km x 5km polygons over the country.

Table SS2. Country-specific suitability weighted by population size in 31 countries and Venezuela.

| **Country** | **Country-specific suitability weighted by population size** | | | |
|---|---|---|---|---|
| | **Mean** | **Lower 95%UI** | **Upper 95%UI** | **Mean suitability from Lim *et al.*** [8] |
| **El Salvador** | 0.974 | 0.971 | 0.978 | 0.853 |
| **Haiti** | 0.966 | 0.964 | 0.969 | 0.721 |

| Country | Country-specific suitability weighted by population size | | | |
|---|---|---|---|---|
| | Mean | Lower 95%UI | Upper 95%UI | Mean suitability from Lim *et al.* [8] |
| **Cambodia** | 0.965 | 0.957 | 0.970 | 0.565 |
| **Dominican Republic** | 0.964 | 0.961 | 0.967 | 0.845 |
| **Thailand** | 0.963 | 0.958 | 0.967 | 0.685 |
| **Malaysia** | 0.953 | 0.949 | 0.958 | 0.791 |
| **St. Lucia** | 0.948 | 0.939 | 0.955 | 0.733 |
| **Indonesia** | 0.943 | 0.937 | 0.948 | 0.731 |
| **Philippines** | 0.936 | 0.933 | 0.939 | 0.751 |
| **Costa Rica** | 0.935 | 0.927 | 0.942 | 0.786 |
| **Nicaragua** | 0.934 | 0.924 | 0.944 | 0.592 |
| **Panama** | 0.930 | 0.922 | 0.938 | 0.727 |
| **Venezuela** | 0.913 | 0.906 | 0.921 | 0.699 |
| **Honduras** | 0.909 | 0.898 | 0.919 | 0.689 |
| **Grenada** | 0.886 | 0.876 | 0.895 | 0.616 |
| **Brazil** | 0.883 | 0.871 | 0.895 | 0.870 |
| **India** | 0.876 | 0.832 | 0.911 | 0.654 |
| **Belize** | 0.841 | 0.804 | 0.871 | 0.415 |
| **Paraguay** | 0.821 | 0.782 | 0.855 | 0.775 |
| **Guyana** | 0.811 | 0.795 | 0.828 | 0.582 |
| **Congo – Democratic Republic** | 0.761 | 0.726 | 0.794 | 0.392 |
| **Guatemala** | 0.733 | 0.719 | 0.747 | 0.642 |
| **Djibouti** | 0.708 | 0.621 | 0.771 | 0.325 |
| **Colombia** | 0.652 | 0.640 | 0.660 | 0.536 |
| **Congo - Republic** | 0.628 | 0.596 | 0.657 | 0.511 |
| **Sudan** | 0.618 | 0.531 | 0.688 | 0.300 |
| **Peru** | 0.614 | 0.607 | 0.621 | 0.547 |
| **Mexico** | 0.585 | 0.577 | 0.595 | 0.733 |
| **Chad** | 0.581 | 0.505 | 0.646 | 0.208 |
| **Ecuador** | 0.539 | 0.522 | 0.550 | 0.473 |
| **Bolivia** | 0.335 | 0.325 | 0.346 | 0.366 |
| **Ethiopia** | 0.327 | 0.292 | 0.361 | 0.226 |

#### *2c. Force of infection estimation*

Force of infection estimates

Force of infection (FoI) was defined as the annual probability of a susceptible individual in a given location becoming infected. Location specific annual long-term average FoI estimates were derived from Kang *et al.*[9], who fitted serocatalytic models to cross-sectional age-stratified seroprevalence datasets, covering 76 locations in 28 countries, including the African Region (AFRO: Benin, Burkina Faso, Cameroon, Comoros, Ethiopia, Gabon, Kenya, Senegal, and Zambia); Eastern Mediterranean Region (EMRO: Djibouti, Iran, Qatar); Region of the Americas (AMRO: Brazil, France-Martinique, Haiti, Mexico, Nicaragua, Puerto Rico, US-Virgin Islands); European Region (EURO: Italy); South-East Asian Region (SEARO: India,

Indonesia, Myanmar, and Thailand); and Western Pacific Region (WPRO: Fiji, Malaysia, Papua New Guinea, and Singapore).

*Mapping force of infection to the other location*

While Kang *et al.*[9] estimated the average annual FoI in locations with age-stratified seroprevalence data, they provided no information about the likely FoI in other locations, including those with known outbreaks or high CHIKV suitability. To predict the average annual FoI in all geographic areas within the 31 countries and Venezuela included in our model, we estimated the relationship between predicted suitability, which is available in all locations based on the BRT model, and the average annual FoIs derived from Kang *et al.*[9].

A logistic growth curve model was used to model the relationship between FoI and environmental CHIKV suitability. The model had the following form:

$$\lambda(x) = \frac{L}{1 + \exp\left(-k(x - x_0)\right)}$$

where $\lambda$ is the estimated annual FoI, $L$ the upper asymptote, $x$ the estimated CHIKV suitability, $x_0$ the inflection point, and $k$ the growth rate. The model was implemented with the following parameter constraints to ensure biologically plausible estimates and model convergence:

$$L \in [0, 1.2 \times \max(\lambda)],$$

$$x_0 \in [\min(x), \max(x)],$$

$$\frac{1}{k} \in [0.01, \max(x) - \min(x)]$$

These constraints reflect the assumption that the relationship between environmental CHIKV suitability and the FoI can be captured by a logistic growth curve: when suitability is low, the risk of infection is negligible, while at higher suitability levels, the risk may increase and eventually plateau once conditions are consistently sufficiently favourable for transmission. This model was subsequently used to predict the potential average annual FoI in all areas of interest using CHIKV suitability estimates derived using the BRTs.

This resulted in predictions of annual FoI for each 5 km x 5 km pixel, which were subsequently aggregated at the country-level by taking the population-weighted average from each pixel. To propagate uncertainty in both suitability and FoI, the logistic growth curve model was fitted on a combination of 100 bootstrapped suitability estimates from the BRTs and 100 posterior draws from the FoI estimates from Kang *et al.*[9] based on their reported mean and 95%CI and assuming the estimate following beta distribution.

With the mean (m) and standard error (se) of the probability estimate, the parameters (α, β) of the Beta distribution [Beta(α, β)] were derived with the following equations

$$\alpha = \frac{m^2(1-m)}{se^2} - m$$

$$\beta = \frac{\alpha(1-m)}{m}$$

In a sensitivity analysis, suitability point estimates were derived from Lim et al.[8] instead to perform the mapping.

*Prediction of force of infection*

Based on the above mapping, we predicted the average annual FoI at the country-level, ranging from 0.006 (95%UI 0.005 - 0.007) in Boliva to 0.015 (95%UI 0.013-0.017) in Cambodia (Table

SS3). In the sensitivity analysis using the suitability surface from Lim *et al.*,[8] similar FoIs were estimated in most countries.

Table SS3. Predicted country-specific annual force of infection weighted by population size in 31 countries and Venezuela.

| Country | Country-specific annual force of infection weighted by population size | | | |
|---|---|---|---|---|
| | **Mean** | **Lower 95%UI** | **Upper 95%UI** | **Mean from the sensitivity analysis using suitability from Lim *et al.* [8]** |
| **Cambodia** | 0.015 | 0.013 | 0.017 | 0.014 |
| **El salvador** | 0.015 | 0.013 | 0.017 | 0.015 |
| **Nicaragua** | 0.015 | 0.013 | 0.017 | 0.014 |
| **Thailand** | 0.015 | 0.013 | 0.017 | 0.014 |
| **Costa rica** | 0.014 | 0.013 | 0.016 | 0.015 |
| **India** | 0.014 | 0.013 | 0.016 | 0.014 |
| **Honduras** | 0.014 | 0.013 | 0.016 | 0.014 |
| **Malaysia** | 0.014 | 0.012 | 0.016 | 0.014 |
| **Indonesia** | 0.014 | 0.012 | 0.016 | 0.013 |
| **Brazil** | 0.014 | 0.012 | 0.016 | 0.015 |
| **Paraguay** | 0.014 | 0.012 | 0.016 | 0.015 |
| **Panama** | 0.014 | 0.012 | 0.016 | 0.013 |
| **Venezuela** | 0.014 | 0.012 | 0.016 | 0.014 |
| **Dominican republic** | 0.014 | 0.012 | 0.016 | 0.014 |
| **Haiti** | 0.013 | 0.012 | 0.015 | 0.013 |
| **Congo - democratic republic** | 0.013 | 0.012 | 0.015 | 0.012 |
| **Belize** | 0.013 | 0.012 | 0.015 | 0.011 |
| **Philippines** | 0.013 | 0.011 | 0.015 | 0.012 |
| **Guyana** | 0.013 | 0.011 | 0.015 | 0.011 |
| **Sudan** | 0.013 | 0.011 | 0.014 | 0.011 |
| **Congo - republic** | 0.012 | 0.011 | 0.014 | 0.013 |
| **Chad** | 0.012 | 0.011 | 0.014 | 0.011 |
| **Saint lucia** | 0.012 | 0.011 | 0.014 | 0.009 |
| **Guatemala** | 0.012 | 0.010 | 0.013 | 0.013 |
| **Grenada** | 0.011 | 0.009 | 0.012 | 0.006 |
| **Djibouti** | 0.010 | 0.009 | 0.012 | 0.008 |
| **Colombia** | 0.010 | 0.009 | 0.012 | 0.011 |
| **Peru** | 0.010 | 0.008 | 0.011 | 0.011 |
| **Mexico** | 0.010 | 0.008 | 0.011 | 0.014 |
| **Ecuador** | 0.009 | 0.008 | 0.010 | 0.009 |
| **Ethiopia** | 0.007 | 0.006 | 0.008 | 0.010 |
| **Bolivia** | 0.006 | 0.005 | 0.007 | 0.009 |

*Validation of estimated force of infection against age-specific seroprevalence data*

To validate our approach for projecting infection incidence globally, we compared CHIKV seroprevalence as estimated by our model in specific locations and timepoints to corresponding reports of observed seroprevalence from the literature in those same locations and timepoints. Using age-specific seroprevalence data synthesised by both Kang *et al.*[9] and Dos Santos *et al.*[10], we identified 69 unique location-years of seroprevalence data from 66 locations (3 locations have two years of seroprevalence data) in 28 countries (multiple locations contributing to the same seroprevalence data were considered as one unique location). We assumed the first year of CHIKV exposure to be 2013 for American countries[11] and to be the first year of exposure as summarized in a review by Wahid *et al.*[12] for non-American countries as below (Table SS4). Calculation of the seroprevalence in the year were explained in detail in section 2c.

Table SS4. First year of exposure to CHIKV in 28 countries where location age-specific seroprevalence data was available for the validation.

| **Country/region** | **First year CHIKV exposure** |
|---|---|
| **American countries** | |
| Brazil, Mexico, Nicaragua, Puerto Rico, US Virgin Islands | 2013 |
| **Non-American countries** | |
| Benin | 2006 |
| Burkina Faso | 1952* |
| Cambodia | 1961 |
| Cameroon | 2008 |
| Comoros | 2006 |
| Djibouti | 1952* |
| Ethiopia | 1952* |
| Fiji | 2012* |
| France - Guadeloupe | 2013 |
| France - Martinique | 2013 |
| Gabon | 2011 |
| India | 1963 |
| Indonesia | 1972 |
| Iran | 2013* |
| Kenya | 1983 |
| Malaysia | 2005 |
| Myanmar | 1973 |
| Papua New Guinea | 2013 |
| Qatar | 2013* |
| Senegal | 1998 |
| Singapore | 2010 |
| Thailand | 1960 |
| Vietnam | 1966 |
| Zambia | 1952* |

*Not reported in Wahid et al[12], based on the earliest first year of exposure in the nearby country (1952 for Burkina Faso, Djibouti, Ethiopia and Zambia assumed the same as Tanzania in Africa; 2013 for Iran and Qatar assumed the same as Saudi Arabia; 2012 for Fiji assumed the same as Papua New Guinea in Oceania/Pacific Islands)

Fig SS4 compares reported age-specific seroprevalence from these 69 location-years to corresponding model predictions. We calculated study-level residuals between observed and predicted seroprevalence, weighted by the inverse of the estimated variance of the local seroprevalence data. Then we calculated the mean residual across studies by averaging study-level residuals weighted by study sample size. The mean residual was -0.076, representing an underestimation of seroprevalence by 7.6 percentage points, and indicating potential underestimation of FoI or misspecification of CHIKV's emergence date in some locations. When using the suitability map from Lim *et al.* [8], the mean residual was estimated to be slightly larger (-0.081).

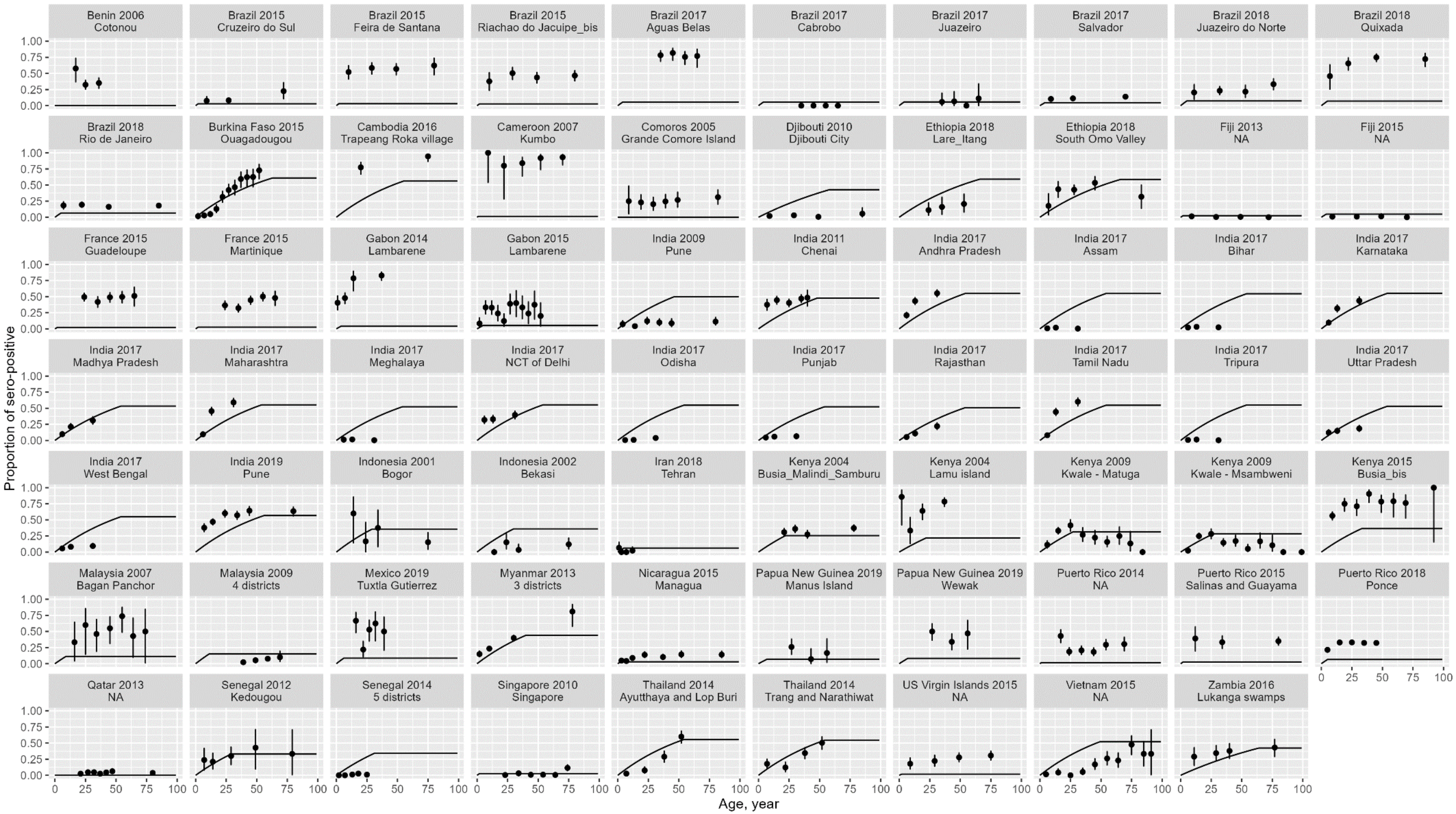

Fig SS4. Estimated age-specific seroprevalence compared with observed age-specific seroprevalence from the data (dot with error bar) in the location and the same year. Line represents the mean estimated seroprevalence; dot and error bar represent the mean and 95%CI of the observed seroprevalence.

#### *2d. Infection projections through time*

As CHIKV antibodies following natural infection provide long-lasting immunity against reinfection[13], infection-induced seropositivity was assumed to be lifelong, and only infection-seronegative individuals were considered at-risk for CHIKV infection. Vaccine-induced immunity was modelled separately and was therefore not influencing the seropositivity of the population. The annual number of infections *Inf* in each country was calculated as, $Inf_{t,age} = \lambda \times N_{t,age} \times (1 - P_{t,age})$

where λ is the country's estimated population-weighted average annual FoI, *N* is its population size at each one-year age group *age* in year *t,* and *P* is its CHIKV seroprevalence (the proportion of the population seropositive due to infection) at each *age* in year *t.*

The annual population sizes *N* at each age year over 2025-2050 were sourced from UN World Population Prospects (medium variant)[14]. For seroprevalence, a recent systematic review and meta-analysis of population-based CHIKV seroprevalence studies by Skalinski *et al.* estimated 31% for AFRO, 29% for AMRO, 5% for EMRO, 24% for SEARO and 18% for WPRO[15]. However, there is no detailed country-specific data on antibody levels, nor data on the age-specific antibody levels. Therefore, we decided to estimate seroprevalence at each age year over 2025-2050 based on the estimated country-level annual average FoI and the assumed first year of CHIKV exposure, accumulating immunity due to infections occurring each year to 2050, while accounting for births and deaths.

First, we calculated the total years of exposure for each age group at each calendar year since the first year of CHIKV exposure. For example, at the first year since CHIKV emergence (beginning of the year), all ages 0-99 had 0-year exposure; at the second year since emergence, all age 1-99 had 1-year exposure; at the third year, all age 2-99 had 2-year exposure, but age 1 had just 1-year exposure (and age 99 in the previous year is assumed to die at age 100). The first year of exposure was assumed to be 2013 for American countries[11], and was based on estimates from the review by Wahid et al[12] for non-American countries (Table SS5).

Table SS5. First year of exposure to CHIKV in 31 included countries and Venezuela.

| **Countries** | **First year CHIKV exposure** |
|---|---|
| **American countries** | |
| Belize, Brazil, Bolivia, Colombia, Costa Rica, Dominican Republic, Ecuador, El Salvador, Grenada, Guatemala, Guyana, Haiti, Honduras, Mexico, Nicaragua, Panama, Paraguay, Peru, St. Lucia, Venezuela | 2013 |
| **Non-American countries** | |
| Chad, Djibouti, Ethiopia | 1952* |
| Thailand | 1960 |
| Cambodia | 1961 |
| India | 1963 |
| Philippines | 1965 |
| Indonesia | 1972 |
| Sudan | 1989 |
| Malaysia | 1998 |
| Democratic Republic of Congo, Republic of Congo | 2011 |

*Not reported in Wahid et al[12], based on the earliest first year of exposure in the nearby country (1952 for Chad, Djibouti, Ethiopia assumed the same as Tanzania in Africa)

Secondly, we calculated seroprevalence for each age year in each calendar year based on the estimated annual FoIs (λ) and calculated total years of CHIKV exposure ($y_{exposure}$), assuming no impact of migration, i.e. equal seroprevalence in each country's immigrating and emigrating populations.

$$P_{y_{exposure}} = 1 - (1 - \lambda)^{y_{exposure}}$$

Seroprevalence by age at 2025 varies by country (Fig SS5). Using either our suitability map or the one from Lim *et al.* [8] led to similar number of infections in most countries (Table SS6). Over 2025-2050, average seroprevalence was predicted to increase (Fig SS6), leading to decreasing rates of infection (Fig SS7), but different trends in the total number of infections due to the trends in population size (Fig SS8).

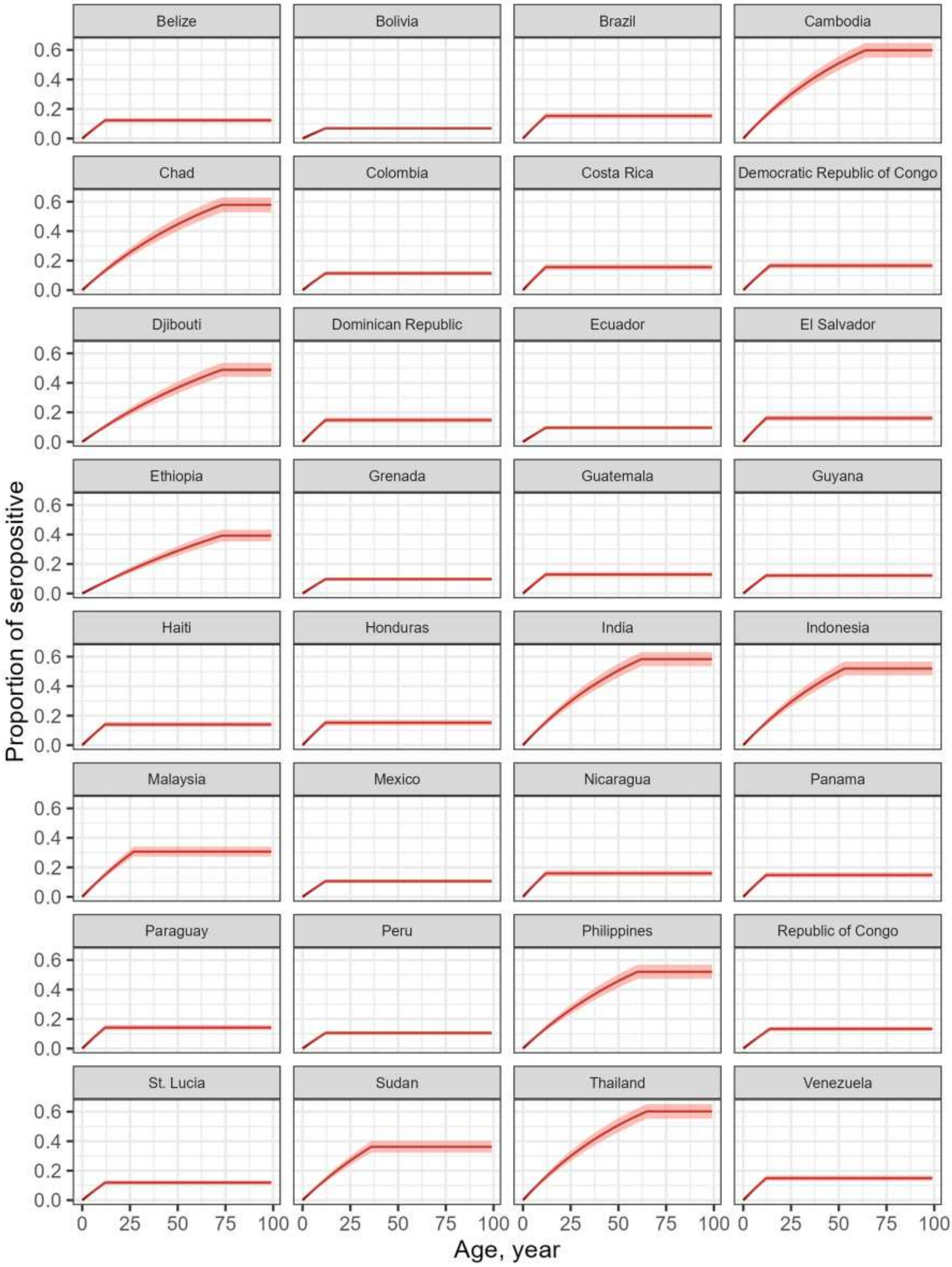

Fig SS5. Estimated age-specific seroprevalence by country in 2025. The range of the shaded area represents the minimum and maximum simulated seroprevalence.

Table SS6. Predicted annual number of infections in 2025 in 31 countries and Venezuela.

| Country | Predicted annual infection, Mean (95%CI) | |
|---|---|---|
| | Base-case | Sensitivity analysis using mean suitability from Lim *et al.* [8] |
| **All** | 27.2M (26.0M, 28.5M) | 28.0M (25.4M, 30.6M) |
| **Belize** | 4.09K (3.61K, 4.58K) | 3.66K (3.23K, 4.09K) |
| **Bolivia** | 69.8K (61.0K, 78.6K) | 96.0K (84.5K, 107K) |
| **Brazil** | 2.50M (2.22M, 2.78M) | 2.63M (2.35M, 2.92M) |
| **Cambodia** | 174K (159K, 189K) | 164K (149K, 178K) |
| **Chad** | 198K (176K, 220K) | 178K (159K, 197K) |
| **Colombia** | 477K (422K, 533K) | 494K (437K, 551K) |
| **Costa Rica** | 61.9K (54.9K, 69.0K) | 63.1K (56.2K, 70.1K) |
| **Democratic Republic of Congo** | 1.27M (1.13M, 1.41M) | 1.17M (1.04M, 1.31M) |
| **Djibouti** | 8.47K (7.57K, 9.38K) | 7.16K (6.40K, 7.91K) |
| **Dominican Republic** | 132K (117K, 147K) | 133K (118K, 148K) |
| **Ecuador** | 139K (122K, 157K) | 152K (134K, 170K) |
| **El Salvador** | 78.4K (69.2K, 87.7K) | 81.4K (72.5K, 90.4K) |
| **Ethiopia** | 790K (696K, 883K) | 1.11M (999K, 1.23M) |
| **Grenada** | 906 (793, 1.02K) | 598 (524, 672) |
| **Guatemala** | 189K (167K, 211K) | 209K (185K, 233K) |
| **Guyana** | 8.01K (7.04K, 8.98K) | 7.49K (6.61K, 8.37K) |
| **Haiti** | 130K (115K, 146K) | 125K (110K, 139K) |
| **Honduras** | 131K (116K, 145K) | 130K (116K, 145K) |
| **India** | 13.7M (12.6M, 14.9M) | 13.7M (12.6M, 14.8M) |
| **Indonesia** | 2.64M (2.42M, 2.86M) | 2.59M (2.37M, 2.81M) |
| **Malaysia** | 363K (327K, 400K) | 361K (327K, 395K) |
| **Mexico** | 1.11M (975K, 1.24M) | 1.59M (1.42M, 1.77M) |
| **Nicaragua** | 85.8K (76.1K, 95.5K) | 81.3K (72.2K, 90.5K) |
| **Panama** | 52.4K (46.4K, 58.4K) | 50.9K (45.2K, 56.7K) |
| **Paraguay** | 77.9K (68.6K, 87.1K) | 82.9K (73.6K, 92.2K) |
| **Peru** | 289K (255K, 324K) | 318K (281K, 355K) |
| **Philippines** | 1.03M (932K, 1.12M) | 1.01M (919K, 1.10M) |
| **Republic of Congo** | 58.5K (51.7K, 65.4K) | 59.7K (52.8K, 66.7K) |
| **St. Lucia** | 1.69K (1.49K, 1.89K) | 1.42K (1.25K, 1.59K) |
| **Sudan** | 508K (454K, 561K) | 473K (424K, 522K) |
| **Thailand** | 601K (560K, 641K) | 590K (550K, 631K) |

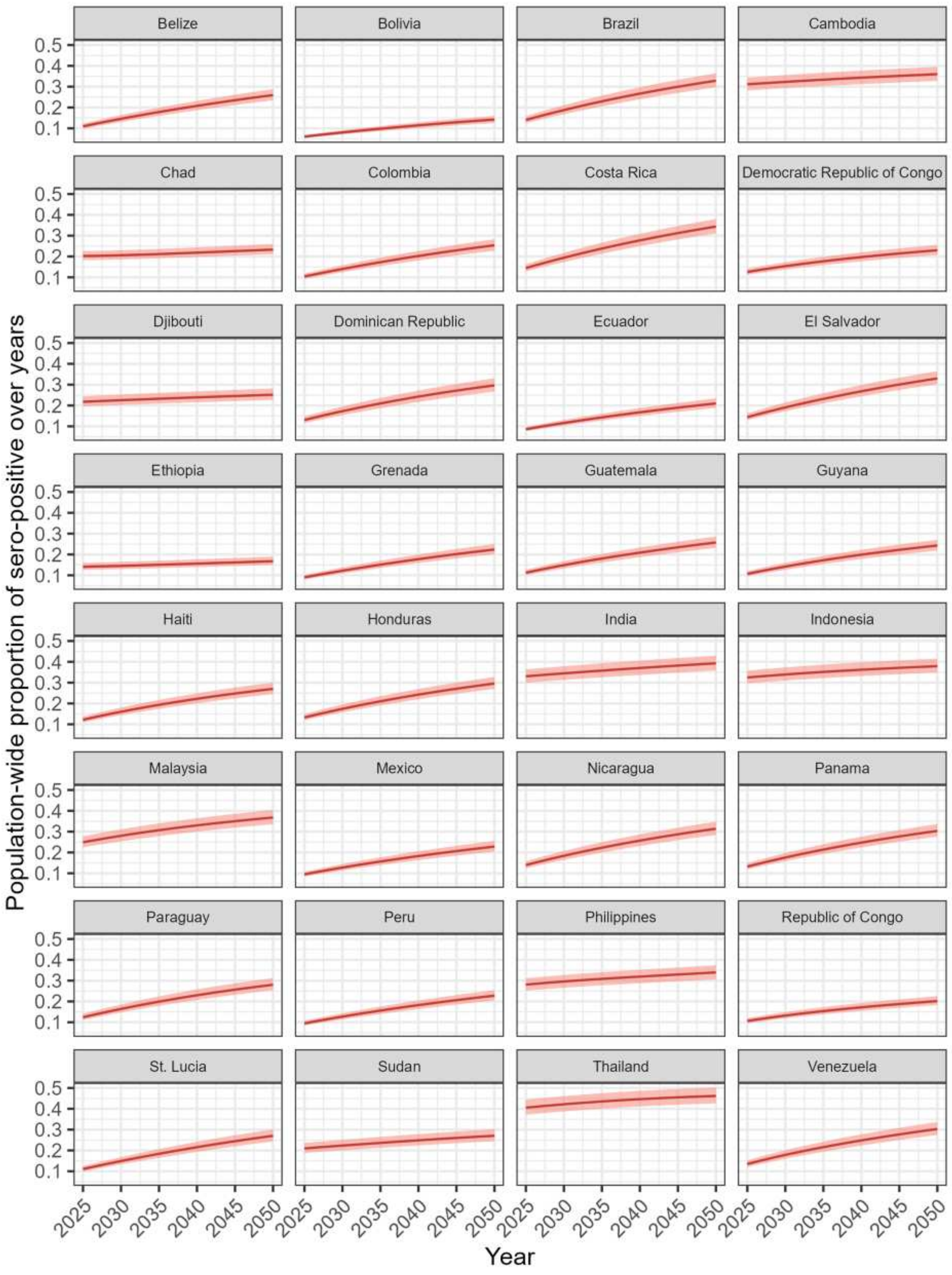

Fig SS6. Estimated population wide seroprevalence by country over 2025-2050. The range of the shaded area represents the minimum and maximum simulated seroprevalence.

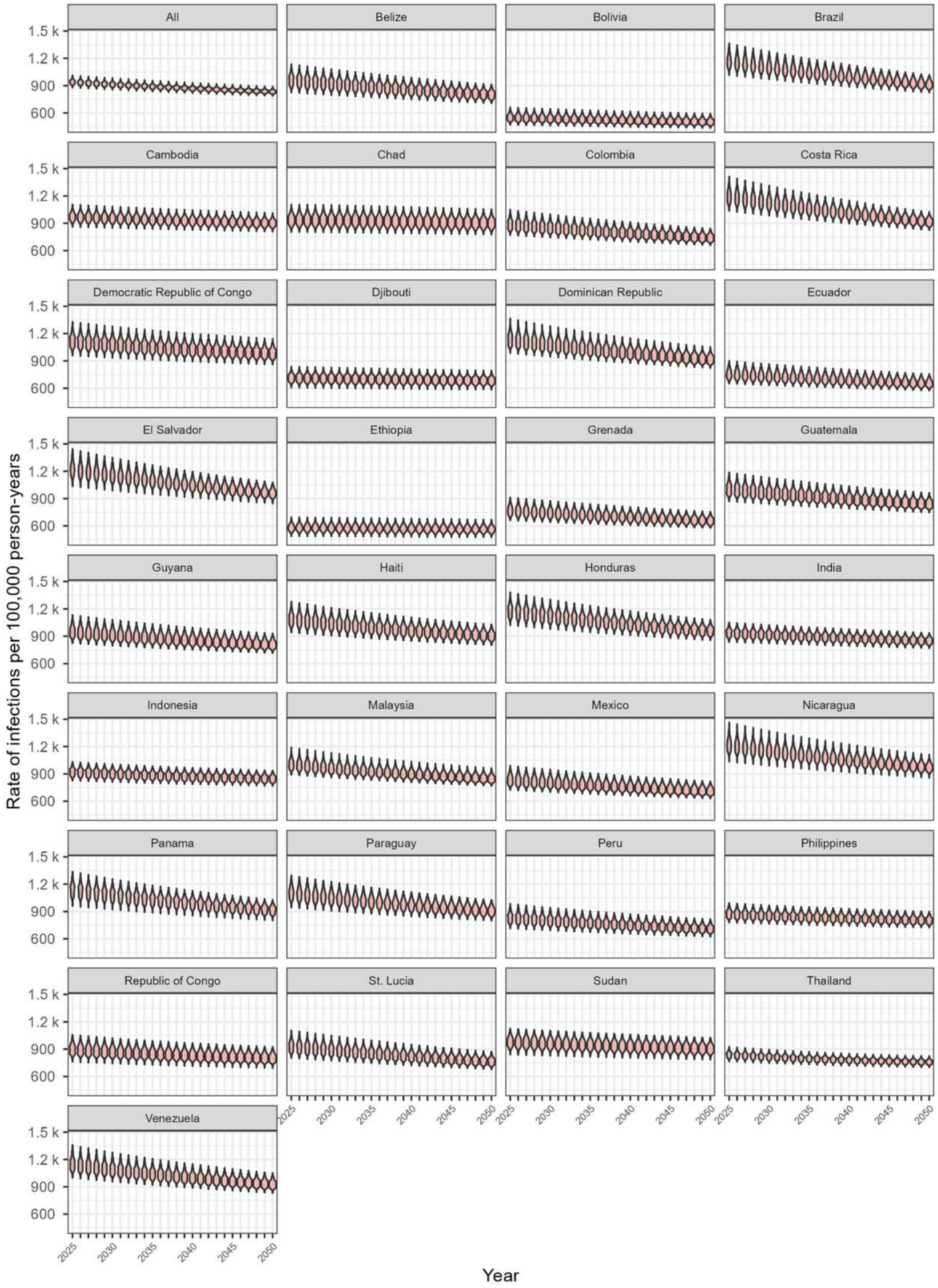

Fig SS7. Rate of infections per 100,000 person-years by country and year across all ages over 2025-2050. The range of the shaded area represents the minimum and maximum simulated rate of infection.

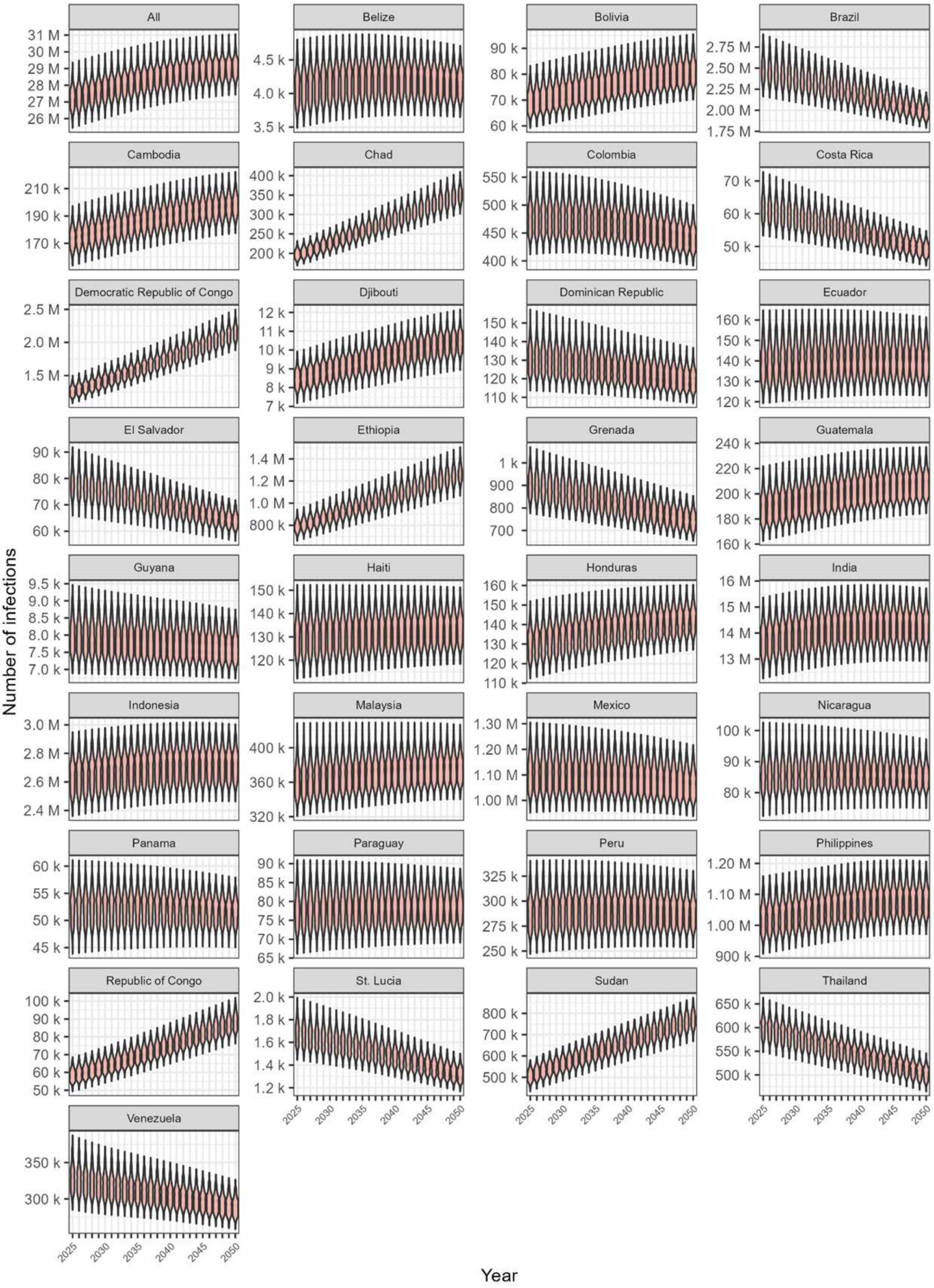

Fig SS8. Total number of infections by country and year across all ages over 2025-2050. The range of the shaded area represents the minimum and maximum simulated number of infections.

***2e. Age- and sex-specific disease risk***

*Probability of symptomatic infection by age and sex*

We estimated the probability of symptomatic infection by age and sex, using (a) the probability of an infected person developing symptoms, (b) the distribution of symptomatic infections by age and sex, and (c) the population size distribution by age and sex in the country from which our input for (b) is taken. Here, (c) is effectively the infection distribution by age and sex, as we assumed equal risk of infection regardless of age and sex. Therefore, the probability of symptomatic infection for a specific age and sex group *i* was derived using (a) × ($b_i$) / ($c_i$).

In the base-case analysis, we used the probability of symptomatic infection derived from Bustos Carillo *et al.* based on the 2015 outbreak in Nicaragua[16], which is in accordance with the report of epidemics from 2014-2015 Colombian outbreaks[17]. We used the distribution of symptomatic infections by age and sex from Sanches Palasio *et al.* based on 635,195 symptomatic infections over 2015-2021 in Brazil[18], which is the study reporting the largest number of symptomatic infections stratified by age and sex. Correspondingly, we used the latest estimate (from the UN World Population Prospects 2024) of the Brazilian population's age and sex distribution to derive probabilities of symptomatic infection by age and sex.

In a sensitivity analysis, we used an alternative estimate of the probability of symptomatic infection derived from Yoon *et al.*, who reported a much lower probability of symptomatic infection based on the 2012-2013 outbreak in Philippines (Table SS6)[19]. In another sensitivity analysis, we used an alternative distribution of symptomatic cases by age and sex from Imad *et al.*, which reported a slightly different distribution based on 1,470 symptomatic infections in the 2019 outbreak in the Maldives[20] (using the latest estimate of population size distribution by age and sex in Maldives to derive probabilities of symptomatic infection by age and sex).

Uncertainty was estimated by applying a Beta distribution to the probability estimates (mean and standard error from the 95%CIs), and applying a Dirichlet distribution to the reported symptomatic infections stratified by age and sex.

With the mean (m) and standard error (se) of the probability estimate, the parameters (α, β) of the Beta distribution [Beta(α, β)] were derived with the following equations

$$\alpha = \frac{m^2(1-m)}{se^2} - m$$

$$\beta = \frac{\alpha(1-m)}{m}$$

Which were derived from the below equations

$$m = \frac{\alpha}{\alpha+\beta}$$

$$se^2 = \frac{\alpha\beta}{(\alpha+\beta)^2(\alpha+\beta+1)}$$

In the base-case analysis, we used 6,352 (i.e. 1% of the original sample) symptomatic infections in the Dirichlet distribution rather than the full count. This was done to avoid overly narrow uncertainty intervals that would arise from the large sample size. We considered that the observed distribution from several years from a single country likely underestimates the true uncertainty about future age- and sex-specific distributions. (Table SS7, Fig SS9)

As the generated probabilities of symptomatic infection in some group by age and sex may include values larger than 1 (proportion of such samples <8%), they were further summarized into mean and SE (Table SS8) to derive the parameters (α, β) of the Beta distribution [Beta(α, β)] to resample the probabilities (all ranging from 0 to 1) for uncertainty consideration.

Table SS7. Probability of symptomatic infection.

| **Probability of symptoms given CHIKV infection** | **Mean (95% CI)** | **Source** |
|---|---|---|
| **Base-case** | 0.505 (0.380, 0.627) | Bustos Carillo *et al.* [16] |
| **Sensitivity analysis** | 0.180 (0.110, 0.260) | Yoon *et al.* [19] |

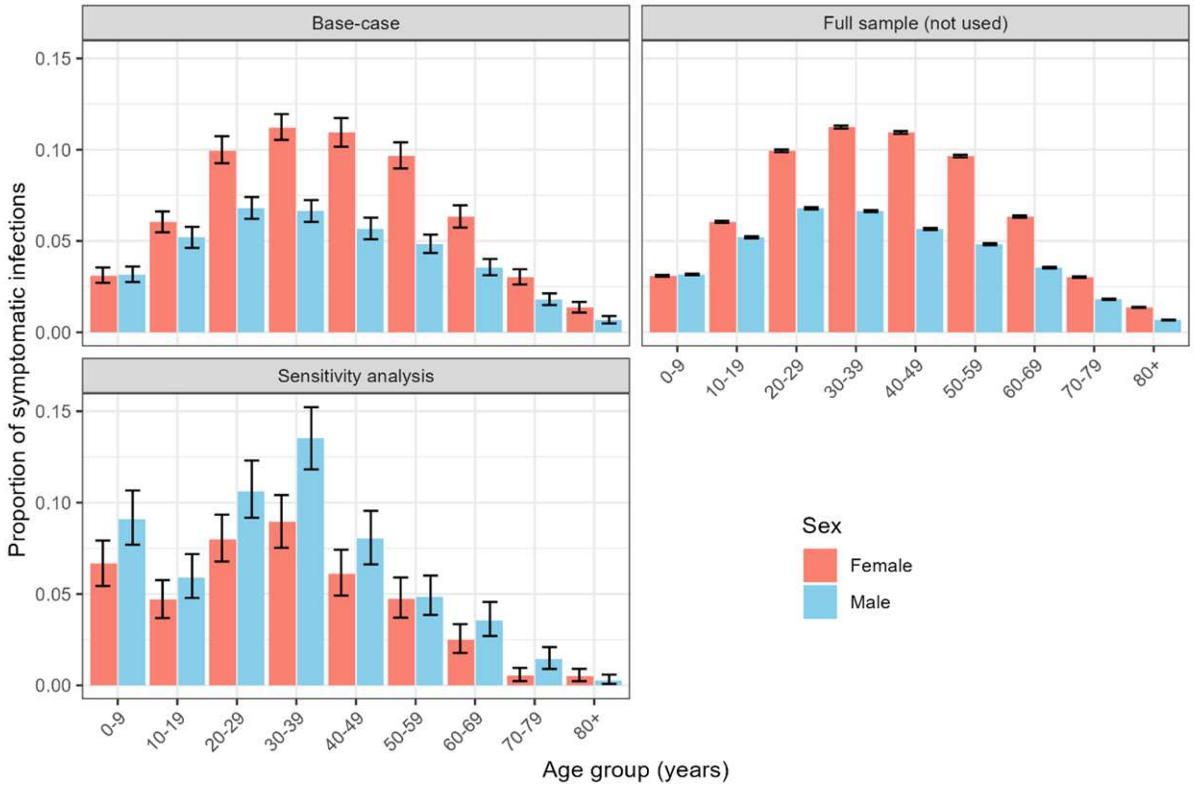

Fig SS9. Proportion of symptomatic infections by age and sex used in the model.

Table SS8. Probability of symptomatic infection by age and sex

| **Age group** | **Mean probability of symptomatic infection given CHIKV infection (SE)** | | | | | |
|---|---|---|---|---|---|---|
| | **Base-case** | | **Sensitivity analysis 1 (probability of symptomatic infection)** | | **Sensitivity analysis 2 (Symptomatic case distribution by age and sex)** | |
| | **Male** | **Female** | **Male** | **Female** | **Male** | **Female** |
| 0-9 | 0.250 (0.036) | 0.253 (0.036) | 0.087 (0.019) | 0.090 (0.020) | 0.749 (0.113) | 0.608 (0.096) |
| 10-19 | 0.377 (0.051) | 0.457 (0.062) | 0.135 (0.030) | 0.162 (0.034) | 0.400 (0.066) | 0.375 (0.064) |
| 20-29 | 0.454 (0.060) | 0.681 (0.089) | 0.163 (0.035) | 0.242 (0.050) | 0.560 (0.084) | 0.757 (0.111) |
| 30-39 | 0.443 (0.060) | 0.748 (0.094) | 0.158 (0.034) | 0.264 (0.056) | 0.378 (0.056) | 0.583 (0.084) |
| 40-49 | 0.389 (0.053) | 0.732 (0.095) | 0.138 (0.029) | 0.259 (0.056) | 0.363 (0.055) | 0.541 (0.089) |
| 50-59 | 0.427 (0.061) | 0.792 (0.107) | 0.150 (0.032) | 0.279 (0.059) | 0.484 (0.081) | 0.669 (0.116) |
| 60-69 | 0.412 (0.060) | 0.651 (0.088) | 0.145 (0.031) | 0.230 (0.048) | 0.663 (0.130) | 0.538 (0.107) |
| 70-79 | 0.399 (0.061) | 0.524 (0.072) | 0.140 (0.031) | 0.186 (0.041) | 0.719 (0.187) | 0.298 (0.110) |
| 80+ | 0.425 (0.082) | 0.556 (0.090) | 0.152 (0.041) | 0.197 (0.047) | 0.301 (0.156) | 0.608 (0.243) |

*Probability of severe symptoms by age and sex*

We assumed that all individuals experiencing severe CHIKV symptoms are hospitalised. For each age and sex group, we estimated the probability that an individual with symptomatic infection develops severe symptoms resulting in hospitalisation (Table SS9). First, we assumed that all medically attended infections are detected, with all other infections going undetected, and estimated the proportion of symptomatic infections by age and sex that seek healthcare and are therefore detected. Reflecting data from the 2023 Paraguay outbreak[10,21], we multiplied the base-case estimated number of symptomatic infections by 12% to estimate the number of symptomatic infections that seek care and are detected. Among these detected cases, we assumed that 3.2% are admitted to hospital due to severe symptoms, corresponding to 58,250 hospitalised infections out of 1,820,656 total detected infections reported from January 2017 to April 2025 in  a comprehensive public database of CHIKV infections in Brazil[22]. These hospitalised cases were then distributed into age-sex groups to reproduce the distribution of severe symptoms by age and sex from Torales *et al.* based on the 2022-2023 outbreak in Paraguay[23], allowing derivation of age-sex specific numbers of severe disease (and hospitalisations). Then, for each age-sex group, we divided this number of hospitalised cases by the estimated number of symptomatic infections to derive age-sex-specific probabilities of severe symptoms given symptomatic infection. Uncertainty was captured by using the randomly sampled number of symptomatic infections by age and sex derived above, and randomly sampling age-sex distribution of severe symptoms by applying a Dirichlet distribution to the reported severe symptoms stratified by age and sex from Torales *et al.* Table SS9 summarizes the estimated probability of severe symptoms given symptomatic infection.

Table SS9. Estimated probability of severe symptoms at symptomatic infection.

| **Age group** | **Mean probability of severe symptoms given symptomatic infection (95% UI)** | |
|---|---|---|
| | **Male** | **Female** |
| 0-9 | 0.007 (0.005, 0.009) | 0.009 (0.007, 0.013) |
| 10-19 | 0.002 (0.001, 0.002) | 0.002 (0.001, 0.003) |
| 20-29 | 0.002 (0.002, 0.003) | 0.002 (0.002, 0.003) |
| 30-39 | 0.002 (0.002, 0.003) | 0.002 (0.001, 0.002) |
| 40-49 | 0.002 (0.001, 0.002) | 0.001 (0.001, 0.002) |
| 50-59 | 0.003 (0.002, 0.004) | 0.002 (0.002, 0.003) |
| 60-69 | 0.008 (0.006, 0.011) | 0.006 (0.005, 0.008) |
| 70-79 | 0.024 (0.018, 0.034) | 0.021 (0.016, 0.029) |
| 80+ | 0.055 (0.037, 0.086) | 0.035 (0.026, 0.050) |

*Probability of chikungunya death*

For each age and sex group, we estimated the probability of death, which was assumed to occur only among hospitalised individuals with severe symptoms (Table SS10). First, we estimated the number of deaths by age and sex using the base-case estimated age-sex specific number of symptomatic infections, probability of symptomatic infection being detected (0.12 as mentioned above) and the probability of death at detected symptomatic infections by age and sex from de Souza *et al.* based on the detected symptomatic cases over 2016-2022 in Brazil[24]. Secondly, for each age-sex group, we divided the estimated number of deaths by the base-case estimated number of hospitalised cases to derive the age-sex specific probability of death given severe symptoms. Uncertainty around the probability of death at detected symptomatic infections by age and sex was estimated by applying a Beta distribution to the estimated mean and standard error.

Table SS10. Estimated probability of death at severe symptoms.

| Age group | Probability of chikungunya death at severe symptoms (UIs) | |
|---|---|---|
| | Male | Female |
| 0-9 | 0.051 (0.028, 0.084) | 0.029 (0.015, 0.048) |
| 10-19 | 0.110 (0.054, 0.192) | 0.032 (0.011, 0.062) |
| 20-29 | 0.038 (0.014, 0.073) | 0.048 (0.027, 0.076) |
| 30-39 | 0.061 (0.032, 0.101) | 0.027 (0.014, 0.045) |
| 40-49 | 0.097 (0.049, 0.166) | 0.065 (0.037, 0.100) |
| 50-59 | 0.073 (0.041, 0.116) | 0.060 (0.035, 0.091) |
| 60-69 | 0.024 (0.012, 0.042) | 0.014 (0.007, 0.023) |
| 70-79 | 0.027 (0.016, 0.041) | 0.020 (0.013, 0.030) |
| 80+ | 0.016 (0.008, 0.027) | 0.024 (0.014, 0.037) |

*Probability of chronic chikungunya*

Evidence suggests that all individuals with symptomatic infection, and not only those with severe disease, are at risk of chronic chikungunya following recovery from acute infection [25]. In the absence of evidence that chronic chikungunya risk varies by age or sex, we applied the same mean probability derived from the systematic review by Kang *et al.*[9], 0.51 (95% CI 0.45, 0.58), to all individuals surviving acute detected infection. Uncertainty was estimated by applying a Beta distribution to the observed probability estimate (mean and standard error from the 95%CIs). In the sensitivity analysis, we applied the same probability of chronic symptoms additionally in individuals with undetected symptomatic infections.

#### *2f. Disability-adjusted life years estimation*

The disability-adjusted life year (DALY) is a synthetic indicator for measuring health effects generically, ranging for each person-year from a value of 0 (equal to no negative health effect) to 1 (equal to one year of healthy life lost). DALYs can therefore be used to measure disease burden, in the form of disability and associated health disutility, and years of life lost prematurely. We estimated DALYs by assigning disability weights for the durations of disease estimated for each health state and by calculating the age-specific number of years of life lost due to chikungunya.

The Global Burden of Disease (GBD) study has not produced a disability weight for chikungunya, so proxy measures are necessary for both acute and chronic disease. A key clinical characteristic of acute chikungunya is joint pain. In a Brazilian study by Silva *et al.* describing follow-up in 153 chikungunya patients, approximately 88.9% reported experiencing arthralgia during the acute phase of disease.[26] For detected acute cases with mild/moderate symptoms, we therefore considered the GBD disability weight for mild rheumatoid arthritis of 0.117 (95%CI 0.080, 0.163) as a proxy in 88.9% of patients, and the GBD disability weight for moderate febrile illness of 0.051 (95% CI 0.032, 0.074) in the remaining 11.1%, for a final disability weight of 0.110 (0.074, 0.145).[27] For undetected acute mild/moderate symptomatic cases, in the base-case, a disability weight of 0.006 (95%CI 0.002, 0.012) for mild febrile illness from the global burden of disease (GBD) 2017 study[27] was used. In a sensitivity analysis, the same disability weight was used for detected and undetected acute mild/moderate symptomatic cases. For acute severe symptoms, a disability weight of 0.581 (95% CI 0.403, 0.739) for severe arthralgia from GBD 2017 was used over the prehospitalization period[27], and of 0.810 (Range 0.600, 0.920) was used over the hospitalization period based on the disability weight from a Colombian study by Mora-Salamanca *et al*.[28]

For chronic symptoms, de Roo *et al*.[29] used the disability weight of 0.349 (95% CI 0.117, 0.581) based on the study by Puntasecca *et al*.[30] reporting the disability weight for severe rheumatoid arthritis. Ribeiro dos Santos *et al.*[10] used the disability weight of 0.233 based on the GBD 2013 study reporting the disability weight for arthralgia[31]. None of these studies refer to data specifically from patients with chronic chikungunya.

Therefore, we took a different approach and estimated the disability weight associated with chronic chikungunya based on 6 studies[32-37] reporting Health Assessment Questionnaire - Disability Index (HAQ-DI) for chronic chikungunya. We used these scores to generate the disability weight with the following steps (1) we mapped the HAQ-DI to EQ5D utility based on an algorithm developed based on 3,557 rheumatoid arthritis patients by Kim *et al*.[38]; (2) we used the population utility norm in the corresponding countries[39-41] to subtract the mapped utility to derive the disability weight from each study; and (3) we performed meta-analysis of these disability weights using an inverse-variance approach. Because two studies[33,34] estimated HAQ-DI in patients from Martinique Island and Reunion Island for which population norms are not available, we used population norms from mainland France. Recognising the fact that the populations and living conditions in these overseas territories may be different from mainland France, in the base-case analyses we excluded these two studies from the meta-analysis resulting in a disability weight of 0.230 (95% CI 0.216, 0.243). In sensitivity analysis the two studies were included in the meta-analysis resulting in a disability weight of 0.332 (95% CI 0.307, 0.356).

Table SS11. Summary of reported Health Assessment Questionnaire – Disability Index (HAQ-DI) for chronic chikungunya and disability weight mapping.

| **Study** | **Country** | **Age, years (mean/ median)** | **HAQ-DI, Mean (estimated SE)** | **Mapped EQ-5D utility (SE)** | **Population utility norm (SE)** | **Disability weight (SE)** |
|---|---|---|---|---|---|---|
| Pandya 2008[37] | India | 49.0 | 1.60 (0.03) | 0.42 (0.01) | 0.82 (0.01)[40] | 0.40 (0.02) |
| Ganu 2011[35] | India | NR | 2.18 (0.16) | 0.26 (0.04) | 0.86 (0.01) [40] | 0.60 (0.04) |
| Ravindran 2017[36] | India | 54.1 | 1.96 (0.01) | 0.32 (0.00) | 0.64 (0.05) [40] | 0.32 (0.05) |
| Blettery 2016[33] | France | 64.1 | 1.40 (0.08) | 0.48 (0.02) | 0.90 (0.00)[39] | 0.42 (0.02) |
| Bouquillard 2018[34] | France | 54.0 | 0.44 (0.03) | 0.75 (0.01) | 0.90 (0.00) [39] | 0.15 (0.01) |
| Kennedy Amaral 2019[32] | Brazil | 56.8 | 1.00 (0.07) | 0.59 (0.02) | 0.76 (0.01)[41] | 0.17 (0.02) |

NR = not reported

Estimates reported in the literature were used to define the duration of time spent in each health state. The duration of acute mild/moderate symptoms was 6 days (range 2, 21), as reported in household investigations of the 2014-2015 chikungunya outbreak in Puerto Rico by Sharp *et al*.[42] The duration of acute severe symptoms included the duration for pre-hospitalization and hospitalization. The duration of pre-hospitalization was 6 days (range 2, 21) assuming the same as the duration of acute mild/moderate symptoms; and the duration of hospitalization was 5 days (range 2, 12), as reported in the national hospitalization database analysis in the chikungunya epidemic on La Reuion Island over 2005-2006 by Soumahoro *et al*[43]. The duration of chronic symptoms was 0.602 years (95%CIs 0.510, 0.735) in patients younger than 40 years old, and 1.000 years (95%CIs 0.819, 1.300) in patients older than 40 years old, estimated based on the monthly rate of arthralgia resolution reported in the systematic review of 16 cohort studies by O'Driscoll *et al*[44].

Last, non-survivors were assigned a full DALY for each year of their remaining age-specific life expectancy. Table SS12 summarizes the disability weights and durations associated with each health state included in the model. Uncertainty was estimated by applying a Beta distribution to the observed estimates for disability weight; and normal distribution to the observed estimates for duration of health states.

Table SS12. Summary of the model input for disability weight and duration of model health states.

| Parameter | Mean (95% CI) | Source |
|---|---|---|
| **Disability weight** | | |
| Acute mild/moderate symptoms | | |
| Detected | 0.110 (0.074, 0.145) | Based on Silva *et al*[26], GBD 2017 study[27] |
| Undetected (Base-case) | 0.006 (0.002. 0.012) | GBD 2017 study[27] |
| Undetected (Sensitivity analysis) | 0.110 (0.074, 0.145) | Same as detected case |
| Acute severe symptoms | | |
| Pre-hospitalization | 0.583 (0.403, 0.739) | Silva *et al*[26] |
| Hospitalization | 0.810 (0.675, 0.945) | Mora-Salamanca *et al*[28] |
| Chronic symptoms | | |
| Base-case | 0.229 (0.216, 0.242) | Estimated from multiple sources (Supplementary section 1e) |
| Sensitivity analysis | 0.331 (0.306, 0.355) | |
| **Duration (years)** | | |
| Acute mild/moderate symptoms | 0.024 (0.012, 0.037) | Sharp *et al*[42] |
| Acute severe symptoms | | |
| Pre-hospitalization | 0.024 (0.012, 0.037) | Assumed the same as acute mild/moderate symptom |
| Hospitalization | 0.014 (0.005, 0.033) | Soumahoro *et al*[43] |
| Chronic symptoms | | |
| <40 years old | 0.602 (0.510, 0.735) | Estimated from O'Driscoll *et al*[44] |
| >= 40 years old | 1.000 (0.819, 1.300) | |

#### *2g. Healthcare seeking by symptoms*

As described in supplementary method 2d, we used 12% as the probability of symptomatic infection seeking care[10,21], and 3.8% as the probability of infections seeking care requiring hospitalisation due to severe symptoms (12% × 3.8% = 0.38% probability of symptomatic infections requiring hospitalisation).[22] We therefore assumed that all remaining detected symptomatic infections (96.2%) have mild/moderate symptoms detected in outpatient care. Thus we used 11.6% as the probability of acute mild/moderate symptoms seeking any care [from 12% × 96.2% / (1 - 12% × 3.8%)]. For chronic symptoms, the probability of ever seeking outpatient care was 19.4% (95%CIs: 0.151, 0.237) based on a cohort study of 283 patients with chronic chikungunya in Colombia[45]. This is aligned with a small qualitative study in Curaçao showing that most participants chose to self-manage their symptoms rather than seeking care in a formal healthcare setting[46].

Table SS13. Summary of estimates for healthcare seeking by symptoms.

| Symptoms | Probability of ever seeking care (95%CIs) |
|---|---|
| **Acute mild/moderate symptoms** | 0.116 |

| | |
|---|---|
| **Acute severe symptoms** | 1 |
| **Chronic symptoms** | 0.194 (0.151, 0.237) |

***2h. Economic parameters estimation***

Economic impact was estimated by deriving country-specific economic parameters for all 32 included countries except Venezuela, which was excluded due to a lack of reliable estimates of economic parameters following its economic collapse in recent years.

*Treatment costs*

We considered the following healthcare costs associated with chikungunya disease: (1) outpatient care costs for acute mild/moderate symptoms; (2) inpatient care costs for acute severe symptoms; and (3) outpatient care costs for chronic symptoms.

Costs of outpatient care for acute mild/moderate symptoms were based on a study in Colombia by Alvis-Zakzuk *et al.*, which reported a median cost of $30.9 in 2014 USD for outpatient care per chikungunya patient, accounting for consultation, drugs and laboratory & imaging and procedures[47]. Outpatient care costs for acute mild/moderate disease in other countries were derived using the scaling factor in 2016 PPP I$ estimated by Moses *et al.*[48]. Firstly, the costs in 2014 USD in Colombia were converted to 2016 PPP I$ in Colombia using the approach recommended by Turner *et al.*[49]. Using this approach, we completed the conversion using the following steps (1) convert the costs in 2014 USD to its value in the local currency (COP) using the 2014 exchange rate; (2) inflate the costs in 2014 COP to its value in 2016 using World Bank GDP deflator values[50]; (3) convert the costs in 2016 COP to its value in PPP I$ using the 2016 exchange rate. Then, to derive cost estimates for other countries, the costs in 2016 PPP I$ in Colombia were scaled to the costs in 2016 PPP I$ in the 30 other countries using the scaling factors from a global study by Moses *et al.*[48]. In that study, outpatient costs were estimated as expenditure per capita on outpatient care divided by outpatient utilisation per capita[48]. Finally, the costs in 2016 PPP I$ in each country were converted to its value in 2023 PPP I$ in the country using the approach by Turner *et al.* mentioned above[49].

Costs of inpatient care (hospitalisation) for acute severe symptoms were also based on the study in Colombia by Alvis-Zakzuk *et al.*, which reported a median cost of $318.9 in 2014 USD for inpatient care for chikungunya patients, accounting for consultation, drugs, laboratory & imaging, procedures and hospital admissions[47]. We converted the costs to 2023 PPP I$ in Colombia and derived the costs in the other countries using the same approach described above (except using the scaling factor for inpatient costs instead of the factor for outpatient costs in Moses *et al.*[48]).

Costs of outpatient care for chronic symptoms were based on a study in Brazil by Goncalves *et al.*,[51] which reported a mean cost of R$180.24 and R$314.29 in 2019 BRL for outpatient care for chronic symptoms of different severities. These costs comprise three consultations per patient (regardless of severity) and corresponding medications (hydroxychloroquine 325mg/day for mild patients, and methotrexate 15mg/week plus folic acid for moderate and severe patients) over a 6-month time horizon. The mean costs of R$268.85 were used based on the assumption that 66.1% chronic symptoms are severe, based on another Brazilian study by Silva *et al*[26]. An alternative cost estimate for chronic chikungunya from Cardona-Ospina *et al.* used Colombian rheumatoid arthritis guidelines to define treatment pathways, implicitly only considering severe chronic chikungunya. In their most conservative scenario, they estimated that drugs (62%) – prednisolone 5mg/day, methotrexate 15 mg/week (plus folic acid) and sulfasalazine 1.5g/day) – consultations (11%), and diagnostic tests (27%) would result in I$ 3,582.07 (2021) over 2 years[52]. We used the Brazilian estimate in our model because: (1) it is more recent, (2) it is based on chikungunya-specific guidelines (including for instance the incorporation of hydroxychloraquine) rather than rheumatoid arthritis guidelines; (3) it explicitly accounts for variable symptom severity; and (4) its time horizon is more conservative, projecting 6 months rather than for 2 years of treatment. As for acute care costs, we converted chronic care costs to 2023 PPP I$ in Brazil and derived the costs in the other countries using the approach described above.

Table SS14. Summary of healthcare costs for each health state.

| Country | Unit healthcare costs (I$ 2023) | | |
|---|---|---|---|
| | Outpatient care Acute symptoms Mild/Moderate[a] | Inpatient care Acute symptoms Severe[a] | Outpatient care Chronic symptoms[b] |
| **Belize** | 98 | 303 | 211 |
| **Bolivia** | 43 | 195 | 93 |
| **Brazil** | 149 | 497 | 320 |
| **Cambodia** | 29 | 327 | 63 |
| **Chad** | 14 | 64 | 29 |
| **Colombia** | 73 | 750 | 156 |
| **Costa Rica** | 213 | 710 | 459 |
| **Democratic Republic of Congo** | 6 | 17 | 12 |
| **Djibouti** | 16 | 73 | 35 |
| **Dominican Republic** | 147 | 316 | 316 |
| **Ecuador** | 93 | 410 | 201 |
| **El Salvador** | 143 | 804 | 307 |
| **Ethiopia** | 9 | 75 | 20 |
| **Grenada** | 103 | 272 | 221 |
| **Guatemala** | 52 | 260 | 113 |
| **Guyana** | 30 | 193 | 64 |
| **Haiti** | 21 | 59 | 44 |
| **Honduras** | 41 | 193 | 89 |
| **India** | 23 | 104 | 50 |
| **Indonesia** | 58 | 439 | 125 |
| **Malaysia** | 83 | 1258 | 178 |
| **Mexico** | 148 | 512 | 319 |
| **Nicaragua** | 32 | 229 | 69 |
| **Panama** | 79 | 430 | 170 |
| **Paraguay** | 90 | 333 | 193 |
| **Peru** | 65 | 308 | 140 |
| **Philippines** | 37 | 519 | 79 |
| **Republic of Congo** | 36 | 110 | 77 |
| **St. Lucia** | 102 | 356 | 220 |
| **Sudan** | 19 | 111 | 42 |
| **Thailand** | 137 | 598 | 295 |

[a] healthcare costs for acute symptoms were assumed to incur over 1 month; [b] healthcare costs for chronic symptoms were assumed to incur over 6 months

*Risk of catastrophic / impoverishing healthcare expenditures*

The occurrence of large out-of-pocket (OOP) healthcare expenses can lead to financial hardship, depending on the capacity of the individual or household to pay. Catastrophic healthcare expenditure (CHE) and impoverishing healthcare expenditure (IHE) are two possible metrics to measure this. CHE has a flexible definition and consequently a variety of methods with which to calculate it[53]. Here, we define the risk of CHE as health expenditure constituting ≥ 10% of individual annual income, and the risk of IHE as health expenditure that would impact the individual's poverty status such that they would be pushed below the poverty line. Country-specific poverty lines were from the World Bank were used where available, while the international poverty line of 2.15 I$ per day was used for countries not included in World Bank estimates[54] (Table SS15). Based on these definitions, the estimation of CHE and IHE due to chikungunya can be derived from estimates of the proportion of monthly healthcare expenditures paid OOP and the country's income level.

Table SS15. Summary of poverty line in each country

| Country | Poverty lines of income per day (I$ 2023) |
|---|---|
| **Belize** | 9.59 |
| **Bolivia** | 4.67 |
| **Brazil** | 9.32 |
| **Cambodia*** | 2.15 |
| **Chad** | 3.04 |
| **Colombia** | 10.01 |
| **Costa Rica** | 9.03 |
| **Democratic Republic of Congo** | 3.08 |
| **Djibouti** | 5.03 |
| **Dominican Republic** | 9.88 |
| **Ecuador** | 9.89 |
| **El Salvador** | 9.58 |
| **Ethiopia** | 2.85 |
| **Grenada*** | 2.15 |
| **Guatemala** | 11.88 |
| **Guyana** | 44.33 |
| **Haiti** | 5.17 |
| **Honduras** | 4.89 |
| **India** | 4.74 |
| **Indonesia** | 9.24 |
| **Malaysia** | 9.20 |
| **Mexico** | 9.14 |
| **Nicaragua** | 5.25 |
| **Panama** | 27.58 |
| **Paraguay** | 8.66 |
| **Peru** | 9.63 |
| **Philippines** | 4.59 |
| **Republic of Congo** | 6.75 |

| Country | Poverty lines of income per day (I$ 2023) |
|---|---|
| **St. Lucia** | 11.75 |
| **Sudan** | 3.72 |
| **Thailand** | 9.30 |

* Country specific poverty line was not estimated in the report by the world bank[54] thus the international poverty line of 2.15 I$ was used

### OOP healthcare expenditure

We estimated the proportion of healthcare expenditure paid OOP in each country for each of the three main forms of healthcare utilisation included in the model (outpatient consultation for acute mild/moderate disease, hospitalisation for acute severe disease, and outpatient consultation for chronic disease).

For chikungunya hospitalisation, the OOP proportion was estimated based on costing data from Alvis-Zakzuk *et al.*, who reported both OOP costs ($5 USD) and the total inpatient care costs ($318.9 USD) for acute chikungunya hospitalisation in Colombia in 2014[47]. We used this OOP proportion (1.6%) as the proportion for Colombia and estimated the OOP proportion for the other countries with a scaling factor. Firstly, we sourced country-level World Bank data on current OOP health expenditure as a percentage of current total health expenditure[55]. Secondly, we adjusted these proportions for all countries based on the scaling factor generated by dividing the proportion of chikungunya hospitalisation costs paid OOP in the Colombia study (1.6%) by the World Bank proportion of all healthcare costs paid OOP in Colombia (13.67%) in 2021 (most recent data).

For outpatient care costs, the OOP proportion was assumed to be the same for acute mild/moderate chikungunya and for chronic chikungunya. Given that OOP costs for outpatient chikungunya care are not well known, we used the OOP proportion for costs of acute chikungunya including only drugs and consultations ($5 USD out of $71.6 USD) from Alvis-Zakzuk *et al.* in Colombia in 2014[47]. We used this OOP proportion (7.0%) as the proportion for Colombia and estimated the OOP proportion for the other countries using the same approach mentioned above for chikungunya hospitalisation costs.

The estimated total OOP costs for inpatient and outpatient care were broken into monthly costs, assuming that treatment costs for acute disease are incurred over 1 month and the treatment costs for chronic disease are incurred over 6 months.

Table SS16. Summary of out-of-pocket (OOP) healthcare expenditure for each health state.

| Country | OOP% | | OOP/Month (I$ 2023) | | |
|---|---|---|---|---|---|
| | | | Outpatient care | | Inpatient |
| | Outpatient care | Inpatient care | Acute mild/moderate | Chronic | Acute severe |
| **Belize** | 0.116 | 0.026 | 11.4 | 4.1 | 7.9 |
| **Bolivia** | 0.116 | 0.026 | 5.0 | 1.8 | 5.1 |
| **Brazil** | 0.116 | 0.026 | 17.2 | 6.2 | 12.9 |
| **Cambodia** | 0.281 | 0.063 | 8.2 | 2.9 | 20.6 |
| **Chad** | 0.300 | 0.067 | 4.1 | 1.5 | 4.3 |
| **Colombia** | 0.070 | 0.016 | 5.1 | 1.8 | 11.8 |
| **Costa Rica** | 0.106 | 0.024 | 22.6 | 8.1 | 16.9 |
| **Democratic Republic of Congo** | 0.200 | 0.045 | 1.1 | 0.4 | 0.8 |
| **Djibouti** | 0.104 | 0.023 | 1.7 | 0.6 | 1.7 |

| Country | OOP% | | OOP/Month (I$ 2023) | | |
|---|---|---|---|---|---|
| | | | Outpatient care | | Inpatient |
| | Outpatient care | Inpatient care | Acute mild/moderate | Chronic | Acute severe |
| **Dominican Republic** | 0.121 | 0.027 | 17.7 | 6.3 | 8.6 |
| **Ecuador** | 0.156 | 0.035 | 14.6 | 5.2 | 14.4 |
| **El Salvador** | 0.137 | 0.031 | 19.5 | 7.0 | 24.7 |
| **Ethiopia** | 0.189 | 0.042 | 1.8 | 0.6 | 3.2 |
| **Grenada** | 0.274 | 0.062 | 28.1 | 10.1 | 16.7 |
| **Guatemala** | 0.312 | 0.070 | 16.3 | 5.8 | 18.2 |
| **Guyana** | 0.147 | 0.033 | 4.4 | 1.6 | 6.4 |
| **Haiti** | 0.222 | 0.050 | 4.6 | 1.6 | 2.9 |
| **Honduras** | 0.264 | 0.059 | 10.9 | 3.9 | 11.5 |
| **India** | 0.255 | 0.057 | 5.9 | 2.1 | 6.0 |
| **Indonesia** | 0.140 | 0.032 | 8.2 | 2.9 | 13.8 |
| **Malaysia** | 0.164 | 0.037 | 13.6 | 4.9 | 46.3 |
| **Mexico** | 0.211 | 0.047 | 31.3 | 11.2 | 24.3 |
| **Nicaragua** | 0.157 | 0.035 | 5.0 | 1.8 | 8.1 |
| **Panama** | 0.191 | 0.043 | 15.1 | 5.4 | 18.4 |
| **Paraguay** | 0.184 | 0.041 | 16.4 | 5.9 | 13.7 |
| **Peru** | 0.139 | 0.031 | 9.1 | 3.3 | 9.6 |
| **Philippines** | 0.228 | 0.051 | 8.4 | 3.0 | 26.6 |
| **Republic of Congo** | 0.123 | 0.028 | 4.4 | 1.6 | 3.0 |
| **St. Lucia** | 0.190 | 0.043 | 19.5 | 7.0 | 15.2 |
| **Sudan** | 0.290 | 0.065 | 5.6 | 2.0 | 7.3 |
| **Thailand** | 0.046 | 0.010 | 6.3 | 2.3 | 6.2 |

### Risk of CHE or IHE in different symptoms

The number of individuals at risk of CHE or IHE due to chikungunya healthcare expenditure depends on the sum total of OOP healthcare expenditure per month for acute and chronic disease. In order to calculate CHE, we identified a threshold of country-specific OOP expenditure x 10 as ‘income at risk’, where the percentage of the population with income below this threshold is considered at risk of CHE. A distinct threshold was identified for each considered form of chikungunya healthcare expenditure (acute outpatient, chronic outpatient, acute inpatient). CHE for chronic chikungunya was only considered among those not seeking care for acute disease, to avoid double counting CHE (as OOP expenditure for chronic outpatient care was estimated in all countries to be less than OOP expenditure for either acute outpatient care or acute inpatient care, so patients at risk of CHE/IHE due to OOP expenditure for management of acute disease will also be at risk of CHE/IHE due to OOP expenditure for management of chronic disease; Table SS16).

We fitted a generalized beta distribution of the second kind to decile-level household income data from the World Income Inequality Database (WIID)[56] to estimate the proportion of the population at risk of CHE (Fig SS10). To estimate the population at risk of IHE, this exercise was repeated after altering income thresholds by adding the international poverty line [54] (Table SS15) to each of the acute and chronic OOP costs. The number of individuals falling below this threshold would fall below the poverty line if forced to pay estimated OOP expenditure for the treatment of symptomatic CHIKV infection (Table SS16).

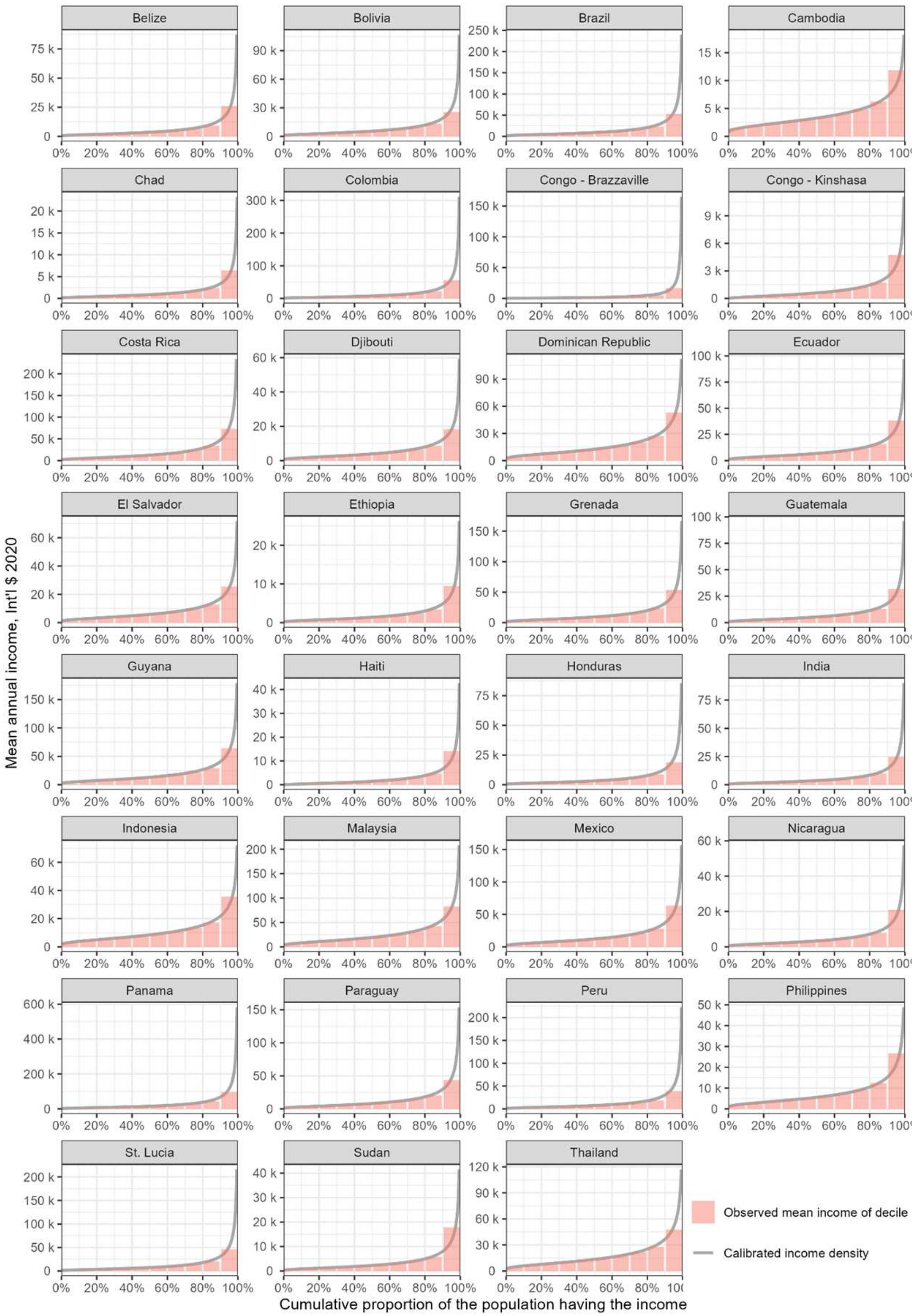

Fig SS10. Income distribution by country. Restricting data to cumulative proportion up until 99.5%.

Table SS17. Summary of proportion of patients at risk of catastrophe healthcare expenditure (CHE) or impoverishing healthcare expenditure (IHE) while receiving care for different symptoms caused by CHIKV infection.

| Country | % at risk CHE | | | % at risk IHE | | |
|---|---|---|---|---|---|---|
| | Mild/Moderate | Severe | Chronic | Mild/Moderate | Severe | Chronic |
| **Belize** | 11.2% | 3.9% | 0.2% | 1.7% | 1.2% | 0.6% |
| **Bolivia** | 0.0% | 0.0% | 0.0% | 0.6% | 0.6% | 0.2% |
| **Brazil** | 2.0% | 0.5% | 0.0% | 1.7% | 1.2% | 0.6% |
| **Cambodia** | 0.9% | 30.2% | 0.0% | 0.2% | 0.9% | 0.1% |
| **Chad** | 24.4% | 26.7% | 1.6% | 1.8% | 1.9% | 0.6% |
| **Colombia** | 0.0% | 1.4% | 0.0% | 0.6% | 1.4% | 0.2% |
| **Costa Rica** | 1.9% | 0.5% | 0.0% | 1.1% | 0.8% | 0.4% |
| **Democratic Republic of Congo** | 8.1% | 4.3% | 1.3% | 0.4% | 0.3% | 0.1% |
| **Djibouti** | 0.0% | 0.0% | 0.0% | 0.4% | 0.4% | 0.1% |
| **Dominican Republic** | 0.1% | 0.0% | 0.0% | 0.6% | 0.3% | 0.2% |
| **Ecuador** | 1.8% | 1.7% | 0.0% | 2.0% | 2.0% | 0.7% |
| **El Salvador** | 8.0% | 15.6% | 0.0% | 3.2% | 4.0% | 1.1% |
| **Ethiopia** | 0.3% | 3.5% | 0.0% | 1.0% | 1.7% | 0.4% |
| **Grenada** | 8.0% | 1.2% | 0.1% | 0.1% | 0.0% | 0.0% |
| **Guatemala** | 8.2% | 11.1% | 0.1% | 2.2% | 2.5% | 0.8% |
| **Guyana** | 0.0% | 0.0% | 0.0% | 0.2% | 0.2% | 0.1% |
| **Haiti** | 20.4% | 11.9% | 5.3% | 1.0% | 0.6% | 0.4% |
| **Honduras** | 13.2% | 14.7% | 0.4% | 2.9% | 3.0% | 1.0% |
| **India** | 1.1% | 1.2% | 0.0% | 1.5% | 1.5% | 0.5% |
| **Indonesia** | 0.0% | 0.2% | 0.0% | 0.7% | 1.1% | 0.2% |
| **Malaysia** | 0.0% | 2.6% | 0.0% | 0.1% | 0.3% | 0.0% |
| **Mexico** | 3.8% | 1.3% | 0.0% | 1.3% | 1.0% | 0.4% |
| **Nicaragua** | 0.5% | 3.8% | 0.0% | 1.4% | 2.2% | 0.5% |
| **Panama** | 0.2% | 0.4% | 0.0% | 0.8% | 0.9% | 0.3% |
| **Paraguay** | 2.3% | 1.1% | 0.0% | 1.7% | 1.4% | 0.6% |
| **Peru** | 0.3% | 0.4% | 0.0% | 1.1% | 1.2% | 0.4% |
| **Philippines** | 0.2% | 19.7% | 0.0% | 0.7% | 2.5% | 0.2% |
| **Republic of Congo** | 12.3% | 5.0% | 0.4% | 0.6% | 0.4% | 0.2% |
| **St. Lucia** | 7.4% | 3.4% | 0.1% | 2.2% | 1.7% | 0.8% |
| **Sudan** | 4.9% | 10.1% | 0.0% | 2.2% | 2.8% | 0.8% |
| **Thailand** | 0.0% | 0.0% | 0.0% | 0.1% | 0.1% | 0.0% |

*Productivity losses*

Productivity losses due to chikungunya were estimated by multiplying days or years of work missed due to disease by the corresponding daily or annual gross national income (GNI). Country-specific per capita GNI (in I$ 2023) used for this calculation was obtained from the World Bank (Table SS18)[57]. We used GNI as a proxy for wages, which we assumed to remain constant for the time horizon of the analysis.

Table SS18. Country-specific per-capita GNI estimates.

| Country | Per-capita GNI estimates (I$ 2023) |
|---|---|
| **Belize** | 13,710 |
| **Bolivia** | 10,490 |
| **Brazil** | 19,990 |
| **Cambodia** | 5,460 |
| **Chad** | 1,940 |
| **Colombia** | 21,420 |
| **Costa Rica** | 25,800 |
| **Democratic Republic of Congo** | 1,620 |
| **Djibouti** | 7,040 |
| **Dominican Republic** | 24,460 |
| **Ecuador** | 15,510 |
| **El Salvador** | 11,750 |
| **Ethiopia** | 3,100 |
| **Grenada** | 16,550 |
| **Guatemala** | 13,820 |
| **Guyana** | 50,060 |
| **Haiti** | 3,260 |
| **Honduras** | 6,680 |
| **India** | 10,030 |
| **Indonesia** | 15,210 |
| **Malaysia** | 36,120 |
| **Mexico** | 24,970 |
| **Nicaragua** | 7,640 |
| **Panama** | 38,110 |
| **Paraguay** | 16,910 |
| **Peru** | 15,860 |
| **Philippines** | 11,940 |
| **Republic of Congo** | 6,550 |
| **St. Lucia** | 22,980 |
| **Sudan** | 3,110 |
| **Thailand** | 22,880 |

Productivity losses include (1) number of productive days lost from work absences due to symptomatic disease; and (2) years lost from premature mortality.

Patients with acute symptoms were assumed to be fully absent from work for the full duration of their symptoms (see Table SS12). Patients with chronic symptoms were assumed to be partially absent for the full duration of their symptoms. The proportion of individuals with chronic symptoms who are absent from work was estimated to be 30.4%, based on a UK study evaluating the impacts of rheumatoid arthritis of different severity levels on employment from Barrett *et al.* (mild: 0.14; moderate: 0.26; severe: 0.33)[58] and the severity distribution of chronic chikungunya from a Brazilian study from Silva *et al.* (mild: 0.016; moderate: 0.323; severe: 0.661)[26]. The latest available proportion of the working population engaged in the workforce by age for each country was derived from the International Labour Organisation (ILO) [59].

For productivity losses due to premature mortality caused by chikungunya, we estimated the years of productive life lost based on age at death, the average age-specific remaining life expectancy, and each age group's labour force participation. Projections of future age-specific life expectancies from 2025 to 2050 were taken from UN World Population Prospects (medium variant), the same demographic data used to define country- and age-specific population sizes over the model's time horizon.)[14] The proportion of working population engaged in the workforce in the country was taken from the latest year with the relevant data. Due to an absence of labour force projections, we assumed that labour force participation by age group and country is stable over the model's time horizon.

Table SS19. Proportion of people working in the age weighted by population size in the country in 2025

| Country | 0-9 | 10-19 | 20-29 | 30-39 | 40-49 | 50-59 | 60-69 | 70-79 | 80+ |
|---|---|---|---|---|---|---|---|---|---|
| **Belize** | 0.0% | 34.9% | 81.5% | 85.5% | 85.4% | 74.7% | 44.2% | 27.1% | 0.0% |
| **Bolivia** | 0.0% | 24.8% | 78.2% | 90.4% | 92.3% | 90.0% | 70.8% | 58.1% | 0.0% |
| **Brazil** | 0.0% | 17.3% | 76.9% | 82.1% | 79.7% | 67.0% | 31.0% | 14.9% | 0.0% |
| **Cambodia** | 0.0% | 50.7% | 86.4% | 92.9% | 92.7% | 83.8% | 58.1% | 40.4% | 0.0% |
| **Chad** | 0.0% | 14.1% | 57.4% | 74.2% | 77.4% | 77.0% | 64.9% | 55.4% | 0.0% |
| **Colombia** | 0.0% | 12.0% | 74.3% | 84.2% | 81.8% | 72.1% | 41.7% | 24.5% | 0.0% |
| **Costa Rica** | 0.0% | 7.4% | 69.8% | 81.2% | 79.9% | 67.2% | 32.2% | 15.7% | 0.0% |
| **Democratic Republic of Congo** | 0.0% | 16.4% | 58.6% | 75.6% | 80.1% | 77.2% | 68.4% | 59.9% | 0.0% |
| **Djibouti** | 0.0% | 3.7% | 31.3% | 43.6% | 48.4% | 37.4% | 21.5% | 12.0% | 0.0% |
| **Dominican Republic** | 0.0% | 12.1% | 75.9% | 85.1% | 82.9% | 71.9% | 44.9% | 26.8% | 0.0% |
| **Ecuador** | 0.0% | 13.7% | 69.1% | 83.0% | 83.0% | 76.9% | 53.7% | 37.3% | 0.0% |
| **El Salvador** | 0.0% | 19.0% | 70.9% | 80.1% | 76.3% | 69.6% | 42.4% | 29.3% | 0.0% |
| **Ethiopia** | 0.0% | 22.6% | 72.0% | 81.7% | 80.7% | 72.8% | 53.6% | 41.6% | 0.0% |
| **Grenada** | 0.0% | 7.8% | 80.1% | 90.0% | 87.8% | 81.2% | 35.0% | 20.6% | 0.0% |
| **Guatemala** | 1.1% | 38.5% | 74.3% | 81.4% | 80.3% | 73.3% | 53.8% | 42.0% | 0.0% |
| **Guyana** | 0.0% | 14.2% | 62.5% | 64.6% | 62.8% | 55.1% | 27.3% | 16.1% | 0.0% |
| **Haiti** | 0.0% | 10.3% | 63.6% | 85.2% | 87.5% | 82.7% | 66.3% | 58.1% | 0.0% |
| **Honduras** | 0.0% | 18.7% | 66.8% | 71.2% | 70.2% | 64.0% | 41.1% | 27.3% | 0.0% |
| **India** | 0.0% | 8.1% | 57.9% | 73.0% | 75.8% | 69.6% | 37.3% | 23.3% | 0.0% |
| **Indonesia** | 0.0% | 12.5% | 70.7% | 77.5% | 80.5% | 78.0% | 58.0% | 46.8% | 0.0% |
| **Malaysia** | 0.0% | 12.2% | 75.6% | 86.3% | 82.5% | 67.3% | 19.5% | 0.0% | 0.0% |
| **Mexico** | 0.0% | 15.0% | 68.4% | 78.0% | 76.7% | 69.5% | 39.1% | 25.7% | 0.0% |
| **Nicaragua** | 0.0% | 25.4% | 70.3% | 79.9% | 77.5% | 73.8% | 46.1% | 32.1% | 0.0% |
| **Panama** | 0.0% | 10.8% | 71.2% | 84.0% | 84.8% | 77.1% | 43.5% | 26.2% | 0.0% |
| **Paraguay** | 0.0% | 19.1% | 79.2% | 85.4% | 85.7% | 77.2% | 52.6% | 35.6% | 0.0% |
| **Peru** | 0.0% | 23.1% | 76.9% | 85.0% | 86.5% | 82.6% | 60.8% | 43.6% | 0.0% |
| **Philippines** | 0.0% | 8.9% | 62.0% | 78.0% | 80.7% | 75.7% | 49.5% | 35.0% | 0.0% |
| **Republic of Congo** | 0.0% | 8.3% | 55.4% | 78.8% | 77.8% | 68.2% | 27.8% | 19.7% | 0.0% |
| **St. Lucia** | 0.0% | 16.6% | 89.3% | 93.6% | 89.8% | 82.4% | 47.2% | 21.4% | 0.0% |
| **Sudan** | 0.0% | 9.8% | 38.4% | 44.9% | 47.3% | 46.9% | 33.5% | 30.6% | 0.0% |
| **Thailand** | 0.0% | 6.8% | 75.9% | 90.1% | 89.0% | 82.6% | 44.7% | 27.6% | 0.0% |

Table SS20. Estimated years of productive life lost due to premature death in each age group, weighted by population size in each country in 2025.

| Country | 0-9 | 10-19 | 20-29 | 30-39 | 40-49 | 50-59 | 60-69 | 70-79 | 80+ |
|---|---|---|---|---|---|---|---|---|---|
| **Belize** | 41.5 | 40.5 | 34.6 | 26.5 | 18.3 | 10.5 | 4.8 | 1.7 | 0.0 |
| **Bolivia** | 45.3 | 45.4 | 40.4 | 32.6 | 24.3 | 16.2 | 9.3 | 3.6 | 0.0 |
| **Brazil** | 36.3 | 36.0 | 31.3 | 23.4 | 15.5 | 8.1 | 2.9 | 0.9 | 0.0 |
| **Cambodia** | 47.2 | 45.4 | 39.2 | 30.5 | 22.1 | 13.4 | 6.9 | 2.6 | 0.0 |
| **Chad** | 30.3 | 32.5 | 30.4 | 25.2 | 19.0 | 12.8 | 7.5 | 3.5 | 0.0 |
| **Colombia** | 38.5 | 38.3 | 34.3 | 26.6 | 18.4 | 10.5 | 4.5 | 1.5 | 0.0 |
| **Costa Rica** | 35.4 | 35.3 | 31.7 | 24.1 | 16.0 | 8.3 | 3.0 | 1.0 | 0.0 |

| | | | | | | | | | |
|---|---|---|---|---|---|---|---|---|---|
| **Democratic Republic of Congo** | 35.3 | 36.4 | 33.5 | 27.9 | 21.1 | 14.5 | 8.8 | 3.8 | 0.0 |
| **Djibouti** | 18.5 | 18.6 | 17.2 | 13.5 | 9.1 | 4.8 | 2.1 | 0.7 | 0.0 |
| **Dominican Republic** | 38.6 | 38.5 | 34.4 | 26.6 | 18.3 | 10.8 | 4.9 | 1.7 | 0.0 |
| **Ecuador** | 40.8 | 40.7 | 36.8 | 29.3 | 21.3 | 13.4 | 6.5 | 2.3 | 0.0 |
| **El Salvador** | 37.1 | 36.6 | 32.5 | 25.4 | 17.3 | 10.4 | 5.0 | 1.8 | 0.0 |
| **Ethiopia** | 38.3 | 38.4 | 34.1 | 27.0 | 19.4 | 12.2 | 6.7 | 2.6 | 0.0 |
| **Grenada** | 39.2 | 39.1 | 35.0 | 26.6 | 18.3 | 9.5 | 3.7 | 1.3 | 0.0 |
| **Guatemala** | 42.1 | 40.8 | 35.5 | 28.2 | 20.5 | 13.2 | 7.0 | 2.6 | 0.0 |
| **Guyana** | 29.0 | 28.8 | 24.8 | 18.7 | 12.2 | 6.7 | 2.8 | 1.0 | 0.0 |
| **Haiti** | 38.5 | 39.0 | 36.2 | 29.3 | 21.6 | 14.2 | 8.3 | 3.7 | 0.0 |
| **Honduras** | 34.2 | 33.9 | 29.7 | 23.0 | 16.1 | 9.7 | 4.5 | 1.7 | 0.0 |
| **India** | 33.0 | 33.0 | 30.1 | 23.6 | 16.4 | 9.4 | 4.1 | 1.5 | 0.0 |
| **Indonesia** | 38.8 | 38.8 | 34.9 | 27.8 | 20.4 | 13.2 | 7.4 | 3.0 | 0.0 |
| **Malaysia** | 33.9 | 33.8 | 29.8 | 21.7 | 13.4 | 5.5 | 0.6 | 0.0 | 0.0 |
| **Mexico** | 36.5 | 36.2 | 32.3 | 24.9 | 17.2 | 10.1 | 4.5 | 1.6 | 0.0 |
| **Nicaragua** | 39.3 | 38.6 | 33.9 | 26.5 | 18.9 | 11.7 | 5.7 | 2.0 | 0.0 |
| **Panama** | 39.5 | 39.3 | 35.6 | 27.8 | 19.4 | 11.2 | 4.8 | 1.6 | 0.0 |
| **Paraguay** | 41.6 | 41.3 | 36.4 | 28.5 | 20.4 | 12.3 | 6.2 | 2.2 | 0.0 |
| **Peru** | 45.0 | 44.6 | 39.6 | 31.6 | 23.4 | 15.1 | 7.5 | 2.6 | 0.0 |
| **Philippines** | 35.9 | 35.9 | 32.8 | 26.0 | 18.5 | 11.2 | 5.6 | 2.3 | 0.0 |
| **Republic of Congo** | 31.3 | 31.3 | 28.6 | 21.9 | 14.4 | 7.4 | 3.0 | 1.3 | 0.0 |
| **St. Lucia** | 42.1 | 41.9 | 36.6 | 27.9 | 19.0 | 10.4 | 4.0 | 1.3 | 0.0 |
| **Sudan** | 22.4 | 22.6 | 20.5 | 16.5 | 12.0 | 7.8 | 4.3 | 2.0 | 0.0 |
| **Thailand** | 40.6 | 40.5 | 36.9 | 28.7 | 20.0 | 11.6 | 4.9 | 1.7 | 0.0 |

*DALY monetization*

DALYs were monetised using country-specific health opportunity costs estimated by Ochalek *et al.*[60], in which DALYs averted for a 1% change in health expenditure are calculated by the product of the estimated DALY burden for that country and the estimated elasticity. Although they were estimated in 2015, as elasticities for such estimation are calculated at the country level with only difference in the interaction of measures of infrastructure and donor funding, there is little indication that elasticities would change over time[60]. For countries included in our study but not reflected in those estimates (Grenada, St. Lucia, Republic of Congo, Djibouti, Democratic Republic of Congo), we used the methodology for DALY monetisation at the level of country income group from another study by the same authors [61]. For high and upper middle income countries, the cost per DALY was estimated as 0.5% of the GNI per capita in the country in 2023; for lower middle and low income countries, we used the cost per DALY by income class estimated by Ochalek *et al.* [61]. As the cost per DALY estimates from Ochalek *et al*. were not estimated in 2023, we inflated it to 2023 I$ using Turner's approach described above[49].

Table SS21. Estimated costs per DALY for each country.

| Approach | Income level | Country | Cost per DALY (I$ 2023) |
|---|---|---|---|
| **From Ochalek *et al.*[60]** | High income | Guyana | 4,897 |
| | High income | Panama | 35,839 |
| | Upper middle income | Belize | 8,623 |

| Approach | Income level | Country | Cost per DALY (I$ 2023) |
|---|---|---|---|
| | Upper middle income | Brazil | 18,339 |
| | Upper middle income | Colombia | 29,833 |
| | Upper middle income | Costa Rica | 33,360 |
| | Upper middle income | Dominican Republic | 10,393 |
| | Upper middle income | Ecuador | 15,420 |
| | Upper middle income | Guatemala | 9,667 |
| | Upper middle income | Indonesia | 5,013 |
| | Upper middle income | Mexico | 2,624 |
| | Upper middle income | Malaysia | 18,791 |
| | Upper middle income | Peru | 19,089 |
| | Upper middle income | Paraguay | 12,161 |
| | Upper middle income | El Salvador | 13,370 |
| | Upper middle income | Thailand | 19,766 |
| | Upper middle income | Venezuela | 11,485 |
| | Lower middle income | Bolivia | 9,475 |
| | Lower middle income | Honduras | 898 |
| | Lower middle income | Haiti | 483 |
| | Lower middle income | India | 6,479 |
| | Lower middle income | Cambodia | 1,554 |
| | Lower middle income | Nicaragua | 8,844 |
| | Lower middle income | Philippines | 2,633 |
| | Low income | Ethiopia | 542 |
| | Low income | Sudan | 790 |
| | Low income | Chad | 865 |
| **Based on Ochalek *et al*. approach [GNI x 0.5][61]** | Upper middle income | Grenada | 8,827 |
| | Upper middle income | St. Lucia | 12,404 |
| **Based on Ochalek et al's population weighted by region[61]** | Lower middle income | Republic of Congo | 748 |
| | Lower middle income | Djibouti | 952 |
| | Low income | Democratic Republic of Congo | 358 |

_Value of statistical life_

The conceptual idea underlying the value of statistical life (VSL) method is to estimate an individual's willingness to pay for a reduction in the probability of dying, which is then averaged across all individuals to estimate the value of preventing one death. To produce global estimates of the VSL, we applied the methodology developed in 2019 by a global expert group of benefit-cost researchers[62]. The reference point of this approach is the recently updated U.S. Department of Health & Human Services VSL estimate of $13.2 million (2023 US$)[63]. Due to the limited availability of direct VSL estimates in LMICs, we applied the value-transfer method outlined in the Reference Case Guidelines for Benefit-Cost Analysis in Global Health and Development.

We used the following formula to compute the VSL of all non-U.S. countries:

$$VSL_{x} = VSL_{USA} \times (GNI\,pC_{x} \div GNI\,pC_{USA})^{E}$$

where VSLx is the unknown VSL of country x, VSL USA is the VSL of the United States ($13.2 million), and GNI pC is the 2023 gross national income per capita (PPP) of country x or the United States, as applicable. GNI pC data were obtained from the World Bank[57]. E denotes the income elasticity of VSL, i.e., the percentage change in VSL associated with a 1% change in real income. The value-transfer method adjusts VSL from the US setting to lower-income settings based on the income ratio between the target and reference countries. The ratio is raised to the income elasticity for the value of reducing mortality risk, which is estimated at 1.2 for middle-income countries, 1.5 for low-income countries, and 1.0 for other high-income countries. World Bank 2023 definitions of country income group classifications were used throughout our analysis.

We also calculated value of statistical life-year (VSLY) lost due to chikungunya mortality, which weights VSL towards individuals dying at younger ages to reflect societal preference for averting deaths in younger relative to older individuals.[62] Following the reference case for cost-benefit analysis in LMICs, we calculated VSLY lost due to chikungunya mortality using the country-specific VSL, life expectancy at birth (LE) and years of life lost (YLLs) due to chikungunya mortality.

$$VSLY_c = \frac{VSL_c}{\frac{LE_c}{2}} \times YLLs$$

Table SS22. Estimated value of statistical life for each country.

| Country | Value of statistical life (I$ 2023) |
|---|---|
| **Belize** | 2.74M |
| **Bolivia** | 3.24M |
| **Brazil** | 5.13M |
| **Cambodia** | 1.53M |
| **Chad** | 135K |
| **Colombia** | 7.99M |
| **Costa Rica** | 6.58M |
| **Democratic Republic of Congo** | 76.5K |
| **Djibouti** | 1.38M |
| **Dominican Republic** | 7.23M |
| **Ecuador** | 4.33M |
| **El Salvador** | 3.00M |
| **Ethiopia** | 226K |
| **Grenada** | 3.25M |
| **Guatemala** | 3.73M |
| **Guyana** | 21.5M |
| **Haiti** | 433K |
| **Honduras** | 1.44M |
| **India** | 4.11M |
| **Indonesia** | 5.37M |
| **Malaysia** | 15.2M |
| **Mexico** | 6.58M |
| **Nicaragua** | 2.39M |
| **Panama** | 13.0M |
| **Paraguay** | 5.29M |
| **Peru** | 4.03M |
| **Philippines** | 3.69M |
| **Republic of Congo** | 1.80M |
| **St. Lucia** | 5.14M |
| **Sudan** | 103K |
| **Thailand** | 9.36M |

#### *2i. Vaccine-associated adverse events and burden*

In the base-case analysis, we included vaccine-associated adverse events for the hypothetical vaccine. We assumed that large-scale preventive vaccination programmes would only be implemented if the overall risk of severe or fatal neurological outcomes due to vaccination would be negligibly small. Therefore, the base-case analysis included only age-specific probabilities of serious adverse events (SAEs) reported by Valneva for IXHCIQ (7 SAEs/32,949 vaccinees aged 18-64 and 22 SAEs/18,445 vaccinees aged 65+)[64], assuming adolescents aged 12-17 have the same probability of events as those aged 18-64.

Similar to Daniel *et al*, SAEs were assumed to have the same duration, disability weight and healthcare costs as medically attended mild chikungunya[65]. No additional vaccine-associated mortality or long-term disability beyond that captured by these SAEs was included in the base-case analysis.

Table SS23. Base-case parameters for vaccine-associated safety outcomes.

| | Probability of vaccine-associated serious adverse events (95%CI) | Source |
|---|---|---|
| **Age 12-17** | 0.000212 (0.000085, 0.000396) | Assume the same as age 18-64 |
| **Age 18-64** | 0.000212 (0.000085, 0.000396) | Daniel *et al*[65], Valneva for VLA1553[64] |
| **Age 65+** | 0.001193 (0.000748, 0.001740) | |

In a scenario analysis, the hypothetical vaccine was assumed to have the broader safety profile reported for the live-attenuated VLA1553 vaccine based on evidence from randomised controlled trials[66-68] and post-marketing surveillance[69].

Acute adverse events were informed by the randomised trial findings[66-68]. Injection-site reactions were excluded, as their transient nature was assumed unlikely to result in measurable functional health loss captured by DALYs. We reviewed acute adverse events on fever, headache, fatigue, myalgia, arthralgia and rash, as these were consistently reported across trials, were biologically plausible vaccine-related reactions, and occurred more frequently among vaccine recipients than controls.

Because these symptoms commonly occur concurrently, we sought to avoid double counting adverse events by modelling only myalgia and arthralgia, and using the larger of the two reported event counts as the total number of acute vaccine-associated adverse events. Events were stratified into mild, moderate and severe categories according to the trial data. For outcomes reported only as non-severe and severe, non-severe cases were further divided into mild and moderate categories, assuming that 88% of total cases were mild, consistent with the distribution assumed for acute chikungunya symptoms in the base-case analysis. Acute symptom duration was assumed to be 4 days, based on VLA1553 package leaflet[70]. Disability weight and healthcare costs were assigned by matching symptom severity to the corresponding severity categories of acute chikungunya symptoms.

Chronic or prolonged adverse events were informed by post-marketing surveillance and stratified into moderate and severe categories[69]. Both categories were assumed to incur the same disability weights and healthcare costs as chronic chikungunya symptoms. Symptom duration was assumed to be 94 days for moderate cases, corresponding to the median duration of 94 days observed for prolonged symptoms in post-marketing surveillance, and 180 days for severe cases, reflecting the reported duration of at least six months for the most persistent chronic symptoms[69]. In a further scenario analysis, we assumed no vaccine-associated safety outcomes.

Table S24. Scenario analysis parameters for vaccine-associated adverse events.

| **Safety outcome** | **Age** | **Probability (95%CI)** | **Duration** | **Disability weight**^ |
|---|---|---|---|---|
| ***Myalgia / Arthralgia*** | | | | |
| *Mild* | 12-17 | 0.1197 (0.0706, 0.1796) | 4 days | 0.006 (0.002. 0.012) |
| | 18-64 | 0.0617 (0.0532, 0.0709) | 4 days | 0.006 (0.002. 0.012) |
| | 65+ | 0.0557 (0.0341, 0.0822) | 4 days | 0.006 (0.002. 0.012) |
| *Moderate* | 12-17 | 0.0549 (0.0351, 0.0787) | 4 days | 0.110 (0.074, 0.145) |
| | 18-64 | 0.0070 (0.0043, 0.0104) | 4 days | 0.110 (0.074, 0.145) |
| | 65+ | 0.0064 (0.0009, 0.0171) | 4 days | 0.110 (0.074, 0.145) |
| *Severe* | 12-17 | 0* | 4 days | 0.583 (0.403, 0.739) |
| | 18-64 | 0.0014 (0.0004, 0.0029) | 4 days | 0.583 (0.403, 0.739) |
| | 65+ | 0.0012 (0.0000, 0.0066) | 4 days | 0.583 (0.403, 0.739) |
| ***Chronic symptoms*** | | | | |
| *Moderate* | 12+& | 0.00454 (0.00249, 0.00720) | 94 days | 0.229 (0.216, 0.242) |
| *Severe* | 12+& | 0.00032 (0.00001, 0.00120) | 180 days | 0.229 (0.216, 0.242) |

^ matching the disability weight of corresponding chikungunya disease symptoms; * No excess risk in the treatment arm; & Data from people age 18+, assume the same pattern for people age 12-17.
